# Contacts and behaviours of university students during the COVID-19 pandemic at the start of the 2020/21 academic year

**DOI:** 10.1101/2020.12.09.20246421

**Authors:** Emily Nixon, Adam Trickey, Hannah Christensen, Adam Finn, Amy Thomas, Caroline Relton, Clara Montgomery, Gibran Hemani, Jane Metz, Josephine G. Walker, Katy Turner, Rachel Kwiatkowska, Sarah Sauchelli, Leon Danon, Ellen Brooks-Pollock

## Abstract

CONQUEST (COroNavirus QUESTionnaire) is an online survey of contacts, behaviour, and COVID-19 symptoms for University of Bristol (UoB) staff/students. We analysed survey results from the start of the 2020/2021 academic year, prior to the second national lockdown (14/09/2020-01/11/2020), where COVID-19 outbreaks led to lockdown of some student halls of residence. The aim of these analyses was to enhance knowledge of student contact patterns to inform infection disease mathematical modelling approaches.

Responses captured information on demographics, contacts on the previous day, symptoms and self-isolation during the prior week, and COVID-19 status.

740 students provided 1261 unique records. Of 42 (3%) students testing positive in the prior fortnight, 99% had been self-isolating. The median number of contacts on the previous day was 2 (interquartile range: 1-5), mode: 1, mean: 6.1; 8% had ≥20 contacts. 57% of student contacts were other UoB students/staff.

Most students reported few daily contacts but there was heterogeneity, and some reported many. Around 40% of student contacts were with individuals not affiliated with UoB, indicating potential for transmission to non-students/staff.

## Background

By November 2020, the COVID-19 pandemic had caused 1.2 million deaths globally(1) and in many countries had forced the temporary closure of educational institutions, including universities(2). In the Autumn of 2020, with reported daily COVID-19 cases rising nationally(3) students at UK universities began to return for the start of the 2020/2021 term. Whilst university students, due to their young age, are less affected by COVID-19 morbidity and mortality than other groups(4), up to one third still may be medically vulnerable to severe COVID-19(5) and all infected students still have the potential to transmit the virus to others. University students often travel from across the country and the globe to their place of education and raise the possibility of onward transmission of infection carried from their home locations. In addition to the national COVID-19 restrictions in place during Autumn 2020, UK universities implemented a range of measures to reduce transmission such as reducing the amount of in person teaching through delivery of lectures online and restricting student living circles(6). However, despite these measures, large outbreaks of COVID-19 occurred across many UK universities(6, 7).

At the University of Bristol (UoB), there was an online induction week from 28^th^ September to the 2^nd^ October and the first teaching block started on the 30^th^ September. The UoB adopted a “blended” teaching approach based upon a mixture of in-person and online teaching. In university-owned halls of residence, students were divided into households (“living circles”) ranging from 1 to 44 individuals per household (median = 5, interquartile range [IQR]: 1-7)(8). Students were instructed not to host any non-residents in their household but could meet others outside of their household provided they conformed to the government social distancing guidelines and other relevant infection control measures such as use of face coverings where appropriate to do so(9).

The UoB has reported positive test results daily since the 14^th^ October(10), with 1722 positive tests among UoB students being reported up until the 1^st^ November, roughly 7% of students, compared with 48 positive tests among staff (<1%) over the same period. On the 9^th^ October, 300 students in one University-owned hall of residence were requested to begin a 14-day period of mass self-isolation(11) and then on the 13^th^ October an additional 40 students in a block of four flats in a separate location were also asked to start a 14-day period of self-isolation(12). The vast majority of students living in these large halls of residence are first year undergraduates(13). Students that tested positive in other accommodation types are required to isolate along with their household, in line with national guidelines(10).

Although there have been previous studies prior to the COVID-19 pandemic that have collected data on contact patterns(14-17), only a small sample of these were relevant to students(14, 15) or participants could not be identified as students(16, 17). Furthermore, the behaviour of students may have changed in light of the pandemic and in response to government regulations. Understanding more about the contact patterns, COVID-19 related symptoms and behaviour of students is important to inform public health action and mathematical models. We aimed to investigate the reported behaviours and contact patterns among students of the UoB during the start of the 2020/2021 academic term by carrying out an online survey.

## Methods

CONQUEST (COroNavirus QUESTionnaire) is an ongoing survey on contacts, behaviour, and potential SARS-Cov-2 symptoms for staff and students at UoB. This survey has been live since the 23^rd^ June 2020. Participants completed an initial questionnaire which included questions on background demographics and then were given the option to fill out a shorter version of the questionnaire on contacts, symptoms, and whether they had had COVID-19; repeating this every 8 days. Initially there was high participation from staff members, but very low participation from students, principally because the survey date started near the end of the 2019/2020 academic year when most students had returned home. From the start of the 2020/2021 academic year, there were a number of initiatives to recruit more students to complete the survey. Here, we present a subset of the survey data from the 14^th^ September 2020 to the 1^st^ November 2020, mainly focusing on the student data.

### Survey

The survey data were collected and managed using REDCap Electronic Data Capture tools hosted at the UoB(18, 19). The survey captured demographic information, information about participant’s contacts on the previous day, information about symptoms during the previous week, whether participants had been self-isolating during the previous week, and COVID-19 status if known.

Demographic information on participants was captured when they completed the initial survey. This included data on age, gender, ethnicity, whether they were part of a high-risk group, whether they were a student, a member of staff, or both, whether they were an undergraduate or postgraduate, their study year, their UoB department, their residence, and the age of their household members.

Participants were asked about three types of contacts they had had on the previous day:

1. Individual contacts - those who they spoke to in person one-on-one, including those in their household and support bubble.
2. Other contacts - if they spoke in person to many people one-on-one in the same setting (but they did not have the opportunity to speak to each other), for example, as part of working in a customer service role in a shop.
3. Group contacts - large groups of individuals in the same setting (for example, sports teams, tutorials, lectures, religious services, large gatherings with friends and family).

For “individual” contacts (contact type 1), participants were asked about where this contact was made, whether this contact was indoors, outdoors or both, the duration of this contact, whether this contact involved touch, whether this contact studied/worked at the university (and if so which faculty and school they were associated with), their age, whether they were part of their household, and how often they would usually have contact with this person.

For “other” contacts (contact type 2), no additional questions were asked, as it was expected that there often would be a large number of “other” contacts and participants would not be motivated to answer additional questions on them.

For “group” contacts (contact type 3), participants were asked how many individuals this involved, their ages, whether the majority were from UoB (and if so the main faculty and school this group was associated with), where the group met, whether this was indoors, outdoors or both, whether the members of the group talked to each other and how long the contact with this group was for.

Additionally, participants were asked about symptoms in the last 7 days (listed in Table 5), whether they had sought medical attention for these symptoms, whether they had been self-isolating in the last 7 days, and their COVID-19 status. For some analyses, the variable on whether people have had COVID-19 (no, yes confirmed by a test, yes a doctor suspected so, yes my own suspicions) was combined with the date that they had been tested or were suspected to have COVID-19. This was to create new variables on whether they had COVID-19 in the two weeks prior to survey completion, or before this.

Participants who had signed up to repeat questionnaires were sent an email every 8 days with a unique link that allowed their responses to be anonymously connected to those from previous CON-QUEST questionnaires that they had responded to. The reminder emails with the survey links were sent every 8 days regardless of whether participants had filled in surveys from previous reminder emails or when they responded to them.

The full questionnaire has been provided in the supplementary materials. The anonymised raw data can be accessed upon request by contacting the corresponding author.

### Analyses

We include records from the 14th September 2020 - 1st November 2020 in order to capture student behaviours at the beginning of term. For some analyses, a comparator population of staff (not including those listed as staff/students) was created taking the same survey dates. We calculated the mean prevalence of behaviours, symptoms, or contacts, stratified by population subgroups.

To investigate the associations between the overall number of contacts on the previous day and demographics and behaviours, univariable and multivariable negative binomial regression modelling was used. These models included variables on: age group (17-24, 25-44, 45-64, 65-79, 80+ years of age), gender (male, female/other - the “other” category had too few individuals and so were grouped with the largest category), under/postgraduate status, current study year (1, 2, 3, 4+), symptoms during the previous week, cardinal symptoms (loss of taste or smell, fever, persistent cough(20)) during the previous week, self-isolating in the prior week, self-reporting being in a high-risk group, household size (1, 2-3, 4-5, 6-9, 10+, missing), and COVID-19 status (never had, previously thought they had it, previously tested positive for it, thought they had it in the last 2 weeks, tested positive for it in the last 2 weeks). Note that all postgraduates were assigned to the 4+ year group to differentiate them from undergraduates in their first year of study.

### Weighting

Initial analyses suggested males and undergraduates were underrepresented in the survey responses. We therefore weighted analyses, with weights based on publicly available UoB data on student demographics, to make the dataset more representative of the university’s student population - see Supplementary table 1. All tables specify whether weighting was used.

**Table 1:** Unweighted and weighted demographics of the 740 student participants and 1261 student records.

| Characteristic | N (%) participants |  | N (%) records unweighted |  | N (%) records weighted |  |
| --- | --- | --- | --- | --- | --- | --- |
| **Age** |  |  |  |  |  |  |
| 17-24 | 557 | (75.3%) | 857 | (68.0%) | 994 | (78.8%) |
| 25-44 | 168 | (22.7%) | 368 | (29.2%) | 225 | (19.4%) |
| 45-64 | 12 | (1.6%) | 27 | (2.1%) | 17 | (1.4%) |
| 65-79 | 3 | (0.4%) | 9 | (0.7%) | 5 | (0.4%) |
| **Gender** |  |  |  |  |  |  |
| Female | 520 | (70.3%) | 868 | (68.8%) | 675 | (53.6%) |
| Male | 207 | (28.0%) | 368 | (29.2%) | 564 | (44.8%) |
| Other/prefer not to say | 13 | (1.8%) | 25 | (2.0%) | 21 | (1.7%) |
| **Ethnicity** |  |  |  |  |  |  |
| White | 559 | (75.5%) | 1003 | (79.5%) | 1004 | (79.7%) |
| Mixed/multiple ethnic groups | 33 | (4.5%) | 57 | (4.5%) | 56 | (4.5%) |
| Asian/Asian British | 117 | (15.8%) | 163 | (12.9%) | 160 | (12.7%) |
| Black/African/Caribbean/Black British | 7 | (1.0%) | 7 | (0.6%) | 6 | (0.5%) |
| Other/prefer not to say | 24 | (3.2%) | 31 | (2.5%) | 34 | (2.7%) |
| No/don't know/other | 664 | (89.7%) | 1113 | (88.3%) | 1137 | (90.2%) |
| Yes | 76 | (10.3%) | 148 | (11.7%) | 124 | (9.8%) |
| **Student type** |  |  |  |  |  |  |
| Undergraduate | 474 | (64.1%) | 725 | (57.5%) | 931 | (73.9%) |
| Postgraduate | 266 | (34.0%) | 536 | (42.5%) | 330 | (26.2%) |
| **Year group** |  |  |  |  |  |  |
| 1 | 180 | (24.3%) | 260 | (20.6%) | 344 | (27.3%) |
| 2 | 122 | (16.5%) | 205 | (16.3%) | 247 | (19.6%) |
| 3 | 95 | (12.8%) | 156 | (12.4%) | 199 | (15.8%) |
| 4+ | 343 | (46.4%) | 640 | (50.8%) | 470 | (37.3%) |
| **Household size** |  |  |  |  |  |  |
| 1 | 117 | (15.8%) | 227 | (18.0%) | 170 | (13.5%) |
| 2-3 | 245 | (33.1%) | 449 | (35.6%) | 430 | (34.1%) |
| 4-5 | 194 | (26.2%) | 323 | (25.6%) | 362 | (28.7%) |
| 6-9 | 107 | (14.5%) | 153 | (12.1%) | 192 | (15.3%) |
| 10+ | 26 | (3.5%) | 36 | (2.9%) | 45 | (3.6%) |
| Unknown | 51 | (6.9%) | 73 | (5.8%) | 61 | (4.8%) |
| **Residence** |  |  |  |  |  |  |
| Catered halls | 24 | (3.2%) | 34 | (2.7%) | 44 | (3.5%) |
| Self-catered halls | 161 | (21.8%) | 228 | (18.1%) | 280 | (22.2%) |
| Shared house/flat | 349 | (47.2%) | 613 | (48.7%) | 642 | (51.0%) |
| Live with family | 105 | (14.2%) | 196 | (15.5%) | 156 | (12.4%) |
| Live alone | 52 | (7.0%) | 96 | (7.6%) | 75 | (6.0%) |
| Other | 49 | (6.6%) | 94 | (7.5%) | 63 | (5.0%) |

### Ethical approval

Ethical approval was granted on the 14th May 2020 by the Health Sciences University Research Ethics Committee at the University of Bristol (ID = 104903), with four amendment requests approved on the 22nd May 2020, 9th June 2020, 27th August 2020 and 7th September 2020. The purpose of the amendments were either to update the relevance of the questions or to make the survey faster and easier to complete.

## Results

### Demographics

From the 14th September 2020 to the 1st November 2020 there were 740 students that completed the questionnaire 1261 times. For a comparator population there were 1655 records from 433 staff.

Most students were aged 17-24, with a median age of 21 (IQR: 19-24) years and a mean age of 23.3 (standard deviation [SD] = 6.8) years. 26.2% of our student sample (42.5% before weighting) were postgraduates, so many participants were aged 25-64. 37 (3%) of the students also listed themselves as staff. 59.3% of our sample lived in households of 2-5 people. First years have higher mean and maximum household sizes (8.0 - SD: 30.4, max: 400) compared to the other years: 4.3 (SD: 2.4, max: 14), 3.9 (SD: 2.5, max: 20), 3.1 (SD: 4.3, max: 60), for years 2, 3, and 4+, respectively (Table 1).

### Symptoms and behaviours

437 (35%) had had symptoms in the week prior to survey, and 93 (7%) had cardinal symptoms, whilst 179 (14%) student participants had been self-isolating in the week prior to the survey (Table 2). Of those with symptoms, 30 (7%) sought medical attention (this could have included: contacting NHS 111, a pharmacist or GP/Practice nurse; visiting a walk-in centre, Accident and Emergency or other hospital). 152 (12%) students thought that they had had COVID-19 (but did not report having had a positive test) more than two weeks prior to filling in the survey, whilst 20 (2%) had tested positive more than two weeks prior to the survey. 56 (4%) students thought that they had had (but had not tested positive for) COVID-19 within the two weeks before completing the survey. 42 (3%) of respondents had tested positive in the two weeks prior to survey completion. Students in their first year of study more commonly reported isolating and having cardinal COVID-19 symptoms in the last 7 days before taking the survey, compared to students not in their first year (24% and 15%, respectively), and having tested positive for COVID-19 in the two weeks before the survey (10%), than the overall student sample (14% isolating, 7% with cardinal symptoms, and 3%, testing positive).

**Table 2:** Percentage (95% confidence intervals) of student participants isolating within the prior week, with symptoms within the prior week, or suspected of having/testing positive for COVID-19 (all weighted), overall and stratified by study year.

|  | Study year |  |  |  | Overall |
| --- | --- | --- | --- | --- | --- |
|  | 1 | 2 | 3 | 4+ |  |
| **Isolating in the prior 7 days, N=179** | 24%<br>(20-29%) | 13%<br>(9-18%) | 11%<br>(6-15%) | 9%<br>(6-11%) | **14%**<br>**(12-16%)** |
| **Symptoms in the prior 7 days, N=437** | 44%<br>(38-49%) | 36%<br>(30-42%) | 30%<br>(24-37%) | 29% (25-33%) | **35%**<br>**(32-37%)** |
| **Cardinal symptoms in the prior 7 days, N=93** | 15%<br>(11-19%) | 6%<br>(3-9%) | 3%<br>(1-6%) | 4%<br>(2-6%) | **7%**<br>**(6-9%)** |
| **Seeking medical attention for reported symptoms, N=30** | 3%<br>(1-5%) | 3%<br>(1-5%) | 1%<br>(0-2%) | 2%<br>(1-3%) | **2%**<br>**(2-3%)** |
| **Suspected of having COVID-19 more than 2 weeks before survey\*, N=152** | 9%<br>(6-12%) | 16%<br>(11-20%) | 9%<br>(5-14%) | 13%<br>(10-16%) | **12%**<br>**(10-14%)** |
| **Suspected of having COVID-19 last 2 weeks before survey\*, N=56** | 5%<br>(3-7%) | 8%<br>(4-11%) | 6%<br>(3-10%) | 2%<br>(0-3%) | **4%**<br>**(3-6%)** |
| **Tested COVID positive more than 2 weeks before survey, N=20** | 3%<br>(1-5%) | 2%<br>(0-4%) | 0%<br>(0-1%) | 1%<br>(0-1%) | **2%**<br>**(1-2%)** |
| **Tested COVID positive last 2 weeks before survey, N=42** | 10%<br>(7-13%) | 2%<br>(0-4%) | 0%<br>(0-0%) | 1%<br>(0-1%) | **3%**<br>**(2-4%)** |

\*Medical professional's opinion or personal suspicion

Table 3 presents the most common symptoms in the last week reported by students, stratified by their COVID-19 status. All of those that had tested positive in the two weeks prior to the survey reported at least one symptom in the prior week but none of these participants reported chilblains, vomiting, or unusual abdominal pain. The most common symptoms among those that had tested positive in the two weeks before the survey were a runny nose/sneezing (73%), loss or altered sense of smell (59%), a headache (53%), unusual fatigue (51%), loss or altered sense of taste (49%), and a sore throat (42%). Meanwhile, 36% reported a fever, and 35% a persistent cough; both considered cardinal symptoms of COVID-19.

**Table 3:** Number and percentage of students with symptom type within the week before survey completion, stratified by COVID-19 status

| Symptom | No COVID-19 (N=992) | Tested positive more than two weeks before survey (N=20) | Think they have had COVID-19 more than two weeks before survey* (N=152) | Think they have had COVID-19 within prior two weeks before survey* (N=56) | Tested positive within the prior two weeks before survey (N=42) |
| --- | --- | --- | --- | --- | --- |
| None | 688 (69%) | 11 (56%) | 96 (63%) | 29 (51%) | 0 (0%) |
| Fever | 14 (1%) | 2 (11%) | 6 (4%) | 1 (2%) | 15 (35%) |
| Persistent cough | 23 (2%) | 0 (0%) | 2 (2%) | 9 (15%) | 14 (34%) |
| Unusual shortness of breath | 13 (1%) | 5 (26%) | 3 (2%) | 5 (9%) | 3 (8%) |
| Unusual chest pain or chest tightness | 18 (2%) | 2 (10%) | 2 (2%) | 3 (5%) | 7 (16%) |
| Unusual abdominal pain | 16 (2%) | 2 (11%) | 5 (3%) | 3 (5%) | 0 (0%) |
| Confusion, disorientation or drowsiness | 13 (1%) | 0 (0%) | 4 (3%) | 3 (6%) | 4 (8%) |
| Headache | 106 (11%) | 1 (5%) | 16 (11%) | 16 (28%) | 22 (52%) |
| Runny nose/sneezing | 157 (16%) | 5 (26%) | 25 (17%) | 13 (23%) | 31 (73%) |
| Unusual fatigue | 53 (5%) | 0 (0%) | 6 (4%) | 12 (21%) | 21 (51%) |
| Sore throat | 114 (11%) | 1 (5%) | 17 (11%) | 13 (24%) | 18 (42%) |
| Unusual muscle aches | 19 (2%) | 0 (0%) | 6 (4%) | 5 (10%) | 9 (20%) |
| Diarrhoea | 27 (3%) | 3 (15%) | 6 (4%) | 3 (5%) | 5 (11%) |
| Vomiting | 4 (0%) | 0 (0%) | 1 (1%) | 2 (3%) | 0 (0%) |
| Loss or altered sense of taste | 1 (0%) | 0 (0%) | 6 (4%) | 6 (11%) | 21 (49%) |
| Loss or altered sense of smell | 2 (0%) | 0 (0%) | 4 (3%) | 6 (11%) | 24 (58%) |
| Chilblains on toes or hands | 8 (1%) | 0 (0%) | 4 (3%) | 0 (0%) | 0 (0%) |
| Any unexpected rashes | 6 (1%) | 2 (11%) | 0 (0%) | 0 (0%) | 3 (7%) |

\*Medical professional's opinion or personal suspicion

Those with cardinal symptoms in the week prior to taking the survey were far more likely to have been isolating in that week (61%) than those without these symptoms (11%). 99% of those that had tested positive for COVID-19 during the two weeks before survey completion had been isolating within the last week (Table 4). 81% of those that had tested positive for COVID-19 during the two weeks prior to the survey had had the cardinal COVID-19 symptoms within the week prior to the survey and 14% of these had sought medical treatment. Of those that suspected that they had had COVID-19 during the two weeks prior to the survey but that had not received a positive test, 52% had been self-isolating and 21% reported having the cardinal COVID-19 symptoms within the week prior to the survey.

**Table 4:** Percentage (and standard deviation) of students reporting behaviours or COVID-19 characteristics (weighted), stratified by other behaviours and characteristics

| Group | Isolating in the prior 7 days | Symptoms in the prior 7 days | Cardinal symptoms in the prior 7 days | Sought medical attention for reported symptoms | Suspected having COVID-19 more than 2 weeks before survey* | Suspected having COVID-19 prior 2 weeks before survey* | Tested positive for COVID-19 more than 2 weeks before survey | Tested COVID-19 positive prior 2 weeks before survey |
| --- | --- | --- | --- | --- | --- | --- | --- | --- |
| No symptoms within prior week (N=824) | 9% (29%) | 0% (0%) | 0% (0%) | 0% (0%) | 12% (32%) | 3% (18%) | 1% (12%) | 0% (0%) |
| Symptoms within prior week (N=437) | 24% (43%) | 100% (0%) | 21% (41%) | 7% (25%) | 13% (33%) | 6% (24%) | 2% (14%) | 10% (29%) |
| No cardinal symptoms within prior week (N=1168) | 11% (31%) | 29% (46%) | 0% (0%) | 2% (13%) | 12% (32%) | 4% (19%) | 2% (12%) | 1% (8%) |
| Cardinal symptoms within prior week (N=93) | 61% (49%) | 100% (0%) | 100% (0%) | 12% (33%) | 13% (33%) | 12% (33%) | 2% (15%) | 36% (48%) |
| Not had COVID (N=992) | 9% (28%) | 31% (46%) | 3% (18%) | 2% (13%) | 0% (0%) | 0% (0%) | 0% (0%) | 0% (0%) |
| Suspected having COVID-19 more than two weeks before survey* (N=152) | 13% (34%) | 36% (48%) | 8% (27%) | 2% (14%) | 100% (0%) | 0% (0%) | 0% (0%) | 0% (0%) |
| Suspected having COVID-19 prior 2 weeks before survey* (N=56) | 52% (50%) | 48% (50%) | 21% (41%) | 5% (22%) | 0% (0%) | 100% (0%) | 0% (0%) | 0% (0%) |
| Tested positive for COVID-19 more than two weeks before survey (N=20) | 21% (42%) | 45% (51%) | 10% (31%) | 5% (22%) | 0% (0%) | 0% (0%) | 100% (0%) | 0% (0%) |
| Tested positive for COVID-19 prior 2 weeks before survey (N=42) | 99% (11%) | 100% (0%) | 81% (40%) | 14% (35%) | 0% (0%) | 0% (0%) | 0% (0%) | 100% (0%) |

\*Medical professional's opinion or personal suspicion

**Table 5:**
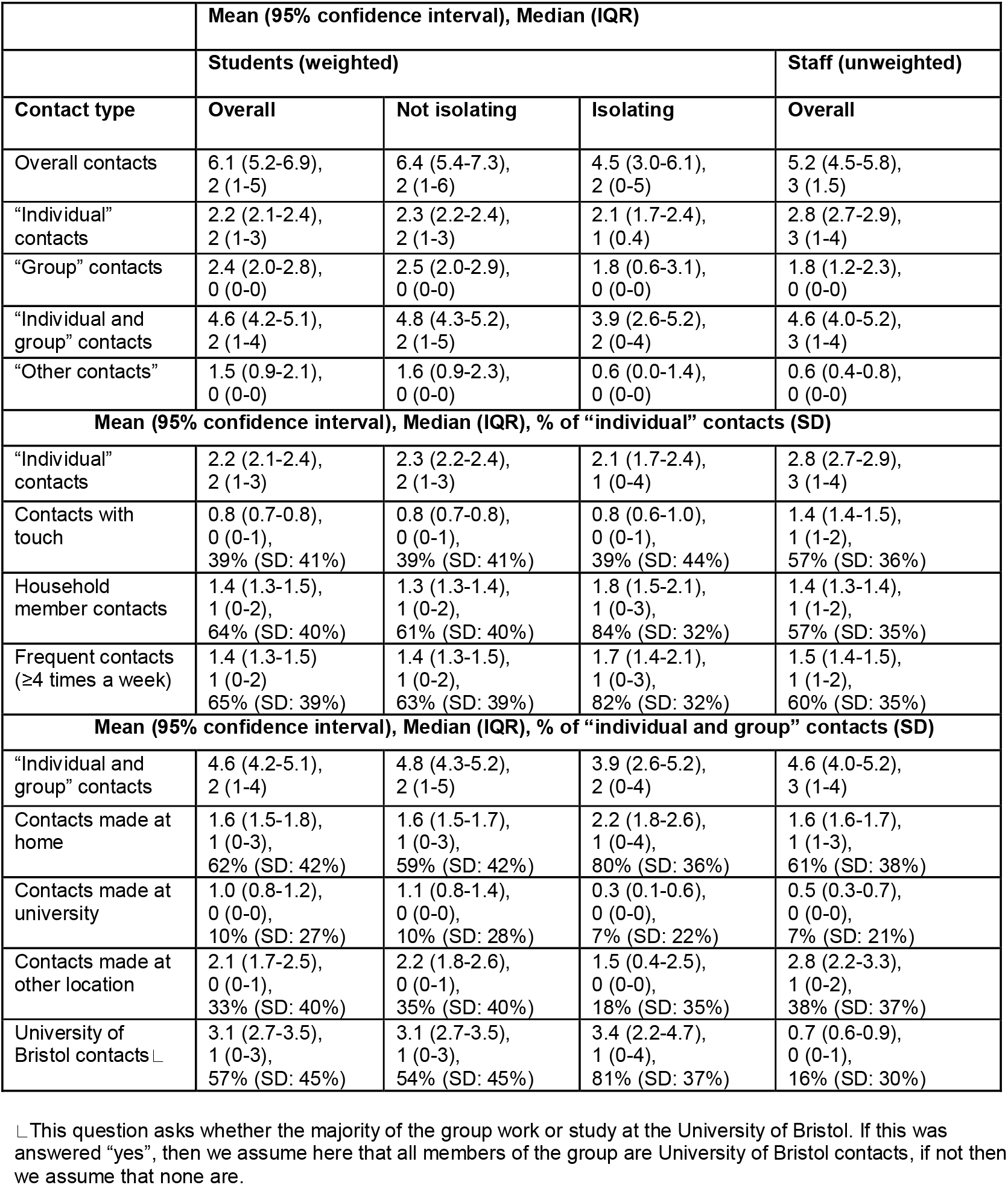
Number of contacts types* overall and stratified by isolation status in the last week for students, and overall for staff. **\*”**Individual” contacts were the people that the participant spoke to in person one-on-one, including those in the participant’s household and support bubble. “Group” contacts were the contacts that the participant had with large groups of individuals in the same setting (for example, sports teams, tutorials, lectures, religious services, large gatherings with friends and family). “Other” contacts were the many people participants spoke to one-on-one in the same setting where the contacts did not have the opportunity to speak to each other (for example, as part of a customer service role in a shop). Not all of the contact types were asked for each category of contacts, so are only comparable to the associated categories indicated here.

### Contacts

The mean number of contacts reported by students for the previous day was 6.1 (SD: 15.0), with a median of 2 (IQR: 1-5). Fewer respondents filled out the survey on Saturdays and Sundays, (10% combined - Supplementary table 2) compared to weekdays, meaning that data are relatively sparse regarding Fridays and Saturdays. Figure 1 and Supplementary figures 1-7 show the distribution of the number of contacts on the previous day for students, staff, and various sub-groups of students, as well as different types of contacts. The weighted mean number of responses where participants had 20 or more contacts on the previous day was 8% (SD: 27%). Numbers of contacts reported for the previous day are shown in Supplementary figure 8, stratified by week. The mean number of contacts appears to be higher from the 5th October onwards; however, there were few survey responses during the first 3 weeks.

**Figure 1:**
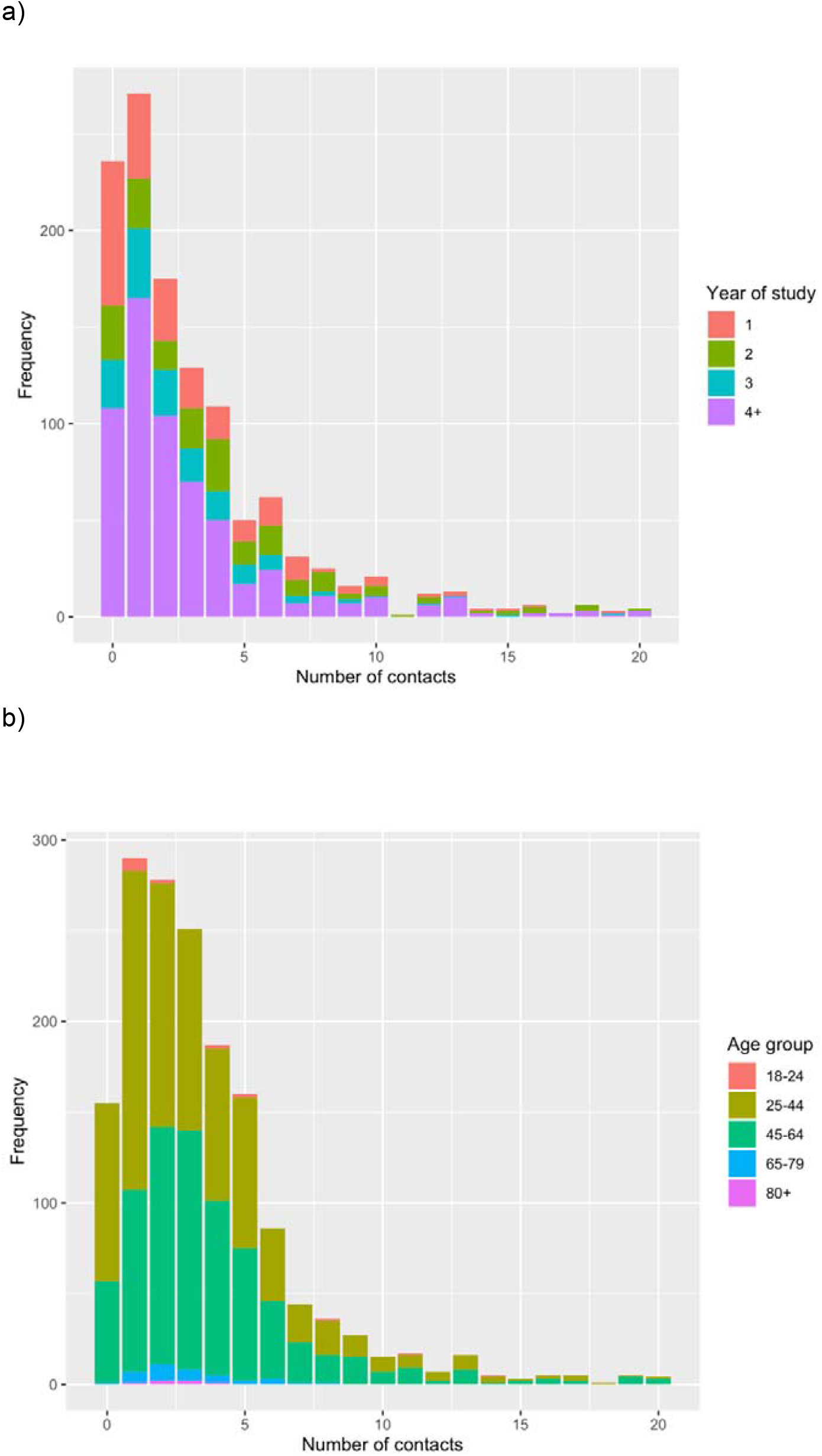
Unweighted histograms of the number of overall contacts* on the previous day among **a)** students (including staff/students); **b)** staff (excluding staff/students) *81 students had more than 20 contacts on the previous day; 58 staff had more than 20 contacts on the previous day - full histograms are shown in supplementary figure 1.

Supplementary table 3 presents a matrix of the mean contacts for the students on the previous day by age-group, with the majority of contacts happening among their own age groups for those aged 18-24 and 25-44. Of the 1261 survey responses, 63 (5%) recorded a contact with someone aged 65 or older, with 27 of these occurring among those aged 17-24, 27 among those aged 25-44, 8 among those aged 45-64, and 1 among those aged 65-79.

The number of contacts on the previous day and the proportion of participants isolating within the last week by residence type are shown in Figure 2. Whilst 31% and 29% of those in catered and self-catered halls, respectively, (the majority of which will be first years) were isolating within the last 7 days, the mean number of contacts on the previous day appeared higher in the self-catered halls (5.6) than in the catered halls (2.3). Those living in other accommodation types were less likely to have been isolating in the prior week. Participants living with their family appeared to have had the highest mean number of contacts on the previous day (7.5).

**Figure 2:**
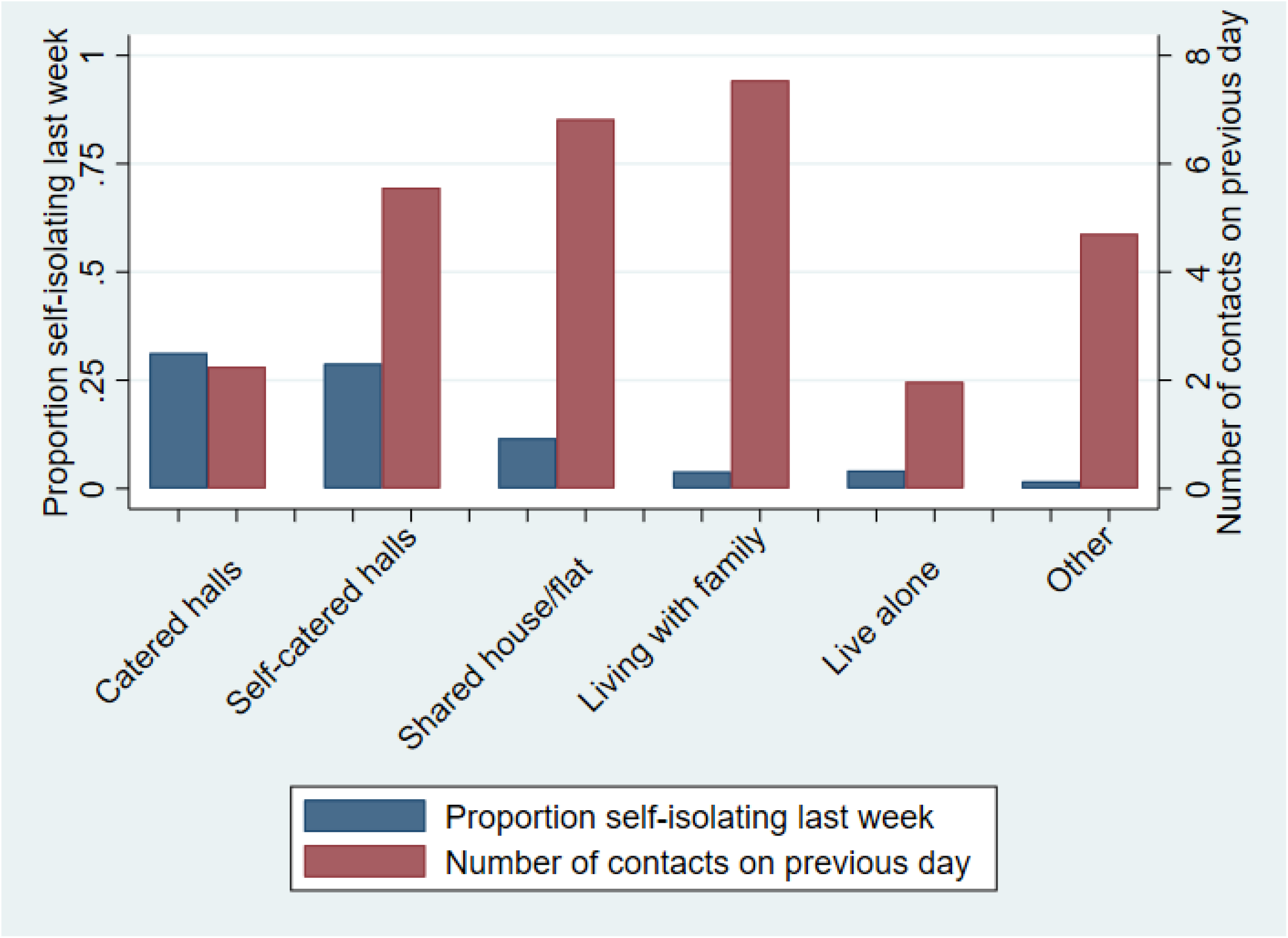
Mean number of contacts on the previous day and the proportion of people isolating within the prior week by residence type

Students that reported isolating within the last week had a lower mean number of contacts on the previous day (4.5) than those not isolating (6.4) (Table 5). The number of “individual” contacts appeared to be similar between those not isolating (2.3) and those isolating (2.1), however the “group” contacts were higher among those not isolating (2.5) than those isolating (1.8), as were “other” contacts (1.6 vs 0.6). Staff had lower mean numbers of overall contacts on the previous day than students (5.2 vs 6.1), which was driven by having lower numbers of “group” (1.8 vs 2.4) and “other” contacts (0.6 vs 1.5).

The mean percentage of “individual” contacts on the previous day that involved touch was 39% (SD: 41.0%) overall, 35% (SD: 42%) for males, and 42% (SD: 41%) for females. Overall, the mean percentage of “individual” contacts on the previous day that were with household members was 64% (Table 5). There was a higher percentage of household contacts on the previous day for those who had been isolating within the last 7 days, than for those who had not been isolating within the last 7 days (84% and 61% respectively). Similar results are seen for the percentage of contacts that were frequent (where the person would usually meet that particular contact ≥4 times a week) as for those seen for household contacts. 62% of “individual and group” contacts on the previous day were made at the home of the respondent, and this percentage was lower among those not isolating within the last 7 days (59%) than among those that had been isolating (80%). Whilst the percentage of contacts on the previous day made at the university were similar between those that had and had not been isolating within the last 7 days (10% vs 7%), the percentage of contacts at other locations was higher among those that had not been isolating in the prior week (35%) than those that had been isolating (18%). 57% of “individual and group” contacts on the previous day were with other UoB students or staff - this percentage was lower among those not isolating within the past week (54%) than those isolating (81%). In comparison to students, staff had a higher number of contacts on the previous day that involved touch (57% for staff vs 39% for students). Similar numbers of their “individual and group” contacts on the previous day were made at home for staff (61%) and students (62%), whilst far fewer of the contacts of staff on the previous day were either UoB staff or students (16% for staff vs 54% for students). The mean percentage of the student’s “individual” non-UoB contacts that were household members was 50%.

Participants that had not been isolating in the prior week had shorter mean contact durations with their contacts at home (3.3 hours) than those that had been isolating (3.9 hours), and longer durations of their contacts on the previous day in a location other than home or university (1.1 vs 0.3 hours), with both groups have a similar duration of contacts at university (0.2 vs 0.3 hours).

In unweighted analyses looking at repeat records from participants, there were 37 records where a participant self-reported not isolating in the 7 days before one survey completion date but then isolating in the 7 days before their next survey completion. For these records, the mean number of contacts was 7.1 (SD: 7.1) for the first survey (when not isolating) and 8.4 (SD: 15.4) at the second (when isolating). There were 20 records where participants went from isolating to not isolating, where the mean number of contacts on the previous day went from 8.7 (SD: 19.6) at the first survey to 9.2 (SD: 13.3) at the second.

There were 17 records where a participant reported a new suspected infection or a positive test within the last two weeks, having previously said they had no history of suspected or confirmed infection with COVID-19 (i.e. new cases). For these records, the mean number of contacts on the previous day was 7.8 (SD: 8.2) at the first survey and 6.2 (SD: 6.1) at the second. Only 6 individuals reported current infection, and subsequently reported a previous infection at the next survey. The mean number of contacts reported by these individuals was 3.9 (SD: 4.0) at the first survey and 5.6 (SD: 6.1) at the second.

### Regression analysis

In the multivariable regression analysis of the number of contacts for the previous day (Table 6), older ages were associated with a lower number of contacts when compared with those aged 17-24 years. Students in their 4th (or higher) year of study reported higher numbers of contacts for the previous day than students in their 1st year. Reporting the cardinal COVID-19 symptoms within the last week was associated with a higher number of contacts on the previous day (versus not having the cardinal COVID-19 symptoms), whilst isolating within the week before the survey was associated with having fewer contacts on the previous day.

**Table 6:** Unweighted negative binomial regression analyses (95% confidence intervals [CI]) of number of contacts on the previous day.

| VARIABLE | N: Mean (SD)<br>contacts | UNIVARIABLE |  | MULTIVARIABLE |  |
| --- | --- | --- | --- | --- | --- |
|  |  | IRR (95%CI) | p-value | IRR (95%CI) | p-value |
| AGE 17-24 | 857: 6.4 (18.3) | Reference | NA | Reference | NA |
| AGE 25-44 | 368: 4.5 (10.6) | 0.71 (0.62, 0.81) | <0.001 | 0.54 (0.45, 0.66) | <0.001 |
| AGE 45-64 | 27: 3.7 (4.4) | 0.57 (0.37, 0.88) | 0.011 | 0.29 (0.18, 0.47) | <0.001 |
| AGE 65-79 | 9: 1.6 (1.0) | 0.24 (0.10, 0.56) | 0.001 | 0.34 (0.13, 0.90) | 0.029 |
| FEMALE/OTHER | 893: 5.5 (15.9) | Reference | NA | Reference | NA |
| MALE | 368: 6.5 (16.7) | 1.18 (1.03, 1.34) | 0.014 | 1.16 (1.01, 1.35) | 0.038 |
| UNDERGRAD | 725: 6.0 (11.1) | Reference | NA | Reference | NA |
| POSTGRAD | 536: 5.5 (21.2) | 0.92 (0.81, 1.04) | 0.171 | 0.76 (0.58, 1.00) | 0.052 |
| STUDY YEAR 1 | 260: 4.4 (8.0) | Reference | NA | Reference | NA |
| STUDY YEAR 2 | 205: 7.5 (10.8) | 1.71 (1.40, 2.09) | <0.001 | 1.11 (0.84, 1.46) | 0.456 |
| STUDY YEAR 3 | 156: 4.7 (8.9) | 1.08 (0.87, 1.35) | 0.480 | 0.76 (0.56, 1.02) | 0.065 |
| STUDY YEAR 4+ | 640: 6.0 (20.7) | 1.38 (1.18, 1.62) | <0.001 | 1.45 (1.08, 1.95) | 0.013 |
| NO SYMPTOMS | 833: 5.6 (17.8) | Reference | NA | Reference | NA |
| SYMPTOMS | 428: 6.2 (12.5) | 1.11 (0.98, 1.26) | 0.100 | 1.26 (1.09, 1.45) | 0.002 |
| NO CARDINAL SYMPTOMS | 1186: 5.7 (15.8) | Reference | NA | Reference | NA |
| CARDINAL SYMPTOMS | 75: 7.3 (21.3) | 1.38 (1.00, 1.64) | 0.052 | 1.62 (1.17, 2.24) | 0.003 |
| NOT ISOLATED LAST WEEK | 1087: 6.0 (16.9) | Reference | NA | Reference | NA |
| ISOLATED LAST WEEK | 167: 4.3 (10.6) | 0.71 (0.60, 0.85) | <0.001 | 0.61 (0.48, 0.76) | <0.001 |
| NOT HIGH RISK | 1113: (5.8 (16.3) | Reference | NA | Reference | NA |
| HIGH RISK | 148: 5.8 (14.9) | 1.02 (0.84, 1.22) | 0.874 | 1.00 (0.81, 1.22) | 0.984 |
| HOUSEHOLD SIZE 1 | 227: 5.9 (16.9) | 1.07 (0.90, 1.28) | 0.420 | 1.24 (1.03, 1.50) | 0.026 |
| HOUSEHOLD SIZE 2-3 | 449: 5.5 (11.9) | Reference | NA | Reference | NA |
| HOUSEHOLD SIZE 4-5 | 323: 7.2 (23.8) | 1.31 (1.12, 1.53) | 0.001 | 1.36 (1.15, 1.61) | <0.001 |
| HOUSEHOLD SIZE 6-9 | 153: 5.7 (9.6) | 1.04 (0.85, 1.26) | 0.733 | 1.27 (1.01, 1.59) | 0.041 |
| HOUSEHOLD SIZE 10+ | 36: 3.3 (5.4) | 0.60 (0.41, 0.88) | 0.010 | 1.23 (0.78, 1.94) | 0.381 |
| HOUSEHOLD SIZE MISSING | 73: 1.8 (4.0) | 0.33 (0.24, 0.44) | <0.001 | 0.49 (0.34, 0.69) | <0.001 |
| NO COVID-19 | 1009: 5.3 (16.3) | Reference | NA | Reference | NA |
| PREVIOUSLY TESTED POSITIVE<br>MORE THAN 2 WEEKS BEFORE<br>SURVEY | 14: 8.2 (10.5) | 1.54 (0.88, 2.69) | 0.130 | 1.30 (0.73, 2.31) | 0.366 |
| PREVIOUSLY SUSPECTED TO BE<br>POSITIVE MORE THAN 2 WEEKS<br>BEFORE SURVEY | 150: 9.2 (19.0) | 1.72 (1.43, 2.06) | <0.001 | 1.53 (1.26, 1.85) | <0.001 |
| SUSPECTED TO BE POSITIVE IN<br>LAST 2 WEEKS | 55: 5.4 (8.4) | 1.01 (0.75, 1.36) | 0.956 | 1.28 (0.93, 1.76) | 0.129 |
| TESTED POSITIVE IN LAST 2<br>WEEKS | 33: 2.9 (3.3) | 0.55 (0.36, 0.81) | 0.003 | 0.55 (0.33, 0.91) | 0.020 |
| CATERED HALLS | 34: 2.0 (3.3) | 0.32 (0.21, 0.48) | <0.001 | 0.34 (0.20, 0.56) | <0.001 |
| SELF-CATERED HALLS | 228: 4.6 (12.6) | 0.73 (0.62, 0.87) | <0.001 | 0.68 (0.52, 0.88) | 0.003 |
| SHARED HOUSE/FLAT | 613: 6.2 (11.5) | Reference | NA | Reference | NA |
| LIVE WITH FAMILY | 196: 8.7 (32.1) | 1.41 (1.19, 1.67) | <0.001 | 1.84 (1.50, 2.25) | <0.001 |
| LIVE ALONE | 96: 2.4 (4.9) | 0.38 (0.30, 0.49) | <0.001 | 0.74 (0.54, 1.00) | 0.051 |
| OTHER | 94: 4.4 (7.5) | 0.70 (0.55, 0.89) | 0.004 | 1.06 (0.80, 1.39) | 0.698 |

In the multivariable regression analysis, participants having a household size of 1 was associated with higher numbers of contacts than participants having a household size of 2-3. Similarly, in comparison to having a household size of 2-3, a household size of 4-5 was associated with more contacts, whilst not reporting household size was associated with reporting fewer contacts. COVID-19 status was associated with number of contacts. Those that had not tested positive for or did not suspect themselves to have had COVID-19 had lower numbers of contacts on the previous day than those that suspected themselves to have had COVID-19 more than two weeks prior to the survey. Those testing positive within the last 2 weeks before survey completion had fewer contacts. Students in catered and self-catered halls had fewer contacts on the previous day then those living in a shared house/flat but students living in a shared house/flat had fewer contacts than those living with their family. Supplementary table 4 shows contact numbers stratified by isolation status and under/postgraduate status, with both undergraduates and postgraduates that had been isolating in the previous week having lower numbers of contacts than that had not been isolating.

## Discussion

Our survey results from the start of the 2020/2021 academic year give insight into the behaviour of students in this unique and important time period in the COVID-19 pandemic. First year undergraduates were more likely to be isolating within the prior 7 days and to have tested positive for COVID-19 in the prior two weeks than other year groups, with higher percentages of respondents isolating that lived in catered and self-catered halls than other accommodation types. This observation confirms that the COVID-19 epidemic among UoB students has been concentrated among first years living in large, shared living residences (as predicted by Brooks-Pollock et al., 2020(13)). There was high compliance to isolation guidelines among students who had a positive test for COVID-19 in the previous two weeks before survey completion, while half of the students who only suspected they had COVID-19 (but did not have this confirmed by a test) isolated. Some of these students may have been required to isolate due to a member of their household or living circle having a positive test, rather than isolating voluntarily. Just over half of those who reported cardinal symptoms self-isolated, indicating that some students that should have been isolating had not been doing so. Students that had been isolating in the prior week had fewer contacts than those that had not been isolating, with a higher percentage of contacts among those isolating being contacts within their home than for those not isolating. This suggests that whilst the number of contacts of the isolating students was often not as low as might be expected, the majority of contacts that took place were with people they lived with, who were also likely to be isolating.

Whilst most students reported a low number of daily contacts on the previous day (mode=1, median=2), 8% of students reported over 20, indicating that these could be individuals with increased likelihood of catching COVID-19 and infecting others (so-called “super spreaders”(21)). Over a third of reported contacts involved touch, whilst around a half of contacts among the students were with other UoB students or staff. When comparing the contacts of students with those of staff, we found that students had slightly higher mean numbers of contacts overall, with the difference driven by having higher numbers of group contacts, possibly due to involvement with university societies, face-to-face teaching (as not all staff are delivering this) and socialising. We also found that staff were far less likely to mix with other university staff and students compared to the students and that staff had higher proportions of contacts that involved touch, possibly as staff are more likely to live with their families. Whilst students appear to mix mostly with other students, our results indicate that there is still potential for considerable crossover of COVID-19 from students to the non-student population.

Our regression analysis results showed that students in their 4^th^ year of study had higher numbers of contacts than those in year 1, despite living in households with fewer members and adjusting for isolation status. This may be due to students in later years already having established social networks that are less disturbed by the COVID-19 guidelines than the nascent social networks being formed by the first years. It could also be because so many first years were isolating that this reduced the number of contacts for first years that were not isolating. Students that reported having had the cardinal COVID-19 symptoms in the last week were more likely to report a higher number of contacts on the previous day than those without these symptoms, which may be due to consistently higher contact patterns leading to infection and exhibiting symptoms. Students in larger households tended to have more contacts than those in households of sizes 2-3, possibly due to an increased pool of readily available contacts, whilst those in one person households also had higher numbers of contacts than those in 2-3 person households, perhaps because they were required to go out to seek social activities. In the multivariable regression analyses, those that tested positive within the 2 weeks prior to survey completion had fewer contacts than those that had not tested positive or suspected themselves to be positive, indicating attempts to adhere with the COVID-19 guidelines. Students living with their family appeared to report the highest number of contacts, with those living in catered and self-catered halls reporting lower numbers of contacts.

Unsurprisingly given the COVID-19 restrictions in place at the time of our survey, we found a lower mean number of daily contacts among our student population (6.1) than was found in the Warwick social contacts survey from 2009(14, 15), either among their entire sample (26.8) or among the students in that sample (29.9). However, that study also found a large amount of heterogeneity in the number of contacts. In the Warwick social contacts survey data, students have more home contacts (3.5) than other participants (2.3), whilst they found that the majority of contacts for students (82%, 95% confidence interval: 79%-86%) were either at home or associated with the university, and that students reported 20 (95% confidence interval: 14.1-28.8) university-related contacts. We also found that a high percentage of student contacts were either at home or university (around 72%). However, we found that both students and our comparator sample of staff had 1.6 contacts at home and that there were only 3.1 university contacts for students on average, with these differences likely due to university and government’s COVID-19 restrictions in place at the time of our survey. Meanwhile, the POLYMOD social contacts survey(17) found a mean of 11.7 contacts per day per person in their Great Britain sample (with an average age around 30), lower than the Warwick social contacts survey, but still much higher than our mean value (6.1). The POLYMOD survey found that people of the same age tended to mix with each other and that people in the main student age group (18-24) have more contacts than older adults, both of which are corroborated by our findings. In the POLYMOD survey, around 75% of contacts at home and 50% of school and leisure contacts involved touch, results that seem closer to our findings for staff (57% of contacts involving touch) than students (39%), which is likely both down to physical distancing measures and due to students being less likely to live with family members than staff or other adults.

The strengths of this survey include the sample size, longitudinal format, and anonymous nature that enable us to capture self-reported behaviours of a large number of students during a key period in the UK’s COVID-19 pandemic. In addition, it provides a unique data source on student behaviour during the pandemic, which will be useful in informing public health action and mathematical models. Our results are likely generalisable to other UK city-based universities, as well as to some city-based universities in other countries which are similar in structure and COVID-19 status to UoB. Many of the questions were designed to be comparable to existing contact surveys(14, 15, 17).

However, this study has some limitations. Firstly, the number of contacts was asked for the previous day, whilst the questions on self-isolation and symptoms asked about the previous week, and a window of 14-days was used to define current COVID-19. This discrepancy in time-windows used for different questions could lead to difficulties in interpreting results, particularly regarding contact patterns for those that had previously been isolating during the prior week but not on the previous day, possibly leading to higher reported contacts for this group. Secondly, the survey questions were devised early in the pandemic when less was known about the epidemiology and possible interventions. We did not capture whether participants had a negative test for COVID-19, which would have been useful information. Thirdly, in order to capture sufficient detail on contacts, the questionnaire is fairly long (5-10 minutes) and complicated, which may deter those with lots of contacts or with little available time from completing it, which may mean it is not representative. Some participants have not filled in their household sizes, which perhaps shows that some people struggled to answer the questionnaires due to the complexity. We include clear instructions defining “contacts” in the survey; however, some people may not read this text or interpret the instructions differently and so there could be variation in what people considered a contact to be.

Selection bias for those who particularly engaged in health-seeking behaviours may have occurred, as those that are less likely to abide by the guidelines may also be less likely to fill out the survey. However, while we are not able to identify the proportion of the population that are not complying with COVID-19 restrictions, we did capture individuals who did not appear to be compliant that were reporting large numbers of contacts and not isolating when experiencing the cardinal symptoms. Another type of selection bias that may have occurred is for students who have had COVID-19. Almost one-fifth of our surveyed student population had tested positive for COVID-19 or suspected that they had had COVID-19, however, only around 7% students had had a positive test as of the 1st November(10). Nevertheless, the true prevalence of COVID-19 in the student population may be greater than 7%, since students with symptoms may not want to present for a test to avoid the potential of obligatory isolation for them and their household. There will inevitably be issues regarding recall bias, particularly when we are asking respondents to estimate when they first think they had COVID-19 (if this hasn’t been confirmed by a positive test), and there will also likely be issues with response bias, leading to inaccurate or false responses.

Our study comes at a crucial time in the COVID-19 pandemic, Autumn 2020, when the disease is resurgent with high numbers of daily cases, including among university students(7). The second national UK lockdown, commenced on the 5th November 2020, has not included the closing of universities(22) and therefore, it is vital that we continue to investigate the contribution of universities to transmission, particularly around Christmas with special measures being implemented for students to return home(23), but with plans for their return post-Christmas currently unclear.

It is important to understand the epidemiology of COVID-19 among students due to high transmission rates and their unique mixing patterns at this time, with thousands of young people moving from all over the country and world to study, forming new social networks in the process. Although the student population is mostly young and therefore unlikely to see the worst effects of COVID-19 infection(4, 24), there is the potential for transmission from students back to their families or to other members of the community. Our study is able to provide novel data on student contacts, symptoms, and behaviours at the beginning of the 2020/21 term when several lockdowns of student residences occurred, enabling us to examine adherence to COVID-19 control measures, as well as the outsized influence on the student COVID-19 pandemic of first year undergraduates that mostly reside in very large accommodation blocks with the potential for large scale indoor transmission(13). This study provides important data for policy makers and mathematical modellers on a key population in the COVID-19 pandemic, as well as crucial data for any future infectious disease outbreaks. We found that the number of daily contacts for students was much lower than in pre-COVID-19 studies, which is likely to be due to the COVID-19 restrictions in place. We show that whilst the majority of students report low numbers of contacts on the previous day, there are a sizeable minority that report large numbers of contacts, highlighting the heterogeneity of transmission and role that individuals with large numbers of daily contacts (potential “super spreaders”) could be having on the spread of disease. Around 40% of student contacts were with people not affiliated to the university, indicating the potential for transmission to groups other than students.

## Data Availability

The datasets generated and analysed during the current study are available from the corresponding author on reasonable request.

## Acknowledgements

We would like to thank the Elizabeth Blackwell Institute for funding this research, our RedCap data manager Alison Horne and our PPI group for their feedback during the development of the survey. We would also like to thank all the participants who have taken part in this study.

## Author contributions

AT and EN wrote the first draft of the manuscript. EN organised data collection, whilst AT analysed the data. EBP had the original idea for the manuscript. HC, AF, ATh, CR, CM, GH, JM, JGW, KT, RK, SS, and LD designed analyses, interpreted the results, and critically reviewed the manuscript.

