## Supplementary data for "Contacts and behaviours of university students during the COVID-19 pandemic at the start of the 2020/21 academic year"

**Supplementary table 1:** Comparison of university data on student demographics with the corresponding CONQUEST survey data demographic information

| **Population** | **University data** http://www.bristol.ac.uk/ssio/statistics/ | **CONQUEST data (including multiple records per respondent)** |
|---|---|---|
| Female, undergraduate | 39.6% | 41.3% |
| Female, postgraduate | 15.7% | 29.5% |
| Male, undergraduate | 34.3% | 16.2% |
| Male, postgraduate | 10.5% | 13.0% |

**Supplementary figure 1:** Histograms of the number of overall contacts on the previous day among **a)** students (including staff/students); **b)** staff (excluding staff/students)

a)

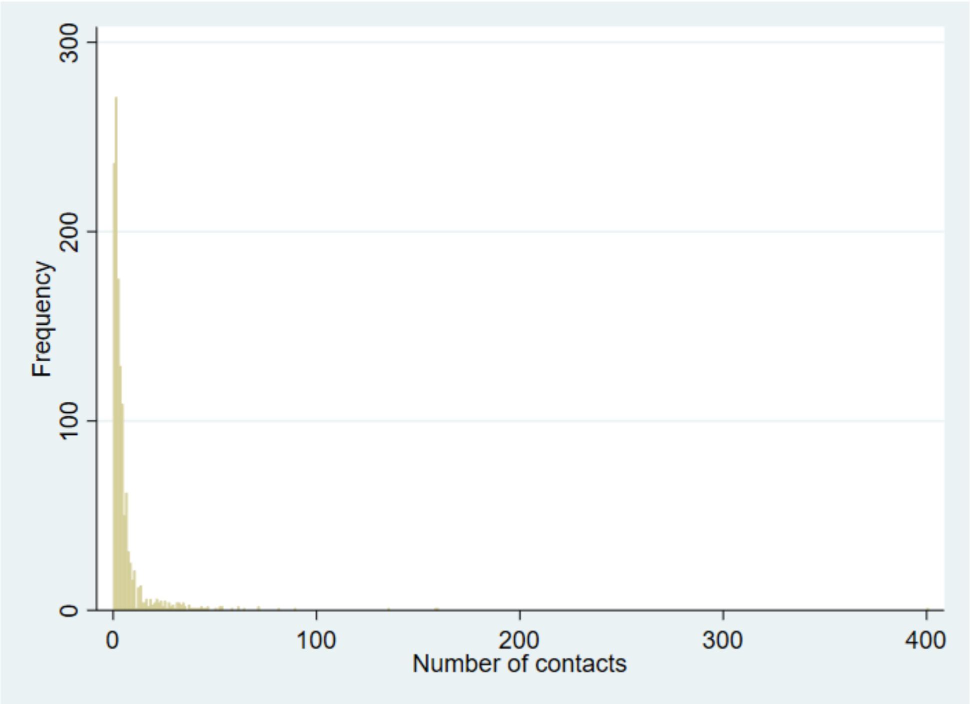


b)

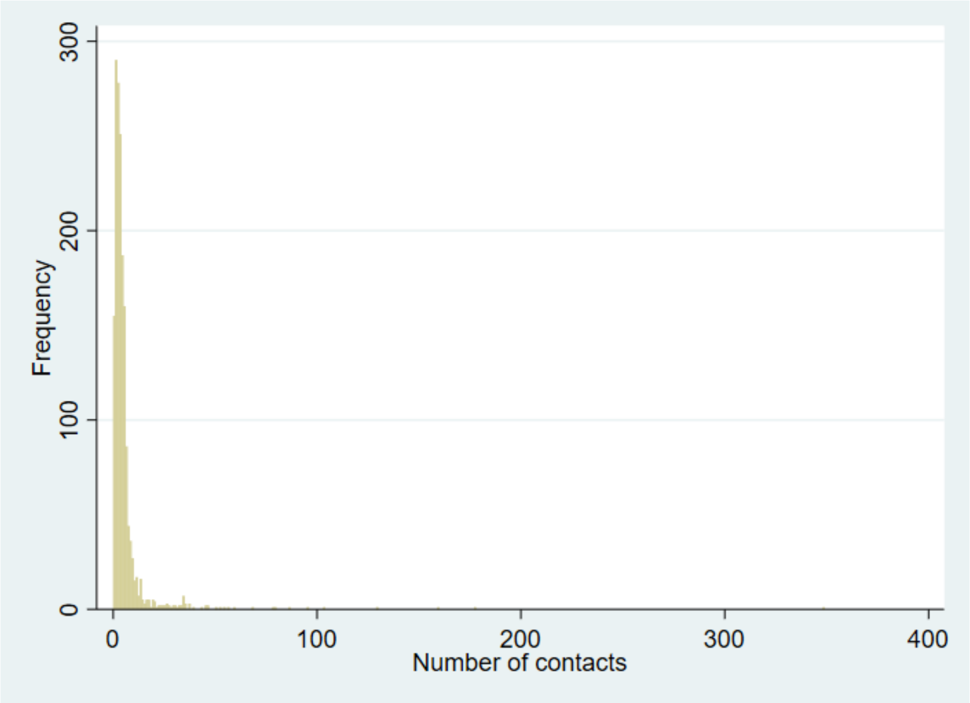

**Supplementary figure 2:** Histogram of the number of contacts on the previous day among students reporting not having the cardinal COVID-19 symptoms (fever, loss or altered taste or smell, persistent cough).

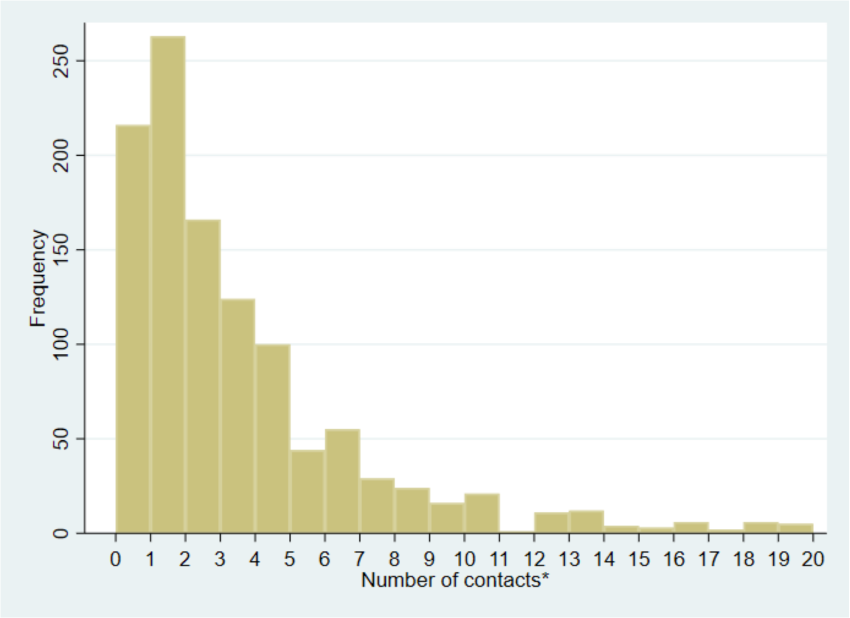


*78 individuals had more than 20 contacts on the previous day

**Supplementary figure 3:** Histogram of the number of contacts on the previous day among students reporting having the cardinal COVID-19 symptoms (fever, loss or altered taste or smell, persistent cough).

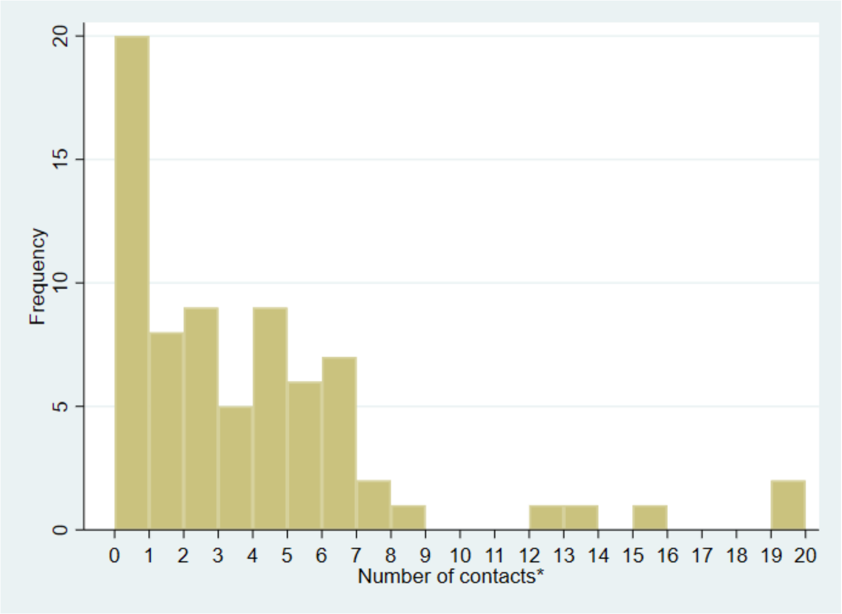


*3 individuals had more than 20 contacts on the previous day

**Supplementary figure 4:** Histogram of the number of “individual contacts” on the previous day among students

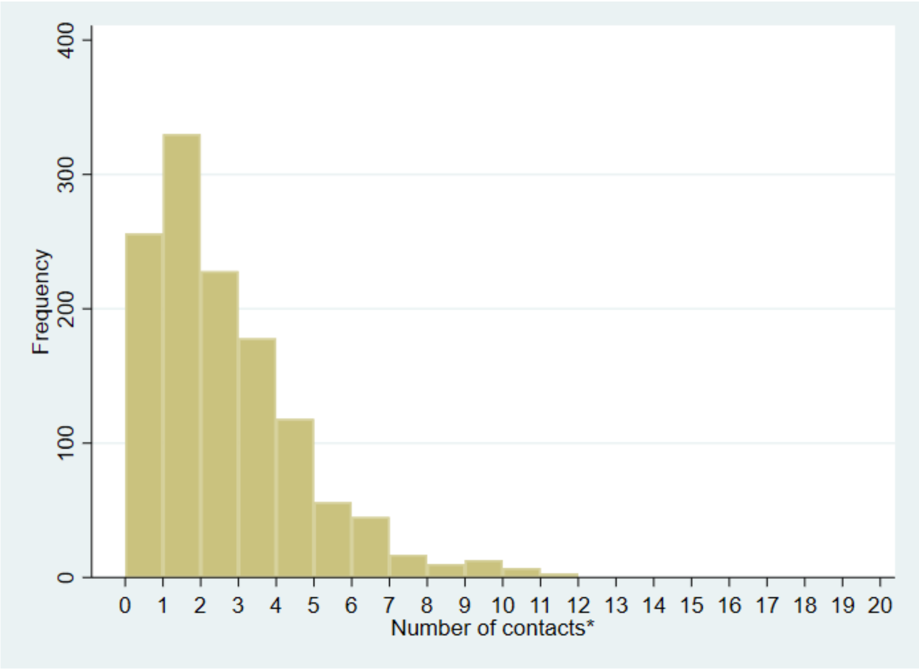


*0 individuals had more than 20 contacts on the previous day

**Supplementary figure 5:** Histogram of the number of “individual contacts” on the previous day among students that involve touch

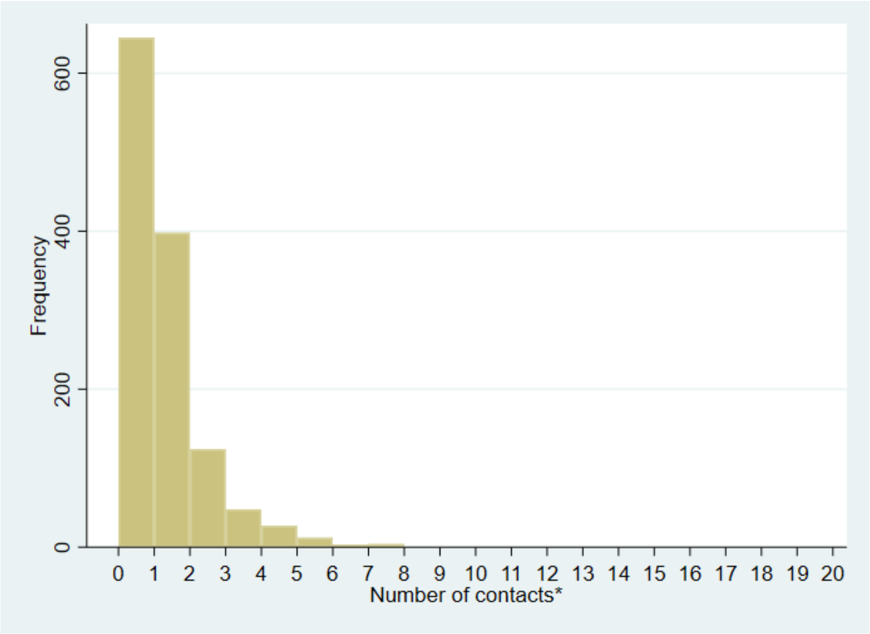


*0 individuals had more than 20 contacts on the previous day

**Supplementary figure 6:** Histogram of the number of overall contacts on the previous day among students not isolating within the last 7 days

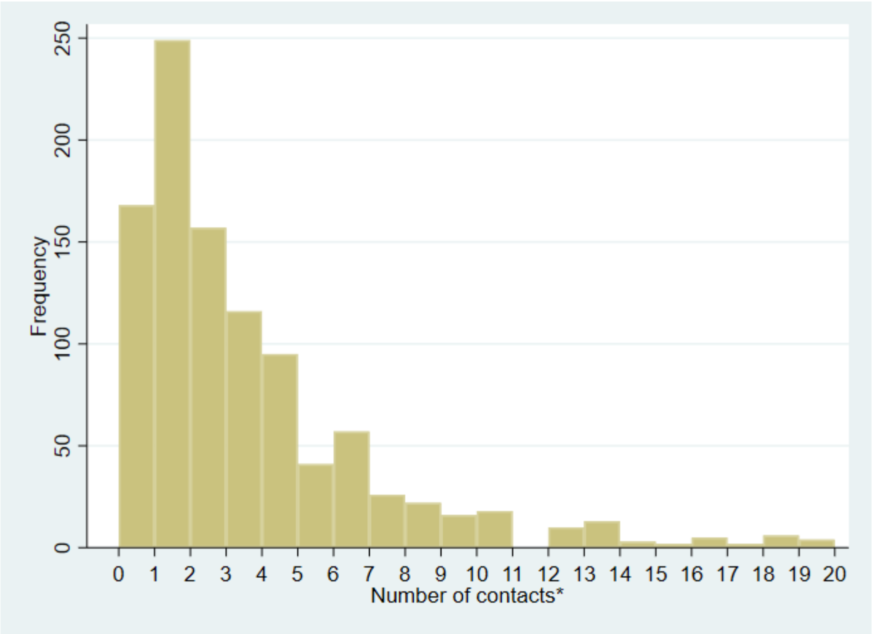


*77 individuals had more than 20 contacts on the previous day

**Supplementary figure 7:** Histogram of the number of overall contacts on the previous day among students isolating within the last 7 days

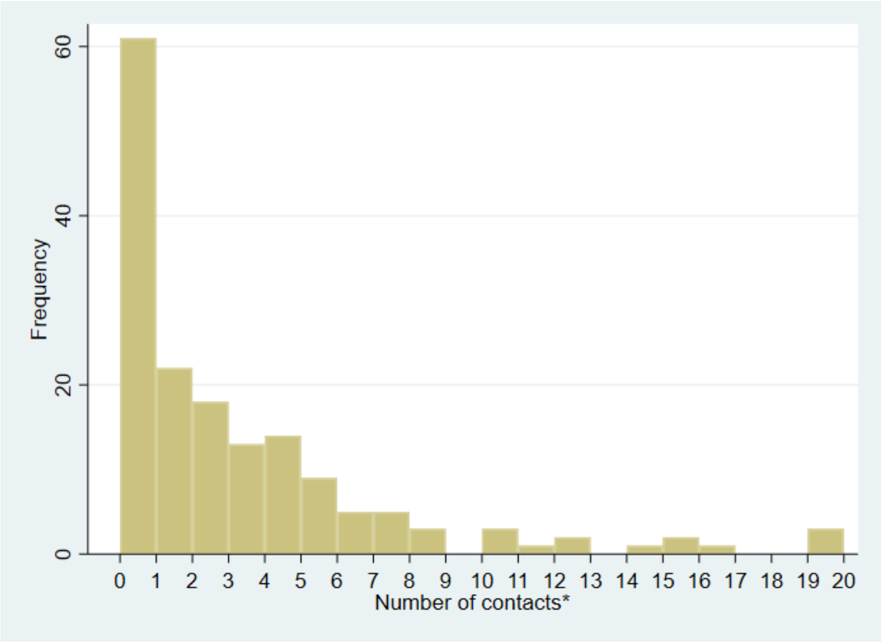


*4 individuals had more than 20 contacts on the previous day

**Supplementary figure 8:** Contacts on the previous day stratified by week (weighted) for students

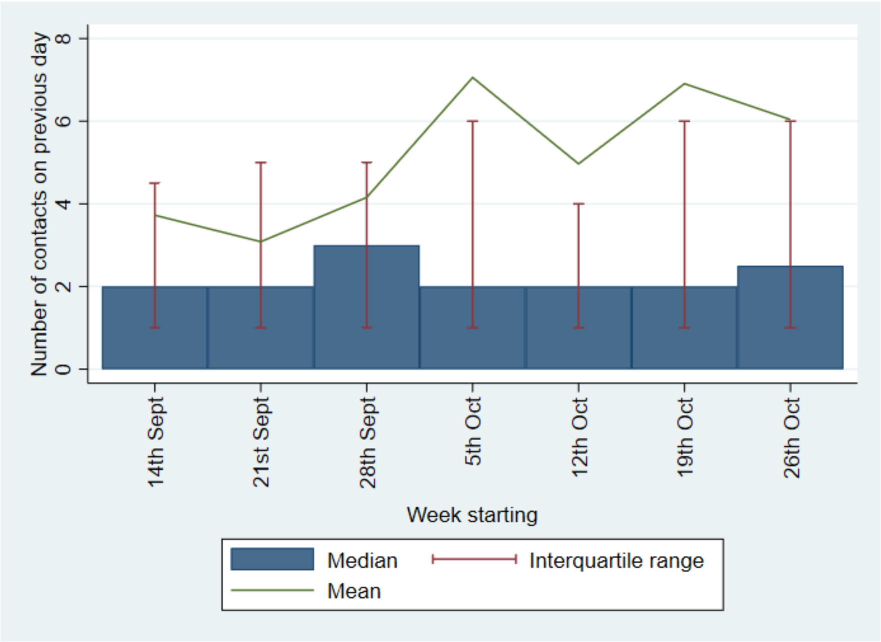

**Supplementary table 2:** Number of participants, and the mean and standard deviation of contacts by weekday.

| **Weekday previous to survey completion** | **N participants: Mean (SD) contacts** |
|---|---|
| Sunday | 265: 6.2 (26.1) |
| Monday | 185: 6.6 (17.4) |
| Tuesday | 190: 5.5 (13.4) |
| Wednesday | 237: 6.0 (11.6) |
| Thursday | 252: 4.8 (7.6) |
| Friday | 78: 5.5 (7.6) |
| Saturday | 54: 5.9 (12.8) |

**Supplementary table 3:** Matrix of mean contacts on the previous day by age (weighted) among students*

| | CONTACT AGE GROUP | | | | | | |
|---|---|---|---|---|---|---|---|
| **PARTICIPANT AGE GROUP** | **0-4** | **5-17** | **18-24** | **25-44** | **45-64** | **65-79** | **80+** |
| **18-24** | 0.006 | 0.159 | 3.978 | 0.511 | 0.239 | 0.030 | 0.002 |
| **25-44** | 0.124 | 0.163 | 0.805 | 1.728 | 0.497 | 0.128 | 0.032 |
| **45-64** | 0.000 | 0.566 | 0.416 | 0.819 | 0.874 | 0.309 | 0.093 |
| **65-79** | 0.000 | 0.000 | 0.222 | 0.667 | 0.556 | 0.111 | 0.000 |

*Contacts can be with people other than students

**Supplementary table 4: Unweighted contacts stratified by under/postgrad and isolation status**

| Group | N: Mean (SD) | Median (IQR) |
|---|---|---|
| Undergrad, not isolating | 598: 6.3 (11.4) | 3 (1-6) |
| Undergrad, isolating | 124: 4.3 (9.1) | 2 (0-5) |
| Postgrad, not isolating | 489: 5.6 (21.8) | 2 (1-4) |
| Postgrad, isolating | 43: 4.3 (14.1) | 1 (0-2) |

**Supplementary data:** The questionnaire

**HIDDEN FIELD**
Survey start date and time - taken from participant's device. ______________________

CON-QUEST (COroNavirus QUESTionnaire): Contact questionnaire on patterns and behaviour in University of Bristol staff and students during the COVID-19 pandemic

Help control COVID-19 Thank you for taking this survey. In doing so, you will be contributing to research into how COVID-19 spreads. Social contact patterns are important for understanding the spread of COVID-19. We are conducting this survey at the University of Bristol to measure social contact patterns in a university setting, as well as other factors important in the transmission of COVID-19. Instructions You must be over 18 years of age to complete this survey and a member of staff or a student at the University of Bristol. The survey is anonymous, and you can opt out at any time by closing the survey. We are unable to remove submitted answers due to the anonymous nature of the survey. Therefore, submission of pages within the survey will be taken as consent that you are happy for the data you have provided to be used for research purposes. Your anonymised data from this questionnaire will be made available publicly through a data repository and through publications to aid with the COVID-19 public health response. As lockdown eases, we will capture how contact patterns and behaviour change over time. If you are happy to be contacted again, you will be able to provide an email address at the end of the survey. Your email address will only be stored for the sole purpose of contacting you about completing this survey in the future. Your email address will not be linked to the information you submit in the questionnaire - your data will remain anonymous. Your information will be kept in accordance with the Data Protection Act. Thank you for completing CON-QUEST! If you have any questions about the survey, then please contact us on:
You can read about the survey and find latest results on the CONQUEST website: bris.ac.uk/con-quest This survey is funded by the Elizabeth Blackwell Institute for Health Research at the University of Bristol.

#### Background information

**HIDDEN FROM SURVEY**
Date and time started (taken from user's device)

__________________________________

The first part of this survey will collect background information about you so that we can understand how representative our survey respondents are compared to the rest of the university population. You will not need to complete this section again if you fill out future versions of the survey.

Age

__________________________________
(Years)

What is your gender?

◯ Female
◯ Male
◯ Other
◯ Prefer not to say

Please specify your gender if you wish __________________________________

How would you describe your ethnicity?

◯ White
◯ Mixed/multiple ethnic groups
◯ Asian/Asian British
◯ Black /African/ Caribbean/Black British
◯ Other ethnic group
◯ Prefer not to say

◯ English / Welsh / Scottish / Northern Irish / British
◯ Irish
◯ Gypsy or Irish traveller
◯ any other white

Please tell us your ethnic group

__________________________________

◯ White and Black Caribbean
◯ White and Black African
◯ White and Asian
◯ Any other mixed/multiple ethnic background

Please tell us your ethnic group

__________________________________

◯ Indian
◯ Pakistani
◯ Bangladeshi
◯ Chinese
◯ Any other Asian background

Please tell us your ethnic group

__________________________________

◯ African
◯ Caribbean
◯ Any other Black/African/Caribbean background

Please tell us your ethnic group

__________________________________

◯ Arab
◯ Any other ethnic group

Please tell us your ethnic group

__________________________________

Are you pregnant?

◯ Yes
◯ No
◯ Not sure

Are you at a higher risk of severe illness if you catch coronavirus?
(see definitions on the NHS website:
https://www.nhs.uk/conditions/coronavirus-covid-19/people-at-higher-risk/whos-at-higher-risk-from-coronavirus/)

◯ Yes, I was sent a letter from the government saying that I am clinically extremely vulnerable (high risk)
◯ Yes, I am in a clinically vulnerable group (moderate risk)
◯ No
◯ Don't know
◯ Other

Other - please explain:

__________________________________

Are you currently living with someone who is shielding?

◯ Yes
◯ No

Are you:
(If you are both then please tick both options)

☐ a student?
☐ a member of staff?

◯ Undergraduate
◯ Postgraduate

What is your main faculty/department

◯ Arts
◯ Engineering
◯ Health Sciences
◯ Life Sciences
◯ Science
◯ Social Sciences and Law
◯ Other

Other - please specify

__________________________________

School

◯ School of Arts
◯ School of Humanities
◯ School of Modern Languages
◯ Centre for Academic Language and Development
◯ Centre for Innovation

School

◯ School of Computer Science, Electrical and Electronic Engineering, and Engineering Mathematics
◯ School of Civil, Aerospace and Mechanical Engineering

School

◯ Bristol Dental School
◯ Bristol Medical School
◯ Bristol Veterinary School
◯ Centre for Health Sciences Education

School

◯ School of Biological Sciences
◯ School of Biochemistry
◯ School of Cellular and Molecular Medicine
◯ School of Physiology, Pharmacology and Neuroscience
◯ School of Psychological Science

School

◯ School of Chemistry
◯ School of Earth Sciences
◯ School of Geographical Sciences
◯ School of Mathematics
◯ School of Physics

School

◯ School of Education
◯ School for Policy Studies
◯ School of Economics, Finance and Management
◯ School of Sociology, Politics and International Studies
◯ University of Bristol Law School

Which academic year of your degree are you currently in (e.g. Year 1 of 4)?

Year

__________________________________

of

__________________________________

Please check - year of study is greater than total number of years!

Year of study

Part time?

◯ Yes
◯ No
◯ Rather not say

What is your job type at the University?

◯ Clinical Academic (e.g. Clinical Lecturer)
◯ Operational Services (e.g. Cleaner, Porter)
◯ Professional/Administrative Services (e.g. Administrator, Director, Manager, Adviser)
◯ Research and Teaching (e.g. Lecturer, Research Associate)
◯ Technical Services (e.g. Research Technician)

Which faculty, directorate or institution are you part of?
(if more than one then please select your main one)

◯ Alumni Campaigns and Relations
◯ Campus Division
◯ Communications and PR
◯ Estates
◯ Faculty of Arts
◯ Faculty of Engineering
◯ Faculty of Health Sciences
◯ Faculty of Life Sciences
◯ Faculty of Science
◯ Faculty of Social Sciences and Law
◯ Finance
◯ HR
◯ Information Technology
◯ Legal services
◯ Library services
◯ Planning and Projects
◯ Research and Enterprise Development
◯ Residential and Hospitality
◯ Secretarial and PA
◯ Senior Management Team
◯ Sports Exercise and Health
◯ Student administration, recruitment and services
◯ Other

---

Which school are you a member of?

◯ School of Arts
◯ School of Humanities
◯ School of Modern Languages
◯ Centre for Academic Language and Development
◯ Centre for Innovation

---

Which school are you a member of?

◯ School of Computer Science, Electrical and Electronic Engineering, and Engineering Mathematics
◯ School of Civil, Aerospace and Mechanical Engineering

---

Which school are you a member of?

◯ Bristol Dental School
◯ Bristol Medical School
◯ Bristol Veterinary School
◯ Centre for Health Sciences Education

---

Which school are you a member of?

◯ School of Biological Sciences
◯ School of Biochemistry
◯ School of Cellular and Molecular Medicine
◯ School of Physiology, Pharmacology and Neuroscience
◯ School of Psychological Science

Which school are you a member of?

◯ School of Chemistry
◯ School of Earth Sciences
◯ School of Geographical Sciences
◯ School of Mathematics
◯ School of Physics

Which school are you a member of?

◯ School of Education
◯ School for Policy Studies
◯ School of Economics, Finance and Management
◯ School of Sociology, Politics and International Studies
◯ University of Bristol Law School

Does the nature of your role mean that the majority of your work must be done on location at the University (i.e. working from home full time is not a feasible option for you)?

◯ Yes - the majority of my work takes place in a laboratory
◯ Yes - my role involves maintaining or monitoring University buildings or grounds
◯ Yes - other reason
◯ No
◯ Not sure

Please specify reason

__________________________________

#### Contacts where you live and work

This section will help us understand more about the contacts that you have in the place that you live.

Where do you currently live during term-time?

◯ Hall of residence (Catered)
◯ Hall of residence (Self- Catered)
◯ Shared house/flat
◯ With immediate family (parents/siblings/children)
◯ Live alone
◯ Other

Other - please specify

__________________________________

Has your term time residence changed as a result of COVID-19?

◯ Yes ◯ No

Where would you usually live during term-time?

◯ Hall of residence (Catered)
◯ Hall of residence (Self- Catered)
◯ Shared house/flat
◯ With immediate family (parents/siblings/children)
◯ Live alone
◯ Other

Other - please specify

__________________________________

Where do you usually live out of term time?

◯ The same place as during term time
◯ Hall of residence (Catered)
◯ Hall of residence (Self- Catered)
◯ Shared house/flat
◯ With immediate family (parents/siblings/children)
◯ Live alone
◯ Other

Other - please specify

__________________________________

*This question is hidden from the survey*
Where do you currently live (now that social distancing has been imposed and the University buildings have closed)?

◯ The same place as during term time
◯ Different halls of residence (Catered)
◯ Different halls of residence (Self- Catered)
◯ Different shared house/flat
◯ With immediate family (parents/siblings/children)
◯ Live alone
◯ Other

Other - please specify

__________________________________

How many people in the following age categories are currently living with you (including yourself)?
If you are living at more than one address currently, then please answer this question for the address where you spend most of your time.

If you live in a hall of residence, we are asking about your living circle (https://www.bristol.ac.uk/students/your-studies/study-2020/living-in-residences/)

Age 0-4:

__________________________________

Age 5-17:

__________________________________

Age 18-24:

__________________________________

Age 25-44:

__________________________________

Age 45-64:

__________________________________

Age 65-80:

__________________________________

Age 81+

__________________________________

*This question is hidden from the survey*Is your household made up of different people as a result of the COVID-19 pandemic

◯ Yes ◯ No ◯ Prefer not to say

*This question is hidden from the survey*
How many people lived in your previous household (including yourself) in the following age categories?

*This question is hidden from the survey*
If you lived in a Halls of residence, we are asking about those who you shared a bathroom/kitchen/living space/dining room with - choose the area which you share with the maximum number of people.

*This question is hidden from the survey*Age 0-4:

__________________________________

*This question is hidden from the survey*Age 5-17:

__________________________________

*This question is hidden from the survey*Age 18-24:

__________________________________

*This question is hidden from the survey*Age 25-44:

__________________________________

*This question is hidden from the survey*Age 45-64:

__________________________________

*This question is hidden from the survey*Age 65-80:

__________________________________

*This question is hidden from the survey*Age 81+

__________________________________

Are you currently in a support bubble with another household? (If you are not sure what a support bubble is, then you can find an official description here:
https://www.ageuk.org.uk/information-advice/coronavirus/coronavirus-guidance/support-bubbles/)

◯ Yes ◯ No

How many people in the following age categories are in your support bubble (not including you or those in your household)?

Age 0-4:

__________________________________

Age 5-17:

__________________________________

Age 18-24:

__________________________________

Age 25-44:

__________________________________

Age 45-64:

__________________________________

Age 65-80:

__________________________________

Age 81+

__________________________________

*This question is hidden from the survey*Due to COVID-19 have you, or would you, consider taking the following action when the University is open?

*This question is hidden from the survey*
Avoid lectures attended by many people:

◯ I have done this
◯ I have not done this, but I intend to do so
◯ I have not done this and do not intend to do so

*This question is hidden from the survey*
Work from home where possible :

◯ I have done this
◯ I have not done this, but I intend to do so
◯ I have not done this and do not intend to do so

*This question is hidden from the survey*
Avoid using shared facilities:

◯ I have done this
◯ I have not done this, but I intend to do so
◯ I have not done this and do not intend to do so

*This question is hidden from the survey*
Wear a face mask or covering when at the University:

◯ I have done this
◯ I have not done this, but I intend to do so
◯ I have not done this and do not intend to do so

*This question is hidden from the survey*
Avoid as much contact as possible by not attending any face-to-face activities on campus:

◯ I have done this
◯ I have not done this, but I intend to do so
◯ I have not done this and do not intend to do so

*This question is hidden from the survey*
Reduce or eliminate social activities involving more than two people:

◯ I have done this
◯ I have not done this, but I intend to do so
◯ I have not done this and do not intend to do so

*This question is hidden from the survey*Are you currently using any COVID-19 symptom tracker or contact tracing apps ?

◯ Yes, and I would consider using similar apps in the future.
◯ Yes, but I would not consider using similar apps in the future.
◯ No, but I would consider using similar apps in the future.
◯ No, and I would not consider using similar apps in the future.

Would you be willing to be tested for current or previous COVID-19 infection using (tick all that apply):

☐ Saliva
☐ Throat swab
☐ Blood test
☐ Finger prick
☐ None of these

### Questionnaire

#### COVID-19 symptoms and responses

◯ stop emails

◯ Stop further emails

**HIDDEN FIELD**
Survey start date and time - taken from participant's device. __________________________________

Which country are you currently living in?

◯ United Kingdom
◯ Afghanistan
◯ Åland Islands
◯ Albania
◯ Algeria
◯ American Samoa
◯ Andorra
◯ Angola
◯ Anguilla
◯ Antarctica
◯ Antigua and Barbuda
◯ Argentina
◯ Armenia
◯ Aruba
◯ Australia
◯ Austria
◯ Azerbaijan
◯ Bahamas
◯ Bahrain
◯ Bangladesh
◯ Barbados
◯ Belarus
◯ Belgium
◯ Belize
◯ Benin
◯ Bermuda
◯ Bhutan
◯ Bolivia
◯ Bosnia and Herzegovina
◯ Botswana
◯ Bouvet Island
◯ Brazil
◯ British Indian Ocean Territory
◯ Brunei Darussalam
◯ Bulgaria
◯ Burkina Faso
◯ Burundi
◯ Cambodia
◯ Cameroon
◯ Canada
◯ Cape Verde
◯ Caribbean Netherlands
◯ Cayman Islands
◯ Central African Republic
◯ Chad
◯ Chile
◯ China
◯ Christmas Island
◯ Cocos (Keeling) Islands
◯ Colombia
◯ Comoros
◯ Congo
◯ Democratic Republic of
◯ Cook Islands
◯ Costa Rica
◯ Côte d'Ivoire
◯ Croatia
◯ Cuba
◯ Curaçao
◯ Cyprus
◯ Czech Republic
◯ Denmark
◯ Djibouti
◯ Dominica
◯ Dominican Republic
◯ Ecuador
◯ Egypt

◯ El Salvador
◯ Equatorial Guinea
◯ Eritrea
◯ Estonia
◯ Ethiopia
◯ Falkland Islands
◯ Faroe Islands
◯ Fiji
◯ Finland
◯ France
◯ French Guiana
◯ French Polynesia
◯ French Southern Territories
◯ Gabon
◯ Gambia
◯ Georgia
◯ Germany
◯ Ghana
◯ Gibraltar
◯ Greece
◯ Greenland
◯ Grenada
◯ Guadeloupe
◯ Guam
◯ Guatemala
◯ Guernsey
◯ Guinea
◯ Guinea-Bissau
◯ Guyana
◯ Haiti
◯ Heard and McDonald Islands
◯ Honduras
◯ Hong Kong
◯ Hungary
◯ Iceland
◯ India
◯ Indonesia
◯ Iran
◯ Iraq
◯ Ireland
◯ Isle of Man
◯ Israel
◯ Italy
◯ Jamaica
◯ Japan
◯ Jersey
◯ Jordan
◯ Kazakhstan
◯ Kenya
◯ Kiribati
◯ Kuwait
◯ Kyrgyzstan
◯ Lao People's Democratic Republic
◯ Latvia
◯ Lebanon
◯ Lesotho
◯ Liberia
◯ Libya
◯ Liechtenstein
◯ Lithuania
◯ Luxembourg
◯ Macau
◯ Macedonia
◯ Madagascar
◯ Malawi
◯ Malaysia
◯ Maldives
◯ Mali
◯ Malta
◯ Marshall Islands
◯ Martinique

◯ Mauritania
◯ Mauritius
◯ Mayotte
◯ Mexico
◯ Federated States of
◯ Moldova
◯ Monaco
◯ Mongolia
◯ Montenegro
◯ Montserrat
◯ Morocco
◯ Mozambique
◯ Myanmar
◯ Namibia
◯ Nauru
◯ Nepal
◯ New Caledonia
◯ New Zealand
◯ Nicaragua
◯ Niger
◯ Nigeria
◯ Niue
◯ Norfolk Island
◯ North Korea
◯ Northern Mariana Islands
◯ Norway
◯ Oman
◯ Pakistan
◯ Palau
◯ State of
◯ Panama
◯ Papua New Guinea
◯ Paraguay
◯ Peru
◯ Philippines
◯ Pitcairn
◯ Poland
◯ Portugal
◯ Puerto Rico
◯ Qatar
◯ Réunion
◯ Romania
◯ Russian Federation
◯ Rwanda
◯ Saint Barthélemy
◯ Saint Helena
◯ Saint Kitts and Nevis
◯ Saint Lucia
◯ Saint Vincent and the Grenadines
◯ Saint-Martin (France)
◯ Samoa
◯ San Marino
◯ Sao Tome and Principe
◯ Saudi Arabia
◯ Senegal
◯ Serbia
◯ Seychelles
◯ Sierra Leone
◯ Singapore
◯ Sint Maarten (Dutch part)
◯ Slovakia
◯ Slovenia
◯ Solomon Islands
◯ Somalia
◯ South Africa
◯ South Georgia and the South Sandwich Islands
◯ South Korea
◯ South Sudan
◯ Spain
◯ Sri Lanka
◯ St. Pierre and Miquelon

◯ Sudan
◯ Suriname
◯ Svalbard and Jan Mayen Islands
◯ Swaziland
◯ Sweden
◯ Switzerland
◯ Syria
◯ Taiwan
◯ Tajikistan
◯ Tanzania
◯ Thailand
◯ The Netherlands
◯ Timor-Leste
◯ Togo
◯ Tokelau
◯ Tonga
◯ Trinidad and Tobago
◯ Tunisia
◯ Turkey
◯ Turkmenistan
◯ Turks and Caicos Islands
◯ Tuvalu
◯ Uganda
◯ Ukraine
◯ United Arab Emirates
◯ United States
◯ United States Minor Outlying Islands
◯ Uruguay
◯ Uzbekistan
◯ Vanuatu
◯ Vatican
◯ Venezuela
◯ Vietnam
◯ Virgin Islands (British)
◯ Virgin Islands (U.S.)
◯ Wallis and Futuna Islands
◯ Western Sahara
◯ Yemen
◯ Zambia
◯ Zimbabwe

---

What is the first part of the postcode where you are currently staying?
(This question is optional but it would be useful to know the range of current locations of the questionnaire participants)

________________________________

What forms of transport did you use yesterday (please select all that apply)?

☐ Walking
☐ Cycling
☐ E-scooter
☐ Motorbike
☐ Car/Van
☐ Taxi
☐ Bus
☐ Train
☐ Aeroplane
☐ Ferry/boat
☐ Did not travel
☐ Other

Other - please state:

__________________________________

Do you think you have had COVID-19 at any time?

◯ Yes, confirmed by a positive test
◯ Yes, suspected by a doctor but not tested
◯ Yes, my own suspicions
◯ No

When were you told/ when did you think you first had COVID-19?

__________________________________

In the last two weeks, have you been in close contact with anyone who may have had COVID-19 during that time period?

◯ Yes, I was in contact with a confirmed COVID-19 case
◯ Yes, I was in contact with a suspected COVID-19 case
◯ No, not to my knowledge

Symptoms in the past seven days

Did you have any of the following symptoms in the past seven days (tick all that apply)?
Even if you believe that these symptoms might be related to a health condition other than COVID-19, please still select the relevant symptoms that you have experienced.

☐ No symptoms
☐ Fever (temperature)
☐ Persistent cough
☐ Unusual shortness of breath
☐ Unusual chest pain or chest tightness
☐ Unusual abdominal pain
☐ Confusion, disorientation or drowsiness?
☐ Headache
☐ Runny nose / sneezing
☐ Unusual fatigue
☐ Sore throat
☐ Unusual muscle aches
☐ Diarrhoea
☐ Vomiting
☐ Loss or altered sense of taste
☐ Loss or altered sense of smell
☐ Chilblains (small, red patches) on toes or hands, sometimes itchy or painful
☐ Any other unexplained rashes

---

Other rashes - please describe where affected

__________________________________

---

Highest recorded temperature in Celsius (if measured)

__________________________________
(°C)

---

Please check - finish date is before start date!

---

When did the symptoms start?

---

__________________________________

---

◯ I can't remember

---

When did the symptoms finish?

---

__________________________________

◯ I can't remember
◯ I still have symptoms

What kind of medical attention did you access?

☐ None
☐ Contacted NHS 111, by phone or online
☐ Visited pharmacist
☐ Spoke to pharmacist on the phone
☐ Consulted GP/practice nurse over the phone or online
☐ Consulted GP/practice nurse face to face
☐ Walk-in centre
☐ Accident and Emergency
☐ Other hospital
☐ Other

Other - please specify:

__________________________________

Have you been self-isolating in the past seven days (have you not left home at all due to you or a household member having COVID-19 symptoms)?

◯ Yes ◯ No

How many days did you self-isolate for in total (or if you are still self-isolating how many total days do you plan to self-isolate for?)

__________________________________

#### Section 6 - Your contacts yesterday

This section is asking about the contacts that you had with other people yesterday including those that you live with. There are three parts to this section of the survey about three different types of contacts you might have had. Read the following instructions carefully so that you know where to enter each contact.
(1) Individual contacts- those who you spoke in person to one-on-one, including those in your household and support bubble.
(2) If you spoke in person to many people one-on-one in the same setting (but they did not have the opportunity to speak to each other), for example, as part of working in a customer service role in a shop.
(3) Large groups of individuals in the same setting (e.g. sports teams, tutorials, lectures, religious services, large gatherings with friends and family).

PLEASE RECORD EACH PERSON ONLY ONCE, on a single row, or as a member of just one group, even if you meet the same person several times during the day and/or as part of a group. If you spoke to an individual in multiple locations, choose the location where you spent the most time with that person.
This survey is anonymous - it is important for you to record all of your contacts and how they happened, whether they fit with the current social/physical distancing advice or not.

Individual contacts yesterday Thinking about yesterday, who did you speak to (in person) yesterday?
It can help to think about your day from when you got up, at breakfast, in the morning, at lunch, in the afternoon, at teatime, and in the evening.
Please include everyone you have been close to. This includes:
People you had a face to face conversation without a complete barrier between you. Please include people you were within 3 meters (10 feet) of and spoke to. Do not include people you talked to through a solid barrier, for example, a closed window. People you had physical contact with for example, handshake, hug, kiss. Examples of these contacts could be your partner, a parcel delivery person, a friend or a shop worker. - Only include individuals you spoke to one-on-one as you will be able to enter those you met in a group in a later section.
- Do include those that are a part of your household.
- If you spoke to many people one-on-one in the same setting (but they did not have an opportunity to speak to each other), for example, as part of a customer service role, you are not required to enter these particular contacts separately but will be prompted to include these in a later question.
- If you spoke to an individual in multiple locations, choose the location where you spent the most time with that person.

◯ I did not speak to anyone in person yesterday on a one-on-one basis.

Person 1 Description of person e.g. "partner" or "delivery person":

______________________________

Age:

◯ 0-4
◯ 5-17
◯ 18-24
◯ 25-44
◯ 45-64
◯ 65-80
◯ 81+

Is this person part of your household?

◯ Yes ◯ No

Does this person work or study at the University of Bristol?

◯ Yes ◯ No ◯ Don't know

What is the main faculty this person is associated with?

◯ Arts
◯ Engineering
◯ Health Sciences
◯ Life Sciences
◯ Science
◯ Social Sciences and Law
◯ Other
◯ Don't know

Which school are they in?

◯ Bristol Dental School
◯ Bristol Medical School
◯ Bristol Veterinary School
◯ Centre for Academic Language and Development
◯ Centre for Health Sciences Education
◯ Centre for Innovation
◯ School for Policy Studies
◯ School of Arts
◯ School of Biochemistry
◯ School of Biological Sciences
◯ School of Cellular and Molecular Medicine
◯ School of Chemistry
◯ School of Civil, Aerospace and Mechanical Engineering
◯ School of Computer Science, Electrical and Electronic Engineering, and Engineering Mathematics
◯ School of Earth Sciences
◯ School of Economics, Finance and Management
◯ School of Education
◯ School of Geographical Sciences
◯ School of Humanities
◯ School of Mathematics
◯ School of Modern Languages
◯ School of Physics
◯ School of Physiology, Pharmacology and Neuroscience
◯ School of Psychological Science
◯ School of Sociology, Politics and International Studies
◯ University of Bristol Law School
◯ Other
◯ Don't know

---

Other - please specify

__________________________________

---

Which school are they in?

◯ School of Arts
◯ School of Humanities
◯ School of Modern Languages
◯ Centre for Academic Language and Development
◯ Centre for Innovation
◯ Don't know

---

Which school are they in?

◯ School of Computer Science, Electrical and Electronic Engineering, and Engineering Mathematics
◯ School of Civil, Aerospace and Mechanical Engineering
◯ Don't know

---

Which school are they in?

◯ Bristol Dental School
◯ Bristol Medical School
◯ Bristol Veterinary School
◯ Centre for Health Sciences Education
◯ Don't know

Which school are they in?

◯ School of Biological Sciences
◯ School of Biochemistry
◯ School of Cellular and Molecular Medicine
◯ School of Physiology, Pharmacology and Neuroscience
◯ School of Psychological Science
◯ Don't know

Which school are they in?

◯ School of Chemistry
◯ School of Earth Sciences
◯ School of Geographical Sciences
◯ School of Mathematics
◯ School of Physics
◯ Don't know

Which school are they in?

◯ School of Education
◯ School for Policy Studies
◯ School of Economics, Finance and Management
◯ School of Sociology, Politics and International Studies
◯ University of Bristol Law School
◯ Don't know

Where were you when you spoke to this person?

◯ Home
◯ Another home
◯ University
◯ Work or volunteering not at university
◯ School or nursery
◯ Shopping
◯ Medical or health centre
◯ Transport
◯ Place of worship
◯ Social (pub/nightclub/café/restaurant)
◯ Exercise
◯ Park
◯ Other location

Other location - please specify

__________________________________

Was this inside or outside?

◯ Inside
◯ Outside
◯ Both inside and outside

How long did you talk to this person for?

◯ Less than 10 minutes
◯ Between 10 minutes and an hour
◯ Between 1 and 4 hours
◯ 4+ hours

Did you also touch this person?

◯ Yes ◯ No

How often would you expect to meet this person under current physical distancing measures?

◯ 4 or more days a week
◯ 2-3 days a week
◯ Once a week
◯ Less often than once a week
◯ Met for the first time this day

Person 2 Description of person:

__________________________________

◯ I did not speak to anyone else in person yesterday on a one-to-one basis.

Age:

◯ 0-4
◯ 5-17
◯ 18-24
◯ 25-44
◯ 45-64
◯ 65-80
◯ 81+

Is this person part of your household?

◯ Yes ◯ No

Does this person work or study at the University of Bristol?

◯ Yes ◯ No ◯ Don't know

What is the main faculty this person is associated with?

◯ Arts
◯ Engineering
◯ Health Sciences
◯ Life Sciences
◯ Science
◯ Social Sciences and Law
◯ Other
◯ Don't know

Which school are they in?

◯ Bristol Dental School
◯ Bristol Medical School
◯ Bristol Veterinary School
◯ Centre for Academic Language and Development
◯ Centre for Health Sciences Education
◯ Centre for Innovation
◯ School for Policy Studies
◯ School of Arts
◯ School of Biochemistry
◯ School of Biological Sciences
◯ School of Cellular and Molecular Medicine
◯ School of Chemistry
◯ School of Civil, Aerospace and Mechanical Engineering
◯ School of Computer Science, Electrical and Electronic Engineering, and Engineering Mathematics
◯ School of Earth Sciences
◯ School of Economics, Finance and Management
◯ School of Education
◯ School of Geographical Sciences
◯ School of Humanities
◯ School of Mathematics
◯ School of Modern Languages
◯ School of Physics
◯ School of Physiology, Pharmacology and Neuroscience
◯ School of Psychological Science
◯ School of Sociology, Politics and International Studies
◯ University of Bristol Law School
◯ Other
◯ Don't know

---

Other - please specify

__________________________________

---

Which school are they in?

◯ School of Arts
◯ School of Humanities
◯ School of Modern Languages
◯ Centre for Academic Language and Development
◯ Centre for Innovation
◯ Don't know

---

Which school are they in?

◯ School of Computer Science, Electrical and Electronic Engineering, and Engineering Mathematics
◯ School of Civil, Aerospace and Mechanical Engineering
◯ Don't know

---

Which school are they in?

◯ Bristol Dental School
◯ Bristol Medical School
◯ Bristol Veterinary School
◯ Centre for Health Sciences Education
◯ Don't know

Which school are they in?

◯ School of Biological Sciences
◯ School of Biochemistry
◯ School of Cellular and Molecular Medicine
◯ School of Physiology, Pharmacology and Neuroscience
◯ School of Psychological Science
◯ Don't know

Which school are they in?

◯ School of Chemistry
◯ School of Earth Sciences
◯ School of Geographical Sciences
◯ School of Mathematics
◯ School of Physics
◯ Don't know

Which school are they in?

◯ School of Education
◯ School for Policy Studies
◯ School of Economics, Finance and Management
◯ School of Sociology, Politics and International Studies
◯ University of Bristol Law School
◯ Don't know

Where were you when you spoke to this person?

◯ Home
◯ Another home
◯ University
◯ Work or volunteering not at university
◯ School or nursery
◯ Shopping
◯ Medical or health centre
◯ Transport
◯ Place of worship
◯ Social (pub/nightclub/café/restaurant)
◯ Exercise
◯ Park
◯ Other location

Other location - please specify

__________________________________

Was this inside or outside?

◯ Inside
◯ Outside
◯ Both inside and outside

How long did you talk to this person for?

◯ Less than 10 minutes
◯ Between 10 minutes and an hour
◯ Between 1 and 4 hours
◯ 4+ hours

Did you also touch this person?

◯ Yes ◯ No

How often would you expect to meet this person under current physical distancing measures?

◯ 4 or more days a week
◯ 2-3 days a week
◯ Once a week
◯ Less often than once a week
◯ Met for the first time this day

Person 3 Description of person:

__________________________________

◯ I did not speak to anyone else in person yesterday on a one-to-one basis.

Age:

◯ 0-4
◯ 5-17
◯ 18-24
◯ 25-44
◯ 45-64
◯ 65-80
◯ 81+

Is this person part of your household?

◯ Yes ◯ No

Does this person work or study at the University of Bristol?

◯ Yes ◯ No ◯ Don't know

What is the main faculty this person is associated with?

◯ Arts
◯ Engineering
◯ Health Sciences
◯ Life Sciences
◯ Science
◯ Social Sciences and Law
◯ Other
◯ Don't know

Which school are they in?

◯ Bristol Dental School
◯ Bristol Medical School
◯ Bristol Veterinary School
◯ Centre for Academic Language and Development
◯ Centre for Health Sciences Education
◯ Centre for Innovation
◯ School for Policy Studies
◯ School of Arts
◯ School of Biochemistry
◯ School of Biological Sciences
◯ School of Cellular and Molecular Medicine
◯ School of Chemistry
◯ School of Civil, Aerospace and Mechanical Engineering
◯ School of Computer Science, Electrical and Electronic Engineering, and Engineering Mathematics
◯ School of Earth Sciences
◯ School of Economics, Finance and Management
◯ School of Education
◯ School of Geographical Sciences
◯ School of Humanities
◯ School of Mathematics
◯ School of Modern Languages
◯ School of Physics
◯ School of Physiology, Pharmacology and Neuroscience
◯ School of Psychological Science
◯ School of Sociology, Politics and International Studies
◯ University of Bristol Law School
◯ Other
◯ Don't know

Other - please specify

__________________________________

Which school are they in?

◯ School of Arts
◯ School of Humanities
◯ School of Modern Languages
◯ Centre for Academic Language and Development
◯ Centre for Innovation
◯ Don't know

Which school are they in?

◯ School of Computer Science, Electrical and Electronic Engineering, and Engineering Mathematics
◯ School of Civil, Aerospace and Mechanical Engineering
◯ Don't know

Which school are they in?

◯ Bristol Dental School
◯ Bristol Medical School
◯ Bristol Veterinary School
◯ Centre for Health Sciences Education
◯ Don't know

Which school are they in?

◯ School of Biological Sciences
◯ School of Biochemistry
◯ School of Cellular and Molecular Medicine
◯ School of Physiology, Pharmacology and Neuroscience
◯ School of Psychological Science
◯ Don't know

Which school are they in?

◯ School of Chemistry
◯ School of Earth Sciences
◯ School of Geographical Sciences
◯ School of Mathematics
◯ School of Physics
◯ Don't know

Which school are they in?

◯ School of Education
◯ School for Policy Studies
◯ School of Economics, Finance and Management
◯ School of Sociology, Politics and International Studies
◯ University of Bristol Law School
◯ Don't know

Where were you when you spoke to this person?

◯ Home
◯ Another home
◯ University
◯ Work or volunteering not at university
◯ School or nursery
◯ Shopping
◯ Medical or health centre
◯ Transport
◯ Place of worship
◯ Social (pub/nightclub/café/restaurant)
◯ Exercise
◯ Park
◯ Other location

Other location - please specify

__________________________________

Was this inside or outside?

◯ Inside
◯ Outside
◯ Both inside and outside

How long did you talk to this person for?

◯ Less than 10 minutes
◯ Between 10 minutes and an hour
◯ Between 1 and 4 hours
◯ 4+ hours

Did you also touch this person?

◯ Yes ◯ No

How often would you expect to meet this person under current physical distancing measures?

◯ 4 or more days a week
◯ 2-3 days a week
◯ Once a week
◯ Less often than once a week
◯ Met for the first time this day

Person 4 Description of person:

__________________________________

◯ I did not speak to anyone else in person yesterday on a one-to-one basis.

Age:

◯ 0-4
◯ 5-17
◯ 18-24
◯ 25-44
◯ 45-64
◯ 65-80
◯ 81+

Is this person part of your household?

◯ Yes ◯ No

Does this person work or study at the University of Bristol?

◯ Yes ◯ No ◯ Don't know

What is the main faculty this person is associated with?

◯ Arts
◯ Engineering
◯ Health Sciences
◯ Life Sciences
◯ Science
◯ Social Sciences and Law
◯ Other
◯ Don't know

Which school are they in?

◯ Bristol Dental School
◯ Bristol Medical School
◯ Bristol Veterinary School
◯ Centre for Academic Language and Development
◯ Centre for Health Sciences Education
◯ Centre for Innovation
◯ School for Policy Studies
◯ School of Arts
◯ School of Biochemistry
◯ School of Biological Sciences
◯ School of Cellular and Molecular Medicine
◯ School of Chemistry
◯ School of Civil, Aerospace and Mechanical Engineering
◯ School of Computer Science, Electrical and Electronic Engineering, and Engineering Mathematics
◯ School of Earth Sciences
◯ School of Economics, Finance and Management
◯ School of Education
◯ School of Geographical Sciences
◯ School of Humanities
◯ School of Mathematics
◯ School of Modern Languages
◯ School of Physics
◯ School of Physiology, Pharmacology and Neuroscience
◯ School of Psychological Science
◯ School of Sociology, Politics and International Studies
◯ University of Bristol Law School
◯ Other
◯ Don't know

Other - please specify

__________________________________

Which school are they in?

◯ School of Arts
◯ School of Humanities
◯ School of Modern Languages
◯ Centre for Academic Language and Development
◯ Centre for Innovation
◯ Don't know

Which school are they in?

◯ School of Computer Science, Electrical and Electronic Engineering, and Engineering Mathematics
◯ School of Civil, Aerospace and Mechanical Engineering
◯ Don't know

Which school are they in?

◯ Bristol Dental School
◯ Bristol Medical School
◯ Bristol Veterinary School
◯ Centre for Health Sciences Education
◯ Don't know

Which school are they in?

◯ School of Biological Sciences
◯ School of Biochemistry
◯ School of Cellular and Molecular Medicine
◯ School of Physiology, Pharmacology and Neuroscience
◯ School of Psychological Science
◯ Don't know

Which school are they in?

◯ School of Chemistry
◯ School of Earth Sciences
◯ School of Geographical Sciences
◯ School of Mathematics
◯ School of Physics
◯ Don't know

Which school are they in?

◯ School of Education
◯ School for Policy Studies
◯ School of Economics, Finance and Management
◯ School of Sociology, Politics and International Studies
◯ University of Bristol Law School
◯ Don't know

Where were you when you spoke to this person?

◯ Home
◯ Another home
◯ University
◯ Work or volunteering not at university
◯ School or nursery
◯ Shopping
◯ Medical or health centre
◯ Transport
◯ Place of worship
◯ Social (pub/nightclub/café/restaurant)
◯ Exercise
◯ Park
◯ Other location

Other location - please specify

__________________________________

Was this inside or outside?

◯ Inside
◯ Outside
◯ Both inside and outside

How long did you talk to this person for?

◯ Less than 10 minutes
◯ Between 10 minutes and an hour
◯ Between 1 and 4 hours
◯ 4+ hours

Did you also touch this person?

◯ Yes ◯ No

How often would you expect to meet this person under current physical distancing measures?

◯ 4 or more days a week
◯ 2-3 days a week
◯ Once a week
◯ Less often than once a week
◯ Met for the first time this day

Person 5 Description of person:

__________________________________

◯ I did not speak to anyone else in person yesterday on a one-to-one basis.

Age:

◯ 0-4
◯ 5-17
◯ 18-24
◯ 25-44
◯ 45-64
◯ 65-80
◯ 81+

Is this person part of your household?

◯ Yes ◯ No

Does this person work or study at the University of Bristol?

◯ Yes ◯ No ◯ Don't know

What is the main faculty this person is associated with?

◯ Arts
◯ Engineering
◯ Health Sciences
◯ Life Sciences
◯ Science
◯ Social Sciences and Law
◯ Other
◯ Don't know

Which school are they in?

◯ Bristol Dental School
◯ Bristol Medical School
◯ Bristol Veterinary School
◯ Centre for Academic Language and Development
◯ Centre for Health Sciences Education
◯ Centre for Innovation
◯ School for Policy Studies
◯ School of Arts
◯ School of Biochemistry
◯ School of Biological Sciences
◯ School of Cellular and Molecular Medicine
◯ School of Chemistry
◯ School of Civil, Aerospace and Mechanical Engineering
◯ School of Computer Science, Electrical and Electronic Engineering, and Engineering Mathematics
◯ School of Earth Sciences
◯ School of Economics, Finance and Management
◯ School of Education
◯ School of Geographical Sciences
◯ School of Humanities
◯ School of Mathematics
◯ School of Modern Languages
◯ School of Physics
◯ School of Physiology, Pharmacology and Neuroscience
◯ School of Psychological Science
◯ School of Sociology, Politics and International Studies
◯ University of Bristol Law School
◯ Other
◯ Don't know

---

Other - please specify

__________________________________

---

Which school are they in?

◯ School of Arts
◯ School of Humanities
◯ School of Modern Languages
◯ Centre for Academic Language and Development
◯ Centre for Innovation
◯ Don't know

---

Which school are they in?

◯ School of Computer Science, Electrical and Electronic Engineering, and Engineering Mathematics
◯ School of Civil, Aerospace and Mechanical Engineering
◯ Don't know

---

Which school are they in?

◯ Bristol Dental School
◯ Bristol Medical School
◯ Bristol Veterinary School
◯ Centre for Health Sciences Education
◯ Don't know

Which school are they in?

◯ School of Biological Sciences
◯ School of Biochemistry
◯ School of Cellular and Molecular Medicine
◯ School of Physiology, Pharmacology and Neuroscience
◯ School of Psychological Science
◯ Don't know

Which school are they in?

◯ School of Chemistry
◯ School of Earth Sciences
◯ School of Geographical Sciences
◯ School of Mathematics
◯ School of Physics
◯ Don't know

Which school are they in?

◯ School of Education
◯ School for Policy Studies
◯ School of Economics, Finance and Management
◯ School of Sociology, Politics and International Studies
◯ University of Bristol Law School
◯ Don't know

Where were you when you spoke to this person?

◯ Home
◯ Another home
◯ University
◯ Work or volunteering not at university
◯ School or nursery
◯ Shopping
◯ Medical or health centre
◯ Transport
◯ Place of worship
◯ Social (pub/nightclub/café/restaurant)
◯ Exercise
◯ Park
◯ Other location

Other location - please specify

__________________________________

Was this inside or outside?

◯ Inside
◯ Outside
◯ Both inside and outside

How long did you talk to this person for?

◯ Less than 10 minutes
◯ Between 10 minutes and an hour
◯ Between 1 and 4 hours
◯ 4+ hours

Did you also touch this person?

◯ Yes ◯ No

How often would you expect to meet this person under current physical distancing measures?

◯ 4 or more days a week
◯ 2-3 days a week
◯ Once a week
◯ Less often than once a week
◯ Met for the first time this day

Person 6 Description of person:

__________________________________

◯ I did not speak to anyone else in person yesterday on a one-to-one basis.

Age:

◯ 0-4
◯ 5-17
◯ 18-24
◯ 25-44
◯ 45-64
◯ 65-80
◯ 81+

Is this person part of your household?

◯ Yes ◯ No

Does this person work or study at the University of Bristol?

◯ Yes ◯ No ◯ Don't know

What is the main faculty this person is associated with?

◯ Arts
◯ Engineering
◯ Health Sciences
◯ Life Sciences
◯ Science
◯ Social Sciences and Law
◯ Other
◯ Don't know

Which school are they in?

◯ Bristol Dental School
◯ Bristol Medical School
◯ Bristol Veterinary School
◯ Centre for Academic Language and Development
◯ Centre for Health Sciences Education
◯ Centre for Innovation
◯ School for Policy Studies
◯ School of Arts
◯ School of Biochemistry
◯ School of Biological Sciences
◯ School of Cellular and Molecular Medicine
◯ School of Chemistry
◯ School of Civil, Aerospace and Mechanical Engineering
◯ School of Computer Science, Electrical and Electronic Engineering, and Engineering Mathematics
◯ School of Earth Sciences
◯ School of Economics, Finance and Management
◯ School of Education
◯ School of Geographical Sciences
◯ School of Humanities
◯ School of Mathematics
◯ School of Modern Languages
◯ School of Physics
◯ School of Physiology, Pharmacology and Neuroscience
◯ School of Psychological Science
◯ School of Sociology, Politics and International Studies
◯ University of Bristol Law School
◯ Other
◯ Don't know

---

Other - please specify

__________________________________

---

Which school are they in?

◯ School of Arts
◯ School of Humanities
◯ School of Modern Languages
◯ Centre for Academic Language and Development
◯ Centre for Innovation
◯ Don't know

---

Which school are they in?

◯ School of Computer Science, Electrical and Electronic Engineering, and Engineering Mathematics
◯ School of Civil, Aerospace and Mechanical Engineering
◯ Don't know

---

Which school are they in?

◯ Bristol Dental School
◯ Bristol Medical School
◯ Bristol Veterinary School
◯ Centre for Health Sciences Education
◯ Don't know

Which school are they in?

◯ School of Biological Sciences
◯ School of Biochemistry
◯ School of Cellular and Molecular Medicine
◯ School of Physiology, Pharmacology and Neuroscience
◯ School of Psychological Science
◯ Don't know

Which school are they in?

◯ School of Chemistry
◯ School of Earth Sciences
◯ School of Geographical Sciences
◯ School of Mathematics
◯ School of Physics
◯ Don't know

Which school are they in?

◯ School of Education
◯ School for Policy Studies
◯ School of Economics, Finance and Management
◯ School of Sociology, Politics and International Studies
◯ University of Bristol Law School
◯ Don't know

Where were you when you spoke to this person?

◯ Home
◯ Another home
◯ University
◯ Work or volunteering not at university
◯ School or nursery
◯ Shopping
◯ Medical or health centre
◯ Transport
◯ Place of worship
◯ Social (pub/nightclub/café/restaurant)
◯ Exercise
◯ Park
◯ Other location

Was this inside or outside?

◯ Inside
◯ Outside
◯ Both inside and outside

Other location - please specify

__________________________________

How long did you talk to this person for?

- ◯ Less than 10 minutes
- ◯ Between 10 minutes and an hour
- ◯ Between 1 and 4 hours
- ◯ 4+ hours

Did you also touch this person?

◯ Yes ◯ No

How often would you expect to meet this person under current physical distancing measures?

- ◯ 4 or more days a week
- ◯ 2-3 days a week
- ◯ Once a week
- ◯ Less often than once a week
- ◯ Met for the first time this day

Person 7 Description of person:

________________________________________

◯ I did not speak to anyone else in person yesterday on a one-to-one basis.

Age:

- ◯ 0-4
- ◯ 5-17
- ◯ 18-24
- ◯ 25-44
- ◯ 45-64
- ◯ 65-80
- ◯ 81+

Is this person part of your household?

◯ Yes ◯ No

Does this person work or study at the University of Bristol?

◯ Yes ◯ No ◯ Don't know

What is the main faculty this person is associated with?

- ◯ Arts
- ◯ Engineering
- ◯ Health Sciences
- ◯ Life Sciences
- ◯ Science
- ◯ Social Sciences and Law
- ◯ Other
- ◯ Don't know

Which school are they in?

◯ Bristol Dental School
◯ Bristol Medical School
◯ Bristol Veterinary School
◯ Centre for Academic Language and Development
◯ Centre for Health Sciences Education
◯ Centre for Innovation
◯ School for Policy Studies
◯ School of Arts
◯ School of Biochemistry
◯ School of Biological Sciences
◯ School of Cellular and Molecular Medicine
◯ School of Chemistry
◯ School of Civil, Aerospace and Mechanical Engineering
◯ School of Computer Science, Electrical and Electronic Engineering, and Engineering Mathematics
◯ School of Earth Sciences
◯ School of Economics, Finance and Management
◯ School of Education
◯ School of Geographical Sciences
◯ School of Humanities
◯ School of Mathematics
◯ School of Modern Languages
◯ School of Physics
◯ School of Physiology, Pharmacology and Neuroscience
◯ School of Psychological Science
◯ School of Sociology, Politics and International Studies
◯ University of Bristol Law School
◯ Other
◯ Don't know

---

Other - please specify

__________________________________

---

Which school are they in?

◯ School of Arts
◯ School of Humanities
◯ School of Modern Languages
◯ Centre for Academic Language and Development
◯ Centre for Innovation
◯ Don't know

---

Which school are they in?

◯ School of Computer Science, Electrical and Electronic Engineering, and Engineering Mathematics
◯ School of Civil, Aerospace and Mechanical Engineering
◯ Don't know

---

Which school are they in?

◯ Bristol Dental School
◯ Bristol Medical School
◯ Bristol Veterinary School
◯ Centre for Health Sciences Education
◯ Don't know

Which school are they in?

◯ School of Biological Sciences
◯ School of Biochemistry
◯ School of Cellular and Molecular Medicine
◯ School of Physiology, Pharmacology and Neuroscience
◯ School of Psychological Science
◯ Don't know

Which school are they in?

◯ School of Chemistry
◯ School of Earth Sciences
◯ School of Geographical Sciences
◯ School of Mathematics
◯ School of Physics
◯ Don't know

Which school are they in?

◯ School of Education
◯ School for Policy Studies
◯ School of Economics, Finance and Management
◯ School of Sociology, Politics and International Studies
◯ University of Bristol Law School
◯ Don't know

Where were you when you spoke to this person?

◯ Home
◯ Another home
◯ University
◯ Work or volunteering not at university
◯ School or nursery
◯ Shopping
◯ Medical or health centre
◯ Transport
◯ Place of worship
◯ Social (pub/nightclub/café/restaurant)
◯ Exercise
◯ Park
◯ Other location

Other location - please specify

__________________________________

Was this inside or outside?

◯ Inside
◯ Outside
◯ Both inside and outside

How long did you talk to this person for?

◯ Less than 10 minutes
◯ Between 10 minutes and an hour
◯ Between 1 and 4 hours
◯ 4+ hours

Did you also touch this person?

◯ Yes ◯ No

How often would you expect to meet this person under current physical distancing measures?

◯ 4 or more days a week
◯ 2-3 days a week
◯ Once a week
◯ Less often than once a week
◯ Met for the first time this day

Person 8 Description of person:

__________________________________

◯ I did not speak to anyone else in person yesterday on a one-to-one basis.

Age:

◯ 0-4
◯ 5-17
◯ 18-24
◯ 25-44
◯ 45-64
◯ 65-80
◯ 81+

Is this person part of your household?

◯ Yes ◯ No

Does this person work or study at the University of Bristol?

◯ Yes ◯ No ◯ Don't know

What is the main faculty this person is associated with?

◯ Arts
◯ Engineering
◯ Health Sciences
◯ Life Sciences
◯ Science
◯ Social Sciences and Law
◯ Other
◯ Don't know

Which school are they in?

◯ Bristol Dental School
◯ Bristol Medical School
◯ Bristol Veterinary School
◯ Centre for Academic Language and Development
◯ Centre for Health Sciences Education
◯ Centre for Innovation
◯ School for Policy Studies
◯ School of Arts
◯ School of Biochemistry
◯ School of Biological Sciences
◯ School of Cellular and Molecular Medicine
◯ School of Chemistry
◯ School of Civil, Aerospace and Mechanical Engineering
◯ School of Computer Science, Electrical and Electronic Engineering, and Engineering Mathematics
◯ School of Earth Sciences
◯ School of Economics, Finance and Management
◯ School of Education
◯ School of Geographical Sciences
◯ School of Humanities
◯ School of Mathematics
◯ School of Modern Languages
◯ School of Physics
◯ School of Physiology, Pharmacology and Neuroscience
◯ School of Psychological Science
◯ School of Sociology, Politics and International Studies
◯ University of Bristol Law School
◯ Other
◯ Don't know

Other - please specify

__________________________________

Which school are they in?

◯ School of Arts
◯ School of Humanities
◯ School of Modern Languages
◯ Centre for Academic Language and Development
◯ Centre for Innovation
◯ Don't know

Which school are they in?

◯ School of Computer Science, Electrical and Electronic Engineering, and Engineering Mathematics
◯ School of Civil, Aerospace and Mechanical Engineering
◯ Don't know

Which school are they in?

◯ Bristol Dental School
◯ Bristol Medical School
◯ Bristol Veterinary School
◯ Centre for Health Sciences Education
◯ Don't know

Which school are they in?

◯ School of Biological Sciences
◯ School of Biochemistry
◯ School of Cellular and Molecular Medicine
◯ School of Physiology, Pharmacology and Neuroscience
◯ School of Psychological Science
◯ Don't know

Which school are they in?

◯ School of Chemistry
◯ School of Earth Sciences
◯ School of Geographical Sciences
◯ School of Mathematics
◯ School of Physics
◯ Don't know

Which school are they in?

◯ School of Education
◯ School for Policy Studies
◯ School of Economics, Finance and Management
◯ School of Sociology, Politics and International Studies
◯ University of Bristol Law School
◯ Don't know

Where were you when you spoke to this person?

◯ Home
◯ Another home
◯ University
◯ Work or volunteering not at university
◯ School or nursery
◯ Shopping
◯ Medical or health centre
◯ Transport
◯ Place of worship
◯ Social (pub/nightclub/café/restaurant)
◯ Exercise
◯ Park
◯ Other location

Other location - please specify

__________________________________

Was this inside or outside?

◯ Inside
◯ Outside
◯ Both inside and outside

How long did you talk to this person for?

◯ Less than 10 minutes
◯ Between 10 minutes and an hour
◯ Between 1 and 4 hours
◯ 4+ hours

Did you also touch this person?

◯ Yes ◯ No

How often would you expect to meet this person under current physical distancing measures?

◯ 4 or more days a week
◯ 2-3 days a week
◯ Once a week
◯ Less often than once a week
◯ Met for the first time this day

Person 9 Description of person:

__________________________________

◯ I did not speak to anyone else in person yesterday on a one-to-one basis.

Age:

◯ 0-4
◯ 5-17
◯ 18-24
◯ 25-44
◯ 45-64
◯ 65-80
◯ 81+

Is this person part of your household?

◯ Yes ◯ No

Does this person work or study at the University of Bristol?

◯ Yes ◯ No ◯ Don't know

What is the main faculty this person is associated with?

◯ Arts
◯ Engineering
◯ Health Sciences
◯ Life Sciences
◯ Science
◯ Social Sciences and Law
◯ Other
◯ Don't know

Which school are they in?

◯ Bristol Dental School
◯ Bristol Medical School
◯ Bristol Veterinary School
◯ Centre for Academic Language and Development
◯ Centre for Health Sciences Education
◯ Centre for Innovation
◯ School for Policy Studies
◯ School of Arts
◯ School of Biochemistry
◯ School of Biological Sciences
◯ School of Cellular and Molecular Medicine
◯ School of Chemistry
◯ School of Civil, Aerospace and Mechanical Engineering
◯ School of Computer Science, Electrical and Electronic Engineering, and Engineering Mathematics
◯ School of Earth Sciences
◯ School of Economics, Finance and Management
◯ School of Education
◯ School of Geographical Sciences
◯ School of Humanities
◯ School of Mathematics
◯ School of Modern Languages
◯ School of Physics
◯ School of Physiology, Pharmacology and Neuroscience
◯ School of Psychological Science
◯ School of Sociology, Politics and International Studies
◯ University of Bristol Law School
◯ Other
◯ Don't know

---

Other - please specify

__________________________________

---

Which school are they in?

◯ School of Arts
◯ School of Humanities
◯ School of Modern Languages
◯ Centre for Academic Language and Development
◯ Centre for Innovation
◯ Don't know

---

Which school are they in?

◯ School of Computer Science, Electrical and Electronic Engineering, and Engineering Mathematics
◯ School of Civil, Aerospace and Mechanical Engineering
◯ Don't know

---

Which school are they in?

◯ Bristol Dental School
◯ Bristol Medical School
◯ Bristol Veterinary School
◯ Centre for Health Sciences Education
◯ Don't know

Which school are they in?

◯ School of Biological Sciences
◯ School of Biochemistry
◯ School of Cellular and Molecular Medicine
◯ School of Physiology, Pharmacology and Neuroscience
◯ School of Psychological Science
◯ Don't know

Which school are they in?

◯ School of Chemistry
◯ School of Earth Sciences
◯ School of Geographical Sciences
◯ School of Mathematics
◯ School of Physics
◯ Don't know

Which school are they in?

◯ School of Education
◯ School for Policy Studies
◯ School of Economics, Finance and Management
◯ School of Sociology, Politics and International Studies
◯ University of Bristol Law School
◯ Don't know

Where were you when you spoke to this person?

◯ Home
◯ Another home
◯ University
◯ Work or volunteering not at university
◯ School or nursery
◯ Shopping
◯ Medical or health centre
◯ Transport
◯ Place of worship
◯ Social (pub/nightclub/café/restaurant)
◯ Exercise
◯ Park
◯ Other location

Other location - please specify

__________________________________

Was this inside or outside?

◯ Inside
◯ Outside
◯ Both inside and outside

How long did you talk to this person for?

◯ Less than 10 minutes
◯ Between 10 minutes and an hour
◯ Between 1 and 4 hours
◯ 4+ hours

Did you also touch this person?

◯ Yes ◯ No

How often would you expect to meet this person under current physical distancing measures?

◯ 4 or more days a week
◯ 2-3 days a week
◯ Once a week
◯ Less often than once a week
◯ Met for the first time this day

Person 10 Description of person:

__________________________________

◯ I did not speak to anyone else in person yesterday on a one-to-one basis.

Age:

◯ 0-4
◯ 5-17
◯ 18-24
◯ 25-44
◯ 45-64
◯ 65-80
◯ 81+

Is this person part of your household?

◯ Yes ◯ No

Does this person work or study at the University of Bristol?

◯ Yes ◯ No ◯ Don't know

What is the main faculty this person is associated with?

◯ Arts
◯ Engineering
◯ Health Sciences
◯ Life Sciences
◯ Science
◯ Social Sciences and Law
◯ Other
◯ Don't know

Which school are they in?

◯ Bristol Dental School
◯ Bristol Medical School
◯ Bristol Veterinary School
◯ Centre for Academic Language and Development
◯ Centre for Health Sciences Education
◯ Centre for Innovation
◯ School for Policy Studies
◯ School of Arts
◯ School of Biochemistry
◯ School of Biological Sciences
◯ School of Cellular and Molecular Medicine
◯ School of Chemistry
◯ School of Civil, Aerospace and Mechanical Engineering
◯ School of Computer Science, Electrical and Electronic Engineering, and Engineering Mathematics
◯ School of Earth Sciences
◯ School of Economics, Finance and Management
◯ School of Education
◯ School of Geographical Sciences
◯ School of Humanities
◯ School of Mathematics
◯ School of Modern Languages
◯ School of Physics
◯ School of Physiology, Pharmacology and Neuroscience
◯ School of Psychological Science
◯ School of Sociology, Politics and International Studies
◯ University of Bristol Law School
◯ Other
◯ Don't know

---

Other - please specify

__________________________________

---

Which school are they in?

◯ School of Arts
◯ School of Humanities
◯ School of Modern Languages
◯ Centre for Academic Language and Development
◯ Centre for Innovation
◯ Don't know

---

Which school are they in?

◯ School of Computer Science, Electrical and Electronic Engineering, and Engineering Mathematics
◯ School of Civil, Aerospace and Mechanical Engineering
◯ Don't know

---

Which school are they in?

◯ Bristol Dental School
◯ Bristol Medical School
◯ Bristol Veterinary School
◯ Centre for Health Sciences Education
◯ Don't know

Which school are they in?

◯ School of Biological Sciences
◯ School of Biochemistry
◯ School of Cellular and Molecular Medicine
◯ School of Physiology, Pharmacology and Neuroscience
◯ School of Psychological Science
◯ Don't know

Which school are they in?

◯ School of Chemistry
◯ School of Earth Sciences
◯ School of Geographical Sciences
◯ School of Mathematics
◯ School of Physics
◯ Don't know

Which school are they in?

◯ School of Education
◯ School for Policy Studies
◯ School of Economics, Finance and Management
◯ School of Sociology, Politics and International Studies
◯ University of Bristol Law School
◯ Don't know

Where were you when you spoke to this person?

◯ Home
◯ Another home
◯ University
◯ Work or volunteering not at university
◯ School or nursery
◯ Shopping
◯ Medical or health centre
◯ Transport
◯ Place of worship
◯ Social (pub/nightclub/café/restaurant)
◯ Exercise
◯ Park
◯ Other location

Other location - please specify

__________________________________

Was this inside or outside?

◯ Inside
◯ Outside
◯ Both inside and outside

How long did you talk to this person for?

◯ Less than 10 minutes
◯ Between 10 minutes and an hour
◯ Between 1 and 4 hours
◯ 4+ hours

Did you also touch this person?

◯ Yes ◯ No

How often would you expect to meet this person under current physical distancing measures?

◯ 4 or more days a week
◯ 2-3 days a week
◯ Once a week
◯ Less often than once a week
◯ Met for the first time this day

Person 11 Description of person:

__________________________________

◯ I did not speak to anyone else in person yesterday on a one-to-one basis.

Age:

◯ 0-4
◯ 5-17
◯ 18-24
◯ 25-44
◯ 45-64
◯ 65-80
◯ 81+

Is this person part of your household?

◯ Yes ◯ No

Does this person work or study at the University of Bristol?

◯ Yes ◯ No ◯ Don't know

What is the main faculty this person is associated with?

◯ Arts
◯ Engineering
◯ Health Sciences
◯ Life Sciences
◯ Science
◯ Social Sciences and Law
◯ Other
◯ Don't know

Which school are they in?

◯ Bristol Dental School
◯ Bristol Medical School
◯ Bristol Veterinary School
◯ Centre for Academic Language and Development
◯ Centre for Health Sciences Education
◯ Centre for Innovation
◯ School for Policy Studies
◯ School of Arts
◯ School of Biochemistry
◯ School of Biological Sciences
◯ School of Cellular and Molecular Medicine
◯ School of Chemistry
◯ School of Civil, Aerospace and Mechanical Engineering
◯ School of Computer Science, Electrical and Electronic Engineering, and Engineering Mathematics
◯ School of Earth Sciences
◯ School of Economics, Finance and Management
◯ School of Education
◯ School of Geographical Sciences
◯ School of Humanities
◯ School of Mathematics
◯ School of Modern Languages
◯ School of Physics
◯ School of Physiology, Pharmacology and Neuroscience
◯ School of Psychological Science
◯ School of Sociology, Politics and International Studies
◯ University of Bristol Law School
◯ Other
◯ Don't know

---

Other - please specify

__________________________________

---

Which school are they in?

◯ School of Arts
◯ School of Humanities
◯ School of Modern Languages
◯ Centre for Academic Language and Development
◯ Centre for Innovation
◯ Don't know

---

Which school are they in?

◯ School of Computer Science, Electrical and Electronic Engineering, and Engineering Mathematics
◯ School of Civil, Aerospace and Mechanical Engineering
◯ Don't know

---

Which school are they in?

◯ Bristol Dental School
◯ Bristol Medical School
◯ Bristol Veterinary School
◯ Centre for Health Sciences Education
◯ Don't know

Which school are they in?

◯ School of Biological Sciences
◯ School of Biochemistry
◯ School of Cellular and Molecular Medicine
◯ School of Physiology, Pharmacology and Neuroscience
◯ School of Psychological Science
◯ Don't know

Which school are they in?

◯ School of Chemistry
◯ School of Earth Sciences
◯ School of Geographical Sciences
◯ School of Mathematics
◯ School of Physics
◯ Don't know

Which school are they in?

◯ School of Education
◯ School for Policy Studies
◯ School of Economics, Finance and Management
◯ School of Sociology, Politics and International Studies
◯ University of Bristol Law School
◯ Don't know

Where were you when you spoke to this person?

◯ Home
◯ Another home
◯ University
◯ Work or volunteering not at university
◯ School or nursery
◯ Shopping
◯ Medical or health centre
◯ Transport
◯ Place of worship
◯ Social (pub/nightclub/café/restaurant)
◯ Exercise
◯ Park
◯ Other location

Other location - please specify

__________________________________

Was this inside or outside?

◯ Inside
◯ Outside
◯ Both inside and outside

How long did you talk to this person for?

◯ Less than 10 minutes
◯ Between 10 minutes and an hour
◯ Between 1 and 4 hours
◯ 4+ hours

Did you also touch this person?

◯ Yes ◯ No

How often would you expect to meet this person under current physical distancing measures?

◯ 4 or more days a week
◯ 2-3 days a week
◯ Once a week
◯ Less often than once a week
◯ Met for the first time this day

Person 12 Description of person:

__________________________________

◯ I did not speak to anyone else in person yesterday on a one-to-one basis.

Age:

◯ 0-4
◯ 5-17
◯ 18-24
◯ 25-44
◯ 45-64
◯ 65-80
◯ 81+

Is this person part of your household?

◯ Yes ◯ No

Does this person work or study at the University of Bristol?

◯ Yes ◯ No ◯ Don't know

What is the main faculty this person is associated with?

◯ Arts
◯ Engineering
◯ Health Sciences
◯ Life Sciences
◯ Science
◯ Social Sciences and Law
◯ Other
◯ Don't know

Which school are they in?

◯ Bristol Dental School
◯ Bristol Medical School
◯ Bristol Veterinary School
◯ Centre for Academic Language and Development
◯ Centre for Health Sciences Education
◯ Centre for Innovation
◯ School for Policy Studies
◯ School of Arts
◯ School of Biochemistry
◯ School of Biological Sciences
◯ School of Cellular and Molecular Medicine
◯ School of Chemistry
◯ School of Civil, Aerospace and Mechanical Engineering
◯ School of Computer Science, Electrical and Electronic Engineering, and Engineering Mathematics
◯ School of Earth Sciences
◯ School of Economics, Finance and Management
◯ School of Education
◯ School of Geographical Sciences
◯ School of Humanities
◯ School of Mathematics
◯ School of Modern Languages
◯ School of Physics
◯ School of Physiology, Pharmacology and Neuroscience
◯ School of Psychological Science
◯ School of Sociology, Politics and International Studies
◯ University of Bristol Law School
◯ Other
◯ Don't know

---

Other - please specify

__________________________________

---

Which school are they in?

◯ School of Arts
◯ School of Humanities
◯ School of Modern Languages
◯ Centre for Academic Language and Development
◯ Centre for Innovation
◯ Don't know

---

Which school are they in?

◯ School of Computer Science, Electrical and Electronic Engineering, and Engineering Mathematics
◯ School of Civil, Aerospace and Mechanical Engineering
◯ Don't know

---

Which school are they in?

◯ Bristol Dental School
◯ Bristol Medical School
◯ Bristol Veterinary School
◯ Centre for Health Sciences Education
◯ Don't know

Which school are they in?

◯ School of Biological Sciences
◯ School of Biochemistry
◯ School of Cellular and Molecular Medicine
◯ School of Physiology, Pharmacology and Neuroscience
◯ School of Psychological Science
◯ Don't know

Which school are they in?

◯ School of Chemistry
◯ School of Earth Sciences
◯ School of Geographical Sciences
◯ School of Mathematics
◯ School of Physics
◯ Don't know

Which school are they in?

◯ School of Education
◯ School for Policy Studies
◯ School of Economics, Finance and Management
◯ School of Sociology, Politics and International Studies
◯ University of Bristol Law School
◯ Don't know

Where were you when you spoke to this person?

◯ Home
◯ Another home
◯ University
◯ Work or volunteering not at university
◯ School or nursery
◯ Shopping
◯ Medical or health centre
◯ Transport
◯ Place of worship
◯ Social (pub/nightclub/café/restaurant)
◯ Exercise
◯ Park
◯ Other location

Other location - please specify

__________________________________

Was this inside or outside?

◯ Inside
◯ Outside
◯ Both inside and outside

How long did you talk to this person for?

◯ Less than 10 minutes
◯ Between 10 minutes and an hour
◯ Between 1 and 4 hours
◯ 4+ hours

Did you also touch this person?

◯ Yes ◯ No

How often would you expect to meet this person under current physical distancing measures?

◯ 4 or more days a week
◯ 2-3 days a week
◯ Once a week
◯ Less often than once a week
◯ Met for the first time this day

Person 13 Description of person:

__________________________________

◯ I did not speak to anyone else in person yesterday on a one-to-one basis.

Age:

◯ 0-4
◯ 5-17
◯ 18-24
◯ 25-44
◯ 45-64
◯ 65-80
◯ 81+

Is this person part of your household?

◯ Yes ◯ No

Does this person work or study at the University of Bristol?

◯ Yes ◯ No ◯ Don't know

What is the main faculty this person is associated with?

◯ Arts
◯ Engineering
◯ Health Sciences
◯ Life Sciences
◯ Science
◯ Social Sciences and Law
◯ Other
◯ Don't know

Which school are they in?

◯ Bristol Dental School
◯ Bristol Medical School
◯ Bristol Veterinary School
◯ Centre for Academic Language and Development
◯ Centre for Health Sciences Education
◯ Centre for Innovation
◯ School for Policy Studies
◯ School of Arts
◯ School of Biochemistry
◯ School of Biological Sciences
◯ School of Cellular and Molecular Medicine
◯ School of Chemistry
◯ School of Civil, Aerospace and Mechanical Engineering
◯ School of Computer Science, Electrical and Electronic Engineering, and Engineering Mathematics
◯ School of Earth Sciences
◯ School of Economics, Finance and Management
◯ School of Education
◯ School of Geographical Sciences
◯ School of Humanities
◯ School of Mathematics
◯ School of Modern Languages
◯ School of Physics
◯ School of Physiology, Pharmacology and Neuroscience
◯ School of Psychological Science
◯ School of Sociology, Politics and International Studies
◯ University of Bristol Law School
◯ Other
◯ Don't know

Other - please specify

__________________________________

Which school are they in?

◯ School of Arts
◯ School of Humanities
◯ School of Modern Languages
◯ Centre for Academic Language and Development
◯ Centre for Innovation
◯ Don't know

Which school are they in?

◯ School of Computer Science, Electrical and Electronic Engineering, and Engineering Mathematics
◯ School of Civil, Aerospace and Mechanical Engineering
◯ Don't know

Which school are they in?

◯ Bristol Dental School
◯ Bristol Medical School
◯ Bristol Veterinary School
◯ Centre for Health Sciences Education
◯ Don't know

Which school are they in?

◯ School of Biological Sciences
◯ School of Biochemistry
◯ School of Cellular and Molecular Medicine
◯ School of Physiology, Pharmacology and Neuroscience
◯ School of Psychological Science
◯ Don't know

Which school are they in?

◯ School of Chemistry
◯ School of Earth Sciences
◯ School of Geographical Sciences
◯ School of Mathematics
◯ School of Physics
◯ Don't know

Which school are they in?

◯ School of Education
◯ School for Policy Studies
◯ School of Economics, Finance and Management
◯ School of Sociology, Politics and International Studies
◯ University of Bristol Law School
◯ Don't know

Where were you when you spoke to this person?

◯ Home
◯ Another home
◯ University
◯ Work or volunteering not at university
◯ School or nursery
◯ Shopping
◯ Medical or health centre
◯ Transport
◯ Place of worship
◯ Social (pub/nightclub/café/restaurant)
◯ Exercise
◯ Park
◯ Other location

Other location - please specify

__________________________________

Was this inside or outside?

◯ Inside
◯ Outside
◯ Both inside and outside

How long did you talk to this person for?

◯ Less than 10 minutes
◯ Between 10 minutes and an hour
◯ Between 1 and 4 hours
◯ 4+ hours

Did you also touch this person?

◯ Yes ◯ No

How often would you expect to meet this person under current physical distancing measures?

◯ 4 or more days a week
◯ 2-3 days a week
◯ Once a week
◯ Less often than once a week
◯ Met for the first time this day

Person 14 Description of person:

__________________________________

◯ I did not speak to anyone else in person yesterday on a one-to-one basis.

Age:

◯ 0-4
◯ 5-17
◯ 18-24
◯ 25-44
◯ 45-64
◯ 65-80
◯ 81+

Is this person part of your household?

◯ Yes ◯ No

Does this person work or study at the University of Bristol?

◯ Yes ◯ No ◯ Don't know

What is the main faculty this person is associated with?

◯ Arts
◯ Engineering
◯ Health Sciences
◯ Life Sciences
◯ Science
◯ Social Sciences and Law
◯ Other
◯ Don't know

Which school are they in?

◯ Bristol Dental School
◯ Bristol Medical School
◯ Bristol Veterinary School
◯ Centre for Academic Language and Development
◯ Centre for Health Sciences Education
◯ Centre for Innovation
◯ School for Policy Studies
◯ School of Arts
◯ School of Biochemistry
◯ School of Biological Sciences
◯ School of Cellular and Molecular Medicine
◯ School of Chemistry
◯ School of Civil, Aerospace and Mechanical Engineering
◯ School of Computer Science, Electrical and Electronic Engineering, and Engineering Mathematics
◯ School of Earth Sciences
◯ School of Economics, Finance and Management
◯ School of Education
◯ School of Geographical Sciences
◯ School of Humanities
◯ School of Mathematics
◯ School of Modern Languages
◯ School of Physics
◯ School of Physiology, Pharmacology and Neuroscience
◯ School of Psychological Science
◯ School of Sociology, Politics and International Studies
◯ University of Bristol Law School
◯ Other
◯ Don't know

Other - please specify

__________________________________

Which school are they in?

◯ School of Arts
◯ School of Humanities
◯ School of Modern Languages
◯ Centre for Academic Language and Development
◯ Centre for Innovation
◯ Don't know

Which school are they in?

◯ School of Computer Science, Electrical and Electronic Engineering, and Engineering Mathematics
◯ School of Civil, Aerospace and Mechanical Engineering
◯ Don't know

Which school are they in?

◯ Bristol Dental School
◯ Bristol Medical School
◯ Bristol Veterinary School
◯ Centre for Health Sciences Education
◯ Don't know

Which school are they in?

◯ School of Biological Sciences
◯ School of Biochemistry
◯ School of Cellular and Molecular Medicine
◯ School of Physiology, Pharmacology and Neuroscience
◯ School of Psychological Science
◯ Don't know

Which school are they in?

◯ School of Chemistry
◯ School of Earth Sciences
◯ School of Geographical Sciences
◯ School of Mathematics
◯ School of Physics
◯ Don't know

Which school are they in?

◯ School of Education
◯ School for Policy Studies
◯ School of Economics, Finance and Management
◯ School of Sociology, Politics and International Studies
◯ University of Bristol Law School
◯ Don't know

Where were you when you spoke to this person?

◯ Home
◯ Another home
◯ University
◯ Work or volunteering not at university
◯ School or nursery
◯ Shopping
◯ Medical or health centre
◯ Transport
◯ Place of worship
◯ Social (pub/nightclub/café/restaurant)
◯ Exercise
◯ Park
◯ Other location

Other location - please specify

__________________________________

Was this inside or outside?

◯ Inside
◯ Outside

How long did you talk to this person for?

◯ Less than 10 minutes
◯ Between 10 minutes and an hour
◯ Between 1 and 4 hours
◯ 4+ hours

Did you also touch this person?

◯ Yes ◯ No

How often would you expect to meet this person under current physical distancing measures?

◯ 4 or more days a week
◯ 2-3 days a week
◯ Once a week
◯ Less often than once a week
◯ Met for the first time this day

Person 15 Description of person:

________________________________

◯ I did not speak to anyone else in person yesterday on a one-to-one basis.

Age:

◯ 0-4
◯ 5-17
◯ 18-24
◯ 25-44
◯ 45-64
◯ 65-80
◯ 81+

Is this person part of your household?

◯ Yes ◯ No

What is the main faculty this person is associated with?

◯ Arts
◯ Engineering
◯ Health Sciences
◯ Life Sciences
◯ Science
◯ Social Sciences and Law
◯ Other
◯ Don't know

Does this person work or study at the University of Bristol?

◯ Yes ◯ No ◯ Don't know

Which school are they in?

◯ Bristol Dental School
◯ Bristol Medical School
◯ Bristol Veterinary School
◯ Centre for Academic Language and Development
◯ Centre for Health Sciences Education
◯ Centre for Innovation
◯ School for Policy Studies
◯ School of Arts
◯ School of Biochemistry
◯ School of Biological Sciences
◯ School of Cellular and Molecular Medicine
◯ School of Chemistry
◯ School of Civil, Aerospace and Mechanical Engineering
◯ School of Computer Science, Electrical and Electronic Engineering, and Engineering Mathematics
◯ School of Earth Sciences
◯ School of Economics, Finance and Management
◯ School of Education
◯ School of Geographical Sciences
◯ School of Humanities
◯ School of Mathematics
◯ School of Modern Languages
◯ School of Physics
◯ School of Physiology, Pharmacology and Neuroscience
◯ School of Psychological Science
◯ School of Sociology, Politics and International Studies
◯ University of Bristol Law School
◯ Other
◯ Don't know

---

Other - please specify

__________________________________

---

Which school are they in?

◯ School of Arts
◯ School of Humanities
◯ School of Modern Languages
◯ Centre for Academic Language and Development
◯ Centre for Innovation
◯ Don't know

---

Which school are they in?

◯ School of Computer Science, Electrical and Electronic Engineering, and Engineering Mathematics
◯ School of Civil, Aerospace and Mechanical Engineering
◯ Don't know

---

Which school are they in?

◯ Bristol Dental School
◯ Bristol Medical School
◯ Bristol Veterinary School
◯ Centre for Health Sciences Education
◯ Don't know

Which school are they in?

◯ School of Biological Sciences
◯ School of Biochemistry
◯ School of Cellular and Molecular Medicine
◯ School of Physiology, Pharmacology and Neuroscience
◯ School of Psychological Science
◯ Don't know

Which school are they in?

◯ School of Chemistry
◯ School of Earth Sciences
◯ School of Geographical Sciences
◯ School of Mathematics
◯ School of Physics
◯ Don't know

Which school are they in?

◯ School of Education
◯ School for Policy Studies
◯ School of Economics, Finance and Management
◯ School of Sociology, Politics and International Studies
◯ University of Bristol Law School
◯ Don't know

Where were you when you spoke to this person?

◯ Home
◯ Another home
◯ University
◯ Work or volunteering not at university
◯ School or nursery
◯ Shopping
◯ Medical or health centre
◯ Transport
◯ Place of worship
◯ Social (pub/nightclub/café/restaurant)
◯ Exercise
◯ Park
◯ Other location

Other location - please specify

__________________________________

Was this inside or outside?

◯ Inside
◯ Outside

How long did you talk to this person for?

◯ Less than 10 minutes
◯ Between 10 minutes and an hour
◯ Between 1 and 4 hours
◯ 4+ hours

Did you also touch this person?

◯ Yes ◯ No

How often would you expect to meet this person under current physical distancing measures?

◯ 4 or more days a week
◯ 2-3 days a week
◯ Once a week
◯ Less often than once a week
◯ Met for the first time this day

Person 16 Description of person:

__________________________________

◯ I did not speak to anyone else in person yesterday on a one-to-one basis.

Age:

◯ 0-4
◯ 5-17
◯ 18-24
◯ 25-44
◯ 45-64
◯ 65-80
◯ 81+

Is this person part of your household?

◯ Yes ◯ No

Does this person work or study at the University of Bristol?

◯ Yes ◯ No ◯ Don't know

What is the main faculty this person is associated with?

◯ Arts
◯ Engineering
◯ Health Sciences
◯ Life Sciences
◯ Science
◯ Social Sciences and Law
◯ Other
◯ Don't know

Which school are they in?

◯ Bristol Dental School
◯ Bristol Medical School
◯ Bristol Veterinary School
◯ Centre for Academic Language and Development
◯ Centre for Health Sciences Education
◯ Centre for Innovation
◯ School for Policy Studies
◯ School of Arts
◯ School of Biochemistry
◯ School of Biological Sciences
◯ School of Cellular and Molecular Medicine
◯ School of Chemistry
◯ School of Civil, Aerospace and Mechanical Engineering
◯ School of Computer Science, Electrical and Electronic Engineering, and Engineering Mathematics
◯ School of Earth Sciences
◯ School of Economics, Finance and Management
◯ School of Education
◯ School of Geographical Sciences
◯ School of Humanities
◯ School of Mathematics
◯ School of Modern Languages
◯ School of Physics
◯ School of Physiology, Pharmacology and Neuroscience
◯ School of Psychological Science
◯ School of Sociology, Politics and International Studies
◯ University of Bristol Law School
◯ Other
◯ Don't know

---

Other - please specify

__________________________________

---

Which school are they in?

◯ School of Arts
◯ School of Humanities
◯ School of Modern Languages
◯ Centre for Academic Language and Development
◯ Centre for Innovation
◯ Don't know

---

Which school are they in?

◯ School of Computer Science, Electrical and Electronic Engineering, and Engineering Mathematics
◯ School of Civil, Aerospace and Mechanical Engineering
◯ Don't know

---

Which school are they in?

◯ Bristol Dental School
◯ Bristol Medical School
◯ Bristol Veterinary School
◯ Centre for Health Sciences Education
◯ Don't know

Which school are they in?

◯ School of Biological Sciences
◯ School of Biochemistry
◯ School of Cellular and Molecular Medicine
◯ School of Physiology, Pharmacology and Neuroscience
◯ School of Psychological Science
◯ Don't know

Which school are they in?

◯ School of Chemistry
◯ School of Earth Sciences
◯ School of Geographical Sciences
◯ School of Mathematics
◯ School of Physics
◯ Don't know

Which school are they in?

◯ School of Education
◯ School for Policy Studies
◯ School of Economics, Finance and Management
◯ School of Sociology, Politics and International Studies
◯ University of Bristol Law School
◯ Don't know

Where were you when you spoke to this person?

◯ Home
◯ Another home
◯ University
◯ Work or volunteering not at university
◯ School or nursery
◯ Shopping
◯ Medical or health centre
◯ Transport
◯ Place of worship
◯ Social (pub/nightclub/café/restaurant)
◯ Exercise
◯ Park
◯ Other location

Other location - please specify

__________________________________

Was this inside or outside?

◯ Inside
◯ Outside

How long did you talk to this person for?

◯ Less than 10 minutes
◯ Between 10 minutes and an hour
◯ Between 1 and 4 hours
◯ 4+ hours

Did you also touch this person?

◯ Yes ◯ No

How often would you expect to meet this person under current physical distancing measures?

◯ 4 or more days a week
◯ 2-3 days a week
◯ Once a week
◯ Less often than once a week
◯ Met for the first time this day

Person 17 Description of person:

__________________________________

◯ I did not speak to anyone else in person yesterday on a one-to-one basis.

Age:

◯ 0-4
◯ 5-17
◯ 18-24
◯ 25-44
◯ 45-64
◯ 65-80
◯ 81+

Is this person part of your household?

◯ Yes ◯ No

Does this person work or study at the University of Bristol?

◯ Yes ◯ No ◯ Don't know

What is the main faculty this person is associated with?

◯ Arts
◯ Engineering
◯ Health Sciences
◯ Life Sciences
◯ Science
◯ Social Sciences and Law
◯ Other
◯ Don't know

Which school are they in?

◯ Bristol Dental School
◯ Bristol Medical School
◯ Bristol Veterinary School
◯ Centre for Academic Language and Development
◯ Centre for Health Sciences Education
◯ Centre for Innovation
◯ School for Policy Studies
◯ School of Arts
◯ School of Biochemistry
◯ School of Biological Sciences
◯ School of Cellular and Molecular Medicine
◯ School of Chemistry
◯ School of Civil, Aerospace and Mechanical Engineering
◯ School of Computer Science, Electrical and Electronic Engineering, and Engineering Mathematics
◯ School of Earth Sciences
◯ School of Economics, Finance and Management
◯ School of Education
◯ School of Geographical Sciences
◯ School of Humanities
◯ School of Mathematics
◯ School of Modern Languages
◯ School of Physics
◯ School of Physiology, Pharmacology and Neuroscience
◯ School of Psychological Science
◯ School of Sociology, Politics and International Studies
◯ University of Bristol Law School
◯ Other
◯ Don't know

---

Other - please specify

__________________________________

---

Which school are they in?

◯ School of Arts
◯ School of Humanities
◯ School of Modern Languages
◯ Centre for Academic Language and Development
◯ Centre for Innovation
◯ Don't know

---

Which school are they in?

◯ School of Computer Science, Electrical and Electronic Engineering, and Engineering Mathematics
◯ School of Civil, Aerospace and Mechanical Engineering
◯ Don't know

---

Which school are they in?

◯ Bristol Dental School
◯ Bristol Medical School
◯ Bristol Veterinary School
◯ Centre for Health Sciences Education
◯ Don't know

Which school are they in?

◯ School of Biological Sciences
◯ School of Biochemistry
◯ School of Cellular and Molecular Medicine
◯ School of Physiology, Pharmacology and Neuroscience
◯ School of Psychological Science
◯ Don't know

Which school are they in?

◯ School of Chemistry
◯ School of Earth Sciences
◯ School of Geographical Sciences
◯ School of Mathematics
◯ School of Physics
◯ Don't know

Which school are they in?

◯ School of Education
◯ School for Policy Studies
◯ School of Economics, Finance and Management
◯ School of Sociology, Politics and International Studies
◯ University of Bristol Law School
◯ Don't know

Where were you when you spoke to this person?

◯ Home
◯ Another home
◯ University
◯ Work or volunteering not at university
◯ School or nursery
◯ Shopping
◯ Medical or health centre
◯ Transport
◯ Place of worship
◯ Social (pub/nightclub/café/restaurant)
◯ Exercise
◯ Park
◯ Other location

Other location - please specify

__________________________________

Was this inside or outside?

◯ Inside
◯ Outside

How long did you talk to this person for?

◯ Less than 10 minutes
◯ Between 10 minutes and an hour
◯ Between 1 and 4 hours
◯ 4+ hours

How often would you expect to meet this person under current physical distancing measures?

◯ 4 or more days a week
◯ 2-3 days a week
◯ Once a week
◯ Less often than once a week
◯ Met for the first time this day

Did you also touch this person?

◯ Yes ◯ No

Person 18 Description of person:

__________________________________

◯ I did not speak to anyone else in person yesterday on a one-to-one basis.

Age:

◯ 0-4
◯ 5-17
◯ 18-24
◯ 25-44
◯ 45-64
◯ 65-80
◯ 81+

Is this person part of your household?

◯ Yes ◯ No

Does this person work or study at the University of Bristol?

◯ Yes ◯ No ◯ Don't know

What is the main faculty this person is associated with?

◯ Arts
◯ Engineering
◯ Health Sciences
◯ Life Sciences
◯ Science
◯ Social Sciences and Law
◯ Other
◯ Don't know

Which school are they in?

◯ Bristol Dental School
◯ Bristol Medical School
◯ Bristol Veterinary School
◯ Centre for Academic Language and Development
◯ Centre for Health Sciences Education
◯ Centre for Innovation
◯ School for Policy Studies
◯ School of Arts
◯ School of Biochemistry
◯ School of Biological Sciences
◯ School of Cellular and Molecular Medicine
◯ School of Chemistry
◯ School of Civil, Aerospace and Mechanical Engineering
◯ School of Computer Science, Electrical and Electronic Engineering, and Engineering Mathematics
◯ School of Earth Sciences
◯ School of Economics, Finance and Management
◯ School of Education
◯ School of Geographical Sciences
◯ School of Humanities
◯ School of Mathematics
◯ School of Modern Languages
◯ School of Physics
◯ School of Physiology, Pharmacology and Neuroscience
◯ School of Psychological Science
◯ School of Sociology, Politics and International Studies
◯ University of Bristol Law School
◯ Other
◯ Don't know

Other - please specify

__________________________________

Which school are they in?

◯ School of Arts
◯ School of Humanities
◯ School of Modern Languages
◯ Centre for Academic Language and Development
◯ Centre for Innovation
◯ Don't know

Which school are they in?

◯ School of Computer Science, Electrical and Electronic Engineering, and Engineering Mathematics
◯ School of Civil, Aerospace and Mechanical Engineering
◯ Don't know

Which school are they in?

◯ Bristol Dental School
◯ Bristol Medical School
◯ Bristol Veterinary School
◯ Centre for Health Sciences Education
◯ Don't know

Which school are they in?

◯ School of Biological Sciences
◯ School of Biochemistry
◯ School of Cellular and Molecular Medicine
◯ School of Physiology, Pharmacology and Neuroscience
◯ School of Psychological Science
◯ Don't know

Which school are they in?

◯ School of Chemistry
◯ School of Earth Sciences
◯ School of Geographical Sciences
◯ School of Mathematics
◯ School of Physics
◯ Don't know

Which school are they in?

◯ School of Education
◯ School for Policy Studies
◯ School of Economics, Finance and Management
◯ School of Sociology, Politics and International Studies
◯ University of Bristol Law School
◯ Don't know

Where were you when you spoke to this person?

◯ Home
◯ Another home
◯ University
◯ Work or volunteering not at university
◯ School or nursery
◯ Shopping
◯ Medical or health centre
◯ Transport
◯ Place of worship
◯ Social (pub/nightclub/café/restaurant)
◯ Exercise
◯ Park
◯ Other location

Other location - please specify

__________________________________

Was this inside or outside?

◯ Inside
◯ Outside

How long did you talk to this person for?

◯ Less than 10 minutes
◯ Between 10 minutes and an hour
◯ Between 1 and 4 hours
◯ 4+ hours

Did you also touch this person?

◯ Yes ◯ No

How often would you expect to meet this person under current physical distancing measures?

◯ 4 or more days a week
◯ 2-3 days a week
◯ Once a week
◯ Less often than once a week
◯ Met for the first time this day

Person 19 Description of person:

__________________________________

◯ I did not speak to anyone else in person yesterday on a one-to-one basis.

Age:

◯ 0-4
◯ 5-17
◯ 18-24
◯ 25-44
◯ 45-64
◯ 65-80
◯ 81+

Is this person part of your household?

◯ Yes ◯ No

Does this person work or study at the University of Bristol?

◯ Yes ◯ No ◯ Don't know

What is the main faculty this person is associated with?

◯ Arts
◯ Engineering
◯ Health Sciences
◯ Life Sciences
◯ Science
◯ Social Sciences and Law
◯ Other
◯ Don't know

Which school are they in?

◯ Bristol Dental School
◯ Bristol Medical School
◯ Bristol Veterinary School
◯ Centre for Academic Language and Development
◯ Centre for Health Sciences Education
◯ Centre for Innovation
◯ School for Policy Studies
◯ School of Arts
◯ School of Biochemistry
◯ School of Biological Sciences
◯ School of Cellular and Molecular Medicine
◯ School of Chemistry
◯ School of Civil, Aerospace and Mechanical Engineering
◯ School of Computer Science, Electrical and Electronic Engineering, and Engineering Mathematics
◯ School of Earth Sciences
◯ School of Economics, Finance and Management
◯ School of Education
◯ School of Geographical Sciences
◯ School of Humanities
◯ School of Mathematics
◯ School of Modern Languages
◯ School of Physics
◯ School of Physiology, Pharmacology and Neuroscience
◯ School of Psychological Science
◯ School of Sociology, Politics and International Studies
◯ University of Bristol Law School
◯ Other
◯ Don't know

---

Other - please specify

__________________________________

---

Which school are they in?

◯ School of Arts
◯ School of Humanities
◯ School of Modern Languages
◯ Centre for Academic Language and Development
◯ Centre for Innovation
◯ Don't know

---

Which school are they in?

◯ School of Computer Science, Electrical and Electronic Engineering, and Engineering Mathematics
◯ School of Civil, Aerospace and Mechanical Engineering
◯ Don't know

---

Which school are they in?

◯ Bristol Dental School
◯ Bristol Medical School
◯ Bristol Veterinary School
◯ Centre for Health Sciences Education
◯ Don't know

Which school are they in?

◯ School of Biological Sciences
◯ School of Biochemistry
◯ School of Cellular and Molecular Medicine
◯ School of Physiology, Pharmacology and Neuroscience
◯ School of Psychological Science
◯ Don't know

Which school are they in?

◯ School of Chemistry
◯ School of Earth Sciences
◯ School of Geographical Sciences
◯ School of Mathematics
◯ School of Physics
◯ Don't know

Which school are they in?

◯ School of Education
◯ School for Policy Studies
◯ School of Economics, Finance and Management
◯ School of Sociology, Politics and International Studies
◯ University of Bristol Law School
◯ Don't know

Where were you when you spoke to this person?

◯ Home
◯ Another home
◯ University
◯ Work or volunteering not at university
◯ School or nursery
◯ Shopping
◯ Medical or health centre
◯ Transport
◯ Place of worship
◯ Social (pub/nightclub/café/restaurant)
◯ Exercise
◯ Park
◯ Other location

Other location - please specify

__________________________________

Was this inside or outside?

◯ Inside
◯ Outside

How long did you talk to this person for?

◯ Less than 10 minutes
◯ Between 10 minutes and an hour
◯ Between 1 and 4 hours
◯ 4+ hours

Did you also touch this person?

◯ Yes ◯ No

How often would you expect to meet this person under current physical distancing measures?

◯ 4 or more days a week
◯ 2-3 days a week
◯ Once a week
◯ Less often than once a week
◯ Met for the first time this day

Person 20 Description of person:

__________________________________

◯ I did not speak to anyone else in person yesterday on a one-to-one basis.

Age:

◯ 0-4
◯ 5-17
◯ 18-24
◯ 25-44
◯ 45-64
◯ 65-80
◯ 81+

Is this person part of your household?

◯ Yes ◯ No

Does this person work or study at the University of Bristol?

◯ Yes ◯ No ◯ Don't know

What is the main faculty this person is associated with?

◯ Arts
◯ Engineering
◯ Health Sciences
◯ Life Sciences
◯ Science
◯ Social Sciences and Law
◯ Other
◯ Don't know

Which school are they in?

◯ Bristol Dental School
◯ Bristol Medical School
◯ Bristol Veterinary School
◯ Centre for Academic Language and Development
◯ Centre for Health Sciences Education
◯ Centre for Innovation
◯ School for Policy Studies
◯ School of Arts
◯ School of Biochemistry
◯ School of Biological Sciences
◯ School of Cellular and Molecular Medicine
◯ School of Chemistry
◯ School of Civil, Aerospace and Mechanical Engineering
◯ School of Computer Science, Electrical and Electronic Engineering, and Engineering Mathematics
◯ School of Earth Sciences
◯ School of Economics, Finance and Management
◯ School of Education
◯ School of Geographical Sciences
◯ School of Humanities
◯ School of Mathematics
◯ School of Modern Languages
◯ School of Physics
◯ School of Physiology, Pharmacology and Neuroscience
◯ School of Psychological Science
◯ School of Sociology, Politics and International Studies
◯ University of Bristol Law School
◯ Other
◯ Don't know

---

Other - please specify

__________________________________

---

Which school are they in?

◯ School of Arts
◯ School of Humanities
◯ School of Modern Languages
◯ Centre for Academic Language and Development
◯ Centre for Innovation
◯ Don't know

---

Which school are they in?

◯ School of Computer Science, Electrical and Electronic Engineering, and Engineering Mathematics
◯ School of Civil, Aerospace and Mechanical Engineering
◯ Don't know

---

Which school are they in?

◯ Bristol Dental School
◯ Bristol Medical School
◯ Bristol Veterinary School
◯ Centre for Health Sciences Education
◯ Don't know

Which school are they in?

◯ School of Biological Sciences
◯ School of Biochemistry
◯ School of Cellular and Molecular Medicine
◯ School of Physiology, Pharmacology and Neuroscience
◯ School of Psychological Science
◯ Don't know

Which school are they in?

◯ School of Chemistry
◯ School of Earth Sciences
◯ School of Geographical Sciences
◯ School of Mathematics
◯ School of Physics
◯ Don't know

Which school are they in?

◯ School of Education
◯ School for Policy Studies
◯ School of Economics, Finance and Management
◯ School of Sociology, Politics and International Studies
◯ University of Bristol Law School
◯ Don't know

Where were you when you spoke to this person?

◯ Home
◯ Another home
◯ University
◯ Work or volunteering not at university
◯ School or nursery
◯ Shopping
◯ Medical or health centre
◯ Transport
◯ Place of worship
◯ Social (pub/nightclub/café/restaurant)
◯ Exercise
◯ Park
◯ Other location

Other location - please specify

__________________________________

Was this inside or outside?

◯ Inside
◯ Outside

How long did you talk to this person for?

◯ Less than 10 minutes
◯ Between 10 minutes and an hour
◯ Between 1 and 4 hours
◯ 4+ hours

Did you also touch this person?

◯ Yes ◯ No

How often would you expect to meet this person under current physical distancing measures?

◯ 4 or more days a week
◯ 2-3 days a week
◯ Once a week
◯ Less often than once a week
◯ Met for the first time this day

Person 21 Description of person:

__________________________________

◯ I did not speak to anyone else in person yesterday on a one-to-one basis.

Age:

◯ 0-4
◯ 5-17
◯ 18-24
◯ 25-44
◯ 45-64
◯ 65-80
◯ 81+

Is this person part of your household?

◯ Yes ◯ No

Does this person work or study at the University of Bristol?

◯ Yes ◯ No ◯ Don't know

What is the main faculty this person is associated with?

◯ Arts
◯ Engineering
◯ Health Sciences
◯ Life Sciences
◯ Science
◯ Social Sciences and Law
◯ Other
◯ Don't know

Which school are they in?

◯ Bristol Dental School
◯ Bristol Medical School
◯ Bristol Veterinary School
◯ Centre for Academic Language and Development
◯ Centre for Health Sciences Education
◯ Centre for Innovation
◯ School for Policy Studies
◯ School of Arts
◯ School of Biochemistry
◯ School of Biological Sciences
◯ School of Cellular and Molecular Medicine
◯ School of Chemistry
◯ School of Civil, Aerospace and Mechanical Engineering
◯ School of Computer Science, Electrical and Electronic Engineering, and Engineering Mathematics
◯ School of Earth Sciences
◯ School of Economics, Finance and Management
◯ School of Education
◯ School of Geographical Sciences
◯ School of Humanities
◯ School of Mathematics
◯ School of Modern Languages
◯ School of Physics
◯ School of Physiology, Pharmacology and Neuroscience
◯ School of Psychological Science
◯ School of Sociology, Politics and International Studies
◯ University of Bristol Law School
◯ Other
◯ Don't know

Other - please specify

__________________________________

Which school are they in?

◯ School of Arts
◯ School of Humanities
◯ School of Modern Languages
◯ Centre for Academic Language and Development
◯ Centre for Innovation
◯ Don't know

Which school are they in?

◯ School of Computer Science, Electrical and Electronic Engineering, and Engineering Mathematics
◯ School of Civil, Aerospace and Mechanical Engineering
◯ Don't know

Which school are they in?

◯ Bristol Dental School
◯ Bristol Medical School
◯ Bristol Veterinary School
◯ Centre for Health Sciences Education
◯ Don't know

Which school are they in?

◯ School of Biological Sciences
◯ School of Biochemistry
◯ School of Cellular and Molecular Medicine
◯ School of Physiology, Pharmacology and Neuroscience
◯ School of Psychological Science
◯ Don't know

Which school are they in?

◯ School of Chemistry
◯ School of Earth Sciences
◯ School of Geographical Sciences
◯ School of Mathematics
◯ School of Physics
◯ Don't know

Which school are they in?

◯ School of Education
◯ School for Policy Studies
◯ School of Economics, Finance and Management
◯ School of Sociology, Politics and International Studies
◯ University of Bristol Law School
◯ Don't know

Where were you when you spoke to this person?

◯ Home
◯ Another home
◯ University
◯ Work or volunteering not at university
◯ School or nursery
◯ Shopping
◯ Medical or health centre
◯ Transport
◯ Place of worship
◯ Social (pub/nightclub/café/restaurant)
◯ Exercise
◯ Park
◯ Other location

Other location - please specify

__________________________________

Was this inside or outside?

◯ Inside
◯ Outside

How long did you talk to this person for?

◯ Less than 10 minutes
◯ Between 10 minutes and an hour
◯ Between 1 and 4 hours
◯ 4+ hours

Did you also touch this person?

◯ Yes ◯ No

How often would you expect to meet this person under current physical distancing measures?

◯ 4 or more days a week
◯ 2-3 days a week
◯ Once a week
◯ Less often than once a week
◯ Met for the first time this day

Person 22 Description of person:

________________________________

◯ I did not speak to anyone else in person yesterday on a one-to-one basis.

Age:

◯ 0-4
◯ 5-17
◯ 18-24
◯ 25-44
◯ 45-64
◯ 65-80
◯ 81+

Is this person part of your household?

◯ Yes ◯ No

Does this person work or study at the University of Bristol?

◯ Yes ◯ No ◯ Don't know

What is the main faculty this person is associated with?

◯ Arts
◯ Engineering
◯ Health Sciences
◯ Life Sciences
◯ Science
◯ Social Sciences and Law
◯ Other
◯ Don't know

Which school are they in?

◯ Bristol Dental School
◯ Bristol Medical School
◯ Bristol Veterinary School
◯ Centre for Academic Language and Development
◯ Centre for Health Sciences Education
◯ Centre for Innovation
◯ School for Policy Studies
◯ School of Arts
◯ School of Biochemistry
◯ School of Biological Sciences
◯ School of Cellular and Molecular Medicine
◯ School of Chemistry
◯ School of Civil, Aerospace and Mechanical Engineering
◯ School of Computer Science, Electrical and Electronic Engineering, and Engineering Mathematics
◯ School of Earth Sciences
◯ School of Economics, Finance and Management
◯ School of Education
◯ School of Geographical Sciences
◯ School of Humanities
◯ School of Mathematics
◯ School of Modern Languages
◯ School of Physics
◯ School of Physiology, Pharmacology and Neuroscience
◯ School of Psychological Science
◯ School of Sociology, Politics and International Studies
◯ University of Bristol Law School
◯ Other
◯ Don't know

Other - please specify

__________________________________

Which school are they in?

◯ School of Arts
◯ School of Humanities
◯ School of Modern Languages
◯ Centre for Academic Language and Development
◯ Centre for Innovation
◯ Don't know

Which school are they in?

◯ School of Computer Science, Electrical and Electronic Engineering, and Engineering Mathematics
◯ School of Civil, Aerospace and Mechanical Engineering
◯ Don't know

Which school are they in?

◯ Bristol Dental School
◯ Bristol Medical School
◯ Bristol Veterinary School
◯ Centre for Health Sciences Education
◯ Don't know

Which school are they in?

◯ School of Biological Sciences
◯ School of Biochemistry
◯ School of Cellular and Molecular Medicine
◯ School of Physiology, Pharmacology and Neuroscience
◯ School of Psychological Science
◯ Don't know

Which school are they in?

◯ School of Chemistry
◯ School of Earth Sciences
◯ School of Geographical Sciences
◯ School of Mathematics
◯ School of Physics
◯ Don't know

Which school are they in?

◯ School of Education
◯ School for Policy Studies
◯ School of Economics, Finance and Management
◯ School of Sociology, Politics and International Studies
◯ University of Bristol Law School
◯ Don't know

Where were you when you spoke to this person?

◯ Home
◯ Another home
◯ University
◯ Work or volunteering not at university
◯ School or nursery
◯ Shopping
◯ Medical or health centre
◯ Transport
◯ Place of worship
◯ Social (pub/nightclub/café/restaurant)
◯ Exercise
◯ Park
◯ Other location

Other location - please specify

__________________________________

Was this inside or outside?

◯ Inside
◯ Outside

How long did you talk to this person for?

◯ Less than 10 minutes
◯ Between 10 minutes and an hour
◯ Between 1 and 4 hours
◯ 4+ hours

Did you also touch this person?

◯ Yes ◯ No

How often would you expect to meet this person under current physical distancing measures?

◯ 4 or more days a week
◯ 2-3 days a week
◯ Once a week
◯ Less often than once a week
◯ Met for the first time this day

Person 23 Description of person:

__________________________________

◯ I did not speak to anyone else in person yesterday on a one-to-one basis.

Age:

◯ 0-4
◯ 5-17
◯ 18-24
◯ 25-44
◯ 45-64
◯ 65-80
◯ 81+

Is this person part of your household?

◯ Yes ◯ No

Does this person work or study at the University of Bristol?

◯ Yes ◯ No ◯ Don't know

What is the main faculty this person is associated with?

◯ Arts
◯ Engineering
◯ Health Sciences
◯ Life Sciences
◯ Science
◯ Social Sciences and Law
◯ Other
◯ Don't know

Which school are they in?

◯ Bristol Dental School
◯ Bristol Medical School
◯ Bristol Veterinary School
◯ Centre for Academic Language and Development
◯ Centre for Health Sciences Education
◯ Centre for Innovation
◯ School for Policy Studies
◯ School of Arts
◯ School of Biochemistry
◯ School of Biological Sciences
◯ School of Cellular and Molecular Medicine
◯ School of Chemistry
◯ School of Civil, Aerospace and Mechanical Engineering
◯ School of Computer Science, Electrical and Electronic Engineering, and Engineering Mathematics
◯ School of Earth Sciences
◯ School of Economics, Finance and Management
◯ School of Education
◯ School of Geographical Sciences
◯ School of Humanities
◯ School of Mathematics
◯ School of Modern Languages
◯ School of Physics
◯ School of Physiology, Pharmacology and Neuroscience
◯ School of Psychological Science
◯ School of Sociology, Politics and International Studies
◯ University of Bristol Law School
◯ Other
◯ Don't know

---

Other - please specify

__________________________________

---

Which school are they in?

◯ School of Arts
◯ School of Humanities
◯ School of Modern Languages
◯ Centre for Academic Language and Development
◯ Centre for Innovation
◯ Don't know

---

Which school are they in?

◯ School of Computer Science, Electrical and Electronic Engineering, and Engineering Mathematics
◯ School of Civil, Aerospace and Mechanical Engineering
◯ Don't know

---

Which school are they in?

◯ Bristol Dental School
◯ Bristol Medical School
◯ Bristol Veterinary School
◯ Centre for Health Sciences Education
◯ Don't know

Which school are they in?

◯ School of Biological Sciences
◯ School of Biochemistry
◯ School of Cellular and Molecular Medicine
◯ School of Physiology, Pharmacology and Neuroscience
◯ School of Psychological Science
◯ Don't know

Which school are they in?

◯ School of Chemistry
◯ School of Earth Sciences
◯ School of Geographical Sciences
◯ School of Mathematics
◯ School of Physics
◯ Don't know

Which school are they in?

◯ School of Education
◯ School for Policy Studies
◯ School of Economics, Finance and Management
◯ School of Sociology, Politics and International Studies
◯ University of Bristol Law School
◯ Don't know

Where were you when you spoke to this person?

◯ Home
◯ Another home
◯ University
◯ Work or volunteering not at university
◯ School or nursery
◯ Shopping
◯ Medical or health centre
◯ Transport
◯ Place of worship
◯ Social (pub/nightclub/café/restaurant)
◯ Exercise
◯ Park
◯ Other location

Other location - please specify

__________________________________

Was this inside or outside?

◯ Inside
◯ Outside

How long did you talk to this person for?

◯ Less than 10 minutes
◯ Between 10 minutes and an hour
◯ Between 1 and 4 hours
◯ 4+ hours

Did you also touch this person?

◯ Yes ◯ No

How often would you expect to meet this person under current physical distancing measures?

◯ 4 or more days a week
◯ 2-3 days a week
◯ Once a week
◯ Less often than once a week
◯ Met for the first time this day

Person 24 Description of person:

__________________________________

◯ I did not speak to anyone else in person yesterday on a one-to-one basis.

Age:

◯ 0-4
◯ 5-17
◯ 18-24
◯ 25-44
◯ 45-64
◯ 65-80
◯ 81+

Is this person part of your household?

◯ Yes ◯ No

Does this person work or study at the University of Bristol?

◯ Yes ◯ No ◯ Don't know

What is the main faculty this person is associated with?

◯ Arts
◯ Engineering
◯ Health Sciences
◯ Life Sciences
◯ Science
◯ Social Sciences and Law
◯ Other
◯ Don't know

Which school are they in?

◯ Bristol Dental School
◯ Bristol Medical School
◯ Bristol Veterinary School
◯ Centre for Academic Language and Development
◯ Centre for Health Sciences Education
◯ Centre for Innovation
◯ School for Policy Studies
◯ School of Arts
◯ School of Biochemistry
◯ School of Biological Sciences
◯ School of Cellular and Molecular Medicine
◯ School of Chemistry
◯ School of Civil, Aerospace and Mechanical Engineering
◯ School of Computer Science, Electrical and Electronic Engineering, and Engineering Mathematics
◯ School of Earth Sciences
◯ School of Economics, Finance and Management
◯ School of Education
◯ School of Geographical Sciences
◯ School of Humanities
◯ School of Mathematics
◯ School of Modern Languages
◯ School of Physics
◯ School of Physiology, Pharmacology and Neuroscience
◯ School of Psychological Science
◯ School of Sociology, Politics and International Studies
◯ University of Bristol Law School
◯ Other
◯ Don't know

---

Other - please specify

__________________________________

---

Which school are they in?

◯ School of Arts
◯ School of Humanities
◯ School of Modern Languages
◯ Centre for Academic Language and Development
◯ Centre for Innovation
◯ Don't know

---

Which school are they in?

◯ School of Computer Science, Electrical and Electronic Engineering, and Engineering Mathematics
◯ School of Civil, Aerospace and Mechanical Engineering
◯ Don't know

---

Which school are they in?

◯ Bristol Dental School
◯ Bristol Medical School
◯ Bristol Veterinary School
◯ Centre for Health Sciences Education
◯ Don't know

Which school are they in?

◯ School of Biological Sciences
◯ School of Biochemistry
◯ School of Cellular and Molecular Medicine
◯ School of Physiology, Pharmacology and Neuroscience
◯ School of Psychological Science
◯ Don't know

Which school are they in?

◯ School of Chemistry
◯ School of Earth Sciences
◯ School of Geographical Sciences
◯ School of Mathematics
◯ School of Physics
◯ Don't know

Which school are they in?

◯ School of Education
◯ School for Policy Studies
◯ School of Economics, Finance and Management
◯ School of Sociology, Politics and International Studies
◯ University of Bristol Law School
◯ Don't know

Where were you when you spoke to this person?

◯ Home
◯ Another home
◯ University
◯ Work or volunteering not at university
◯ School or nursery
◯ Shopping
◯ Medical or health centre
◯ Transport
◯ Place of worship
◯ Social (pub/nightclub/café/restaurant)
◯ Exercise
◯ Park
◯ Other location

Other location - please specify

__________________________________

Was this inside or outside?

◯ Inside
◯ Outside

How long did you talk to this person for?

◯ Less than 10 minutes
◯ Between 10 minutes and an hour
◯ Between 1 and 4 hours
◯ 4+ hours

Did you also touch this person?

◯ Yes ◯ No

How often would you expect to meet this person under current physical distancing measures?

◯ 4 or more days a week
◯ 2-3 days a week
◯ Once a week
◯ Less often than once a week
◯ Met for the first time this day

Person 25 Description of person:

__________________________________

◯ I did not speak to anyone else in person yesterday on a one-to-one basis.

Age:

◯ 0-4
◯ 5-17
◯ 18-24
◯ 25-44
◯ 45-64
◯ 65-80
◯ 81+

Is this person part of your household?

◯ Yes ◯ No

Does this person work or study at the University of Bristol?

◯ Yes ◯ No ◯ Don't know

What is the main faculty this person is associated with?

◯ Arts
◯ Engineering
◯ Health Sciences
◯ Life Sciences
◯ Science
◯ Social Sciences and Law
◯ Other
◯ Don't know

Which school are they in?

◯ Bristol Dental School
◯ Bristol Medical School
◯ Bristol Veterinary School
◯ Centre for Academic Language and Development
◯ Centre for Health Sciences Education
◯ Centre for Innovation
◯ School for Policy Studies
◯ School of Arts
◯ School of Biochemistry
◯ School of Biological Sciences
◯ School of Cellular and Molecular Medicine
◯ School of Chemistry
◯ School of Civil, Aerospace and Mechanical Engineering
◯ School of Computer Science, Electrical and Electronic Engineering, and Engineering Mathematics
◯ School of Earth Sciences
◯ School of Economics, Finance and Management
◯ School of Education
◯ School of Geographical Sciences
◯ School of Humanities
◯ School of Mathematics
◯ School of Modern Languages
◯ School of Physics
◯ School of Physiology, Pharmacology and Neuroscience
◯ School of Psychological Science
◯ School of Sociology, Politics and International Studies
◯ University of Bristol Law School
◯ Other
◯ Don't know

Other - please specify

__________________________________

Which school are they in?

◯ School of Arts
◯ School of Humanities
◯ School of Modern Languages
◯ Centre for Academic Language and Development
◯ Centre for Innovation
◯ Don't know

Which school are they in?

◯ School of Computer Science, Electrical and Electronic Engineering, and Engineering Mathematics
◯ School of Civil, Aerospace and Mechanical Engineering
◯ Don't know

Which school are they in?

◯ Bristol Dental School
◯ Bristol Medical School
◯ Bristol Veterinary School
◯ Centre for Health Sciences Education
◯ Don't know

Which school are they in?

◯ School of Biological Sciences
◯ School of Biochemistry
◯ School of Cellular and Molecular Medicine
◯ School of Physiology, Pharmacology and Neuroscience
◯ School of Psychological Science
◯ Don't know

Which school are they in?

◯ School of Chemistry
◯ School of Earth Sciences
◯ School of Geographical Sciences
◯ School of Mathematics
◯ School of Physics
◯ Don't know

Which school are they in?

◯ School of Education
◯ School for Policy Studies
◯ School of Economics, Finance and Management
◯ School of Sociology, Politics and International Studies
◯ University of Bristol Law School
◯ Don't know

Where were you when you spoke to this person?

◯ Home
◯ Another home
◯ University
◯ Work or volunteering not at university
◯ School or nursery
◯ Shopping
◯ Medical or health centre
◯ Transport
◯ Place of worship
◯ Social (pub/nightclub/café/restaurant)
◯ Exercise
◯ Park
◯ Other location

Other location - please specify

__________________________________

Was this inside or outside?

◯ Inside
◯ Outside

How long did you talk to this person for?

◯ Less than 10 minutes
◯ Between 10 minutes and an hour
◯ Between 1 and 4 hours
◯ 4+ hours

Did you also touch this person?

◯ Yes ◯ No

How often would you expect to meet this person under current physical distancing measures?

◯ 4 or more days a week
◯ 2-3 days a week
◯ Once a week
◯ Less often than once a week
◯ Met for the first time this day

Person 26 Description of person:

__________________________________

◯ I did not speak to anyone else in person yesterday on a one-to-one basis.

Age:

◯ 0-4
◯ 5-17
◯ 18-24
◯ 25-44
◯ 45-64
◯ 65-80
◯ 81+

Is this person part of your household?

◯ Yes ◯ No

Does this person work or study at the University of Bristol?

◯ Yes ◯ No ◯ Don't know

What is the main faculty this person is associated with?

◯ Arts
◯ Engineering
◯ Health Sciences
◯ Life Sciences
◯ Science
◯ Social Sciences and Law
◯ Other
◯ Don't know

Which school are they in?

◯ Bristol Dental School
◯ Bristol Medical School
◯ Bristol Veterinary School
◯ Centre for Academic Language and Development
◯ Centre for Health Sciences Education
◯ Centre for Innovation
◯ School for Policy Studies
◯ School of Arts
◯ School of Biochemistry
◯ School of Biological Sciences
◯ School of Cellular and Molecular Medicine
◯ School of Chemistry
◯ School of Civil, Aerospace and Mechanical Engineering
◯ School of Computer Science, Electrical and Electronic Engineering, and Engineering Mathematics
◯ School of Earth Sciences
◯ School of Economics, Finance and Management
◯ School of Education
◯ School of Geographical Sciences
◯ School of Humanities
◯ School of Mathematics
◯ School of Modern Languages
◯ School of Physics
◯ School of Physiology, Pharmacology and Neuroscience
◯ School of Psychological Science
◯ School of Sociology, Politics and International Studies
◯ University of Bristol Law School
◯ Other
◯ Don't know

---

Other - please specify

__________________________________

---

Which school are they in?

◯ School of Arts
◯ School of Humanities
◯ School of Modern Languages
◯ Centre for Academic Language and Development
◯ Centre for Innovation
◯ Don't know

---

Which school are they in?

◯ School of Computer Science, Electrical and Electronic Engineering, and Engineering Mathematics
◯ School of Civil, Aerospace and Mechanical Engineering
◯ Don't know

---

Which school are they in?

◯ Bristol Dental School
◯ Bristol Medical School
◯ Bristol Veterinary School
◯ Centre for Health Sciences Education
◯ Don't know

Which school are they in?

◯ School of Biological Sciences
◯ School of Biochemistry
◯ School of Cellular and Molecular Medicine
◯ School of Physiology, Pharmacology and Neuroscience
◯ School of Psychological Science
◯ Don't know

Which school are they in?

◯ School of Chemistry
◯ School of Earth Sciences
◯ School of Geographical Sciences
◯ School of Mathematics
◯ School of Physics
◯ Don't know

Which school are they in?

◯ School of Education
◯ School for Policy Studies
◯ School of Economics, Finance and Management
◯ School of Sociology, Politics and International Studies
◯ University of Bristol Law School
◯ Don't know

Where were you when you spoke to this person?

◯ Home
◯ Another home
◯ University
◯ Work or volunteering not at university
◯ School or nursery
◯ Shopping
◯ Medical or health centre
◯ Transport
◯ Place of worship
◯ Social (pub/nightclub/café/restaurant)
◯ Exercise
◯ Park
◯ Other location

Other location - please specify

__________________________________

Was this inside or outside?

◯ Inside
◯ Outside

How long did you talk to this person for?

◯ Less than 10 minutes
◯ Between 10 minutes and an hour
◯ Between 1 and 4 hours
◯ 4+ hours

Did you also touch this person?

◯ Yes ◯ No

How often would you expect to meet this person under current physical distancing measures?

◯ 4 or more days a week
◯ 2-3 days a week
◯ Once a week
◯ Less often than once a week
◯ Met for the first time this day

Person 27 Description of person:

__________________________________

◯ I did not speak to anyone else in person yesterday on a one-to-one basis.

Age:

◯ 0-4
◯ 5-17
◯ 18-24
◯ 25-44
◯ 45-64
◯ 65-80
◯ 81+

Is this person part of your household?

◯ Yes ◯ No

Does this person work or study at the University of Bristol?

◯ Yes ◯ No ◯ Don't know

What is the main faculty this person is associated with?

◯ Arts
◯ Engineering
◯ Health Sciences
◯ Life Sciences
◯ Science
◯ Social Sciences and Law
◯ Other
◯ Don't know

Which school are they in?

◯ Bristol Dental School
◯ Bristol Medical School
◯ Bristol Veterinary School
◯ Centre for Academic Language and Development
◯ Centre for Health Sciences Education
◯ Centre for Innovation
◯ School for Policy Studies
◯ School of Arts
◯ School of Biochemistry
◯ School of Biological Sciences
◯ School of Cellular and Molecular Medicine
◯ School of Chemistry
◯ School of Civil, Aerospace and Mechanical Engineering
◯ School of Computer Science, Electrical and Electronic Engineering, and Engineering Mathematics
◯ School of Earth Sciences
◯ School of Economics, Finance and Management
◯ School of Education
◯ School of Geographical Sciences
◯ School of Humanities
◯ School of Mathematics
◯ School of Modern Languages
◯ School of Physics
◯ School of Physiology, Pharmacology and Neuroscience
◯ School of Psychological Science
◯ School of Sociology, Politics and International Studies
◯ University of Bristol Law School
◯ Other
◯ Don't know

---

Other - please specify

__________________________________

---

Which school are they in?

◯ School of Arts
◯ School of Humanities
◯ School of Modern Languages
◯ Centre for Academic Language and Development
◯ Centre for Innovation
◯ Don't know

---

Which school are they in?

◯ School of Computer Science, Electrical and Electronic Engineering, and Engineering Mathematics
◯ School of Civil, Aerospace and Mechanical Engineering
◯ Don't know

---

Which school are they in?

◯ Bristol Dental School
◯ Bristol Medical School
◯ Bristol Veterinary School
◯ Centre for Health Sciences Education
◯ Don't know

Which school are they in?

◯ School of Biological Sciences
◯ School of Biochemistry
◯ School of Cellular and Molecular Medicine
◯ School of Physiology, Pharmacology and Neuroscience
◯ School of Psychological Science
◯ Don't know

Which school are they in?

◯ School of Chemistry
◯ School of Earth Sciences
◯ School of Geographical Sciences
◯ School of Mathematics
◯ School of Physics
◯ Don't know

Which school are they in?

◯ School of Education
◯ School for Policy Studies
◯ School of Economics, Finance and Management
◯ School of Sociology, Politics and International Studies
◯ University of Bristol Law School
◯ Don't know

Where were you when you spoke to this person?

◯ Home
◯ Another home
◯ University
◯ Work or volunteering not at university
◯ School or nursery
◯ Shopping
◯ Medical or health centre
◯ Transport
◯ Place of worship
◯ Social (pub/nightclub/café/restaurant)
◯ Exercise
◯ Park
◯ Other location

Other location - please specify

__________________________________

Was this inside or outside?

◯ Inside
◯ Outside

How long did you talk to this person for?

◯ Less than 10 minutes
◯ Between 10 minutes and an hour
◯ Between 1 and 4 hours
◯ 4+ hours

Did you also touch this person?

◯ Yes ◯ No

How often would you expect to meet this person under current physical distancing measures?

◯ 4 or more days a week
◯ 2-3 days a week
◯ Once a week
◯ Less often than once a week
◯ Met for the first time this day

Person 28 Description of person:

__________________________________

◯ I did not speak to anyone else in person yesterday on a one-to-one basis.

Age:

◯ 0-4
◯ 5-17
◯ 18-24
◯ 25-44
◯ 45-64
◯ 65-80
◯ 81+

Is this person part of your household?

◯ Yes ◯ No

Does this person work or study at the University of Bristol?

◯ Yes ◯ No ◯ Don't know

What is the main faculty this person is associated with?

◯ Arts
◯ Engineering
◯ Health Sciences
◯ Life Sciences
◯ Science
◯ Social Sciences and Law
◯ Other
◯ Don't know

Which school are they in?

◯ Bristol Dental School
◯ Bristol Medical School
◯ Bristol Veterinary School
◯ Centre for Academic Language and Development
◯ Centre for Health Sciences Education
◯ Centre for Innovation
◯ School for Policy Studies
◯ School of Arts
◯ School of Biochemistry
◯ School of Biological Sciences
◯ School of Cellular and Molecular Medicine
◯ School of Chemistry
◯ School of Civil, Aerospace and Mechanical Engineering
◯ School of Computer Science, Electrical and Electronic Engineering, and Engineering Mathematics
◯ School of Earth Sciences
◯ School of Economics, Finance and Management
◯ School of Education
◯ School of Geographical Sciences
◯ School of Humanities
◯ School of Mathematics
◯ School of Modern Languages
◯ School of Physics
◯ School of Physiology, Pharmacology and Neuroscience
◯ School of Psychological Science
◯ School of Sociology, Politics and International Studies
◯ University of Bristol Law School
◯ Other
◯ Don't know

---

Other - please specify

__________________________________

---

Which school are they in?

◯ School of Arts
◯ School of Humanities
◯ School of Modern Languages
◯ Centre for Academic Language and Development
◯ Centre for Innovation
◯ Don't know

---

Which school are they in?

◯ School of Computer Science, Electrical and Electronic Engineering, and Engineering Mathematics
◯ School of Civil, Aerospace and Mechanical Engineering
◯ Don't know

---

Which school are they in?

◯ Bristol Dental School
◯ Bristol Medical School
◯ Bristol Veterinary School
◯ Centre for Health Sciences Education
◯ Don't know

Which school are they in?

◯ School of Biological Sciences
◯ School of Biochemistry
◯ School of Cellular and Molecular Medicine
◯ School of Physiology, Pharmacology and Neuroscience
◯ School of Psychological Science
◯ Don't know

Which school are they in?

◯ School of Chemistry
◯ School of Earth Sciences
◯ School of Geographical Sciences
◯ School of Mathematics
◯ School of Physics
◯ Don't know

Which school are they in?

◯ School of Education
◯ School for Policy Studies
◯ School of Economics, Finance and Management
◯ School of Sociology, Politics and International Studies
◯ University of Bristol Law School
◯ Don't know

Where were you when you spoke to this person?

◯ Home
◯ Another home
◯ University
◯ Work or volunteering not at university
◯ School or nursery
◯ Shopping
◯ Medical or health centre
◯ Transport
◯ Place of worship
◯ Social (pub/nightclub/café/restaurant)
◯ Exercise
◯ Park
◯ Other location

Other location - please specify

__________________________________

Was this inside or outside?

◯ Inside
◯ Outside

How long did you talk to this person for?

◯ Less than 10 minutes
◯ Between 10 minutes and an hour
◯ Between 1 and 4 hours
◯ 4+ hours

Did you also touch this person?

◯ Yes ◯ No

How often would you expect to meet this person under current physical distancing measures?

◯ 4 or more days a week
◯ 2-3 days a week
◯ Once a week
◯ Less often than once a week
◯ Met for the first time this day

Person 29 Description of person:

__________________________________

◯ I did not speak to anyone else in person yesterday on a one-to-one basis.

Age:

◯ 0-4
◯ 5-17
◯ 18-24
◯ 25-44
◯ 45-64
◯ 65-80
◯ 81+

Is this person part of your household?

◯ Yes ◯ No

Does this person work or study at the University of Bristol?

◯ Yes ◯ No ◯ Don't know

What is the main faculty this person is associated with?

◯ Arts
◯ Engineering
◯ Health Sciences
◯ Life Sciences
◯ Science
◯ Social Sciences and Law
◯ Other
◯ Don't know

Which school are they in?

◯ Bristol Dental School
◯ Bristol Medical School
◯ Bristol Veterinary School
◯ Centre for Academic Language and Development
◯ Centre for Health Sciences Education
◯ Centre for Innovation
◯ School for Policy Studies
◯ School of Arts
◯ School of Biochemistry
◯ School of Biological Sciences
◯ School of Cellular and Molecular Medicine
◯ School of Chemistry
◯ School of Civil, Aerospace and Mechanical Engineering
◯ School of Computer Science, Electrical and Electronic Engineering, and Engineering Mathematics
◯ School of Earth Sciences
◯ School of Economics, Finance and Management
◯ School of Education
◯ School of Geographical Sciences
◯ School of Humanities
◯ School of Mathematics
◯ School of Modern Languages
◯ School of Physics
◯ School of Physiology, Pharmacology and Neuroscience
◯ School of Psychological Science
◯ School of Sociology, Politics and International Studies
◯ University of Bristol Law School
◯ Other
◯ Don't know

---

Other - please specify

__________________________________

---

Which school are they in?

◯ School of Arts
◯ School of Humanities
◯ School of Modern Languages
◯ Centre for Academic Language and Development
◯ Centre for Innovation
◯ Don't know

---

Which school are they in?

◯ School of Computer Science, Electrical and Electronic Engineering, and Engineering Mathematics
◯ School of Civil, Aerospace and Mechanical Engineering
◯ Don't know

---

Which school are they in?

◯ Bristol Dental School
◯ Bristol Medical School
◯ Bristol Veterinary School
◯ Centre for Health Sciences Education
◯ Don't know

Which school are they in?

◯ School of Biological Sciences
◯ School of Biochemistry
◯ School of Cellular and Molecular Medicine
◯ School of Physiology, Pharmacology and Neuroscience
◯ School of Psychological Science
◯ Don't know

Which school are they in?

◯ School of Chemistry
◯ School of Earth Sciences
◯ School of Geographical Sciences
◯ School of Mathematics
◯ School of Physics
◯ Don't know

Which school are they in?

◯ School of Education
◯ School for Policy Studies
◯ School of Economics, Finance and Management
◯ School of Sociology, Politics and International Studies
◯ University of Bristol Law School
◯ Don't know

Where were you when you spoke to this person?

◯ Home
◯ Another home
◯ University
◯ Work or volunteering not at university
◯ School or nursery
◯ Shopping
◯ Medical or health centre
◯ Transport
◯ Place of worship
◯ Social (pub/nightclub/café/restaurant)
◯ Exercise
◯ Park
◯ Other location

Other location - please specify

__________________________________

Was this inside or outside?

◯ Inside
◯ Outside

How long did you talk to this person for?

◯ Less than 10 minutes
◯ Between 10 minutes and an hour
◯ Between 1 and 4 hours
◯ 4+ hours

Did you also touch this person?

◯ Yes ◯ No

How often would you expect to meet this person under current physical distancing measures?

◯ 4 or more days a week
◯ 2-3 days a week
◯ Once a week
◯ Less often than once a week
◯ Met for the first time this day

Person 30 Description of person:

__________________________________

◯ I did not speak to anyone else in person yesterday on a one-to-one basis.

Age:

◯ 0-4
◯ 5-17
◯ 18-24
◯ 25-44
◯ 45-64
◯ 65-80
◯ 81+

Is this person part of your household?

◯ Yes ◯ No

Does this person work or study at the University of Bristol?

◯ Yes ◯ No

What is the main faculty this person is associated with?

◯ Arts
◯ Engineering
◯ Health Sciences
◯ Life Sciences
◯ Science
◯ Social Sciences and Law
◯ Other
◯ Don't know

Which school are they in?

◯ Bristol Dental School
◯ Bristol Medical School
◯ Bristol Veterinary School
◯ Centre for Academic Language and Development
◯ Centre for Health Sciences Education
◯ Centre for Innovation
◯ School for Policy Studies
◯ School of Arts
◯ School of Biochemistry
◯ School of Biological Sciences
◯ School of Cellular and Molecular Medicine
◯ School of Chemistry
◯ School of Civil, Aerospace and Mechanical Engineering
◯ School of Computer Science, Electrical and Electronic Engineering, and Engineering Mathematics
◯ School of Earth Sciences
◯ School of Economics, Finance and Management
◯ School of Education
◯ School of Geographical Sciences
◯ School of Humanities
◯ School of Mathematics
◯ School of Modern Languages
◯ School of Physics
◯ School of Physiology, Pharmacology and Neuroscience
◯ School of Psychological Science
◯ School of Sociology, Politics and International Studies
◯ University of Bristol Law School
◯ Other
◯ Don't know

---

Other - please specify

__________________________________

---

Which school are they in?

◯ School of Arts
◯ School of Humanities
◯ School of Modern Languages
◯ Centre for Academic Language and Development
◯ Centre for Innovation
◯ Don't know

---

Which school are they in?

◯ School of Computer Science, Electrical and Electronic Engineering, and Engineering Mathematics
◯ School of Civil, Aerospace and Mechanical Engineering
◯ Don't know

---

Which school are they in?

◯ Bristol Dental School
◯ Bristol Medical School
◯ Bristol Veterinary School
◯ Centre for Health Sciences Education
◯ Don't know

Which school are they in?

◯ School of Biological Sciences
◯ School of Biochemistry
◯ School of Cellular and Molecular Medicine
◯ School of Physiology, Pharmacology and Neuroscience
◯ School of Psychological Science
◯ Don't know

Which school are they in?

◯ School of Chemistry
◯ School of Earth Sciences
◯ School of Geographical Sciences
◯ School of Mathematics
◯ School of Physics
◯ Don't know

Which school are they in?

◯ School of Education
◯ School for Policy Studies
◯ School of Economics, Finance and Management
◯ School of Sociology, Politics and International Studies
◯ University of Bristol Law School
◯ Don't know

Where were you when you spoke to this person?

◯ Home
◯ Another home
◯ University
◯ Work or volunteering not at university
◯ School or nursery
◯ Shopping
◯ Medical or health centre
◯ Transport
◯ Place of worship
◯ Social (pub/nightclub/café/restaurant)
◯ Exercise
◯ Park
◯ Other location

Other location - please specify

__________________________________

Was this inside or outside?

◯ Inside
◯ Outside

How long did you talk to this person for?

◯ Less than 10 minutes
◯ Between 10 minutes and an hour
◯ Between 1 and 4 hours
◯ 4+ hours

Did you also touch this person?

◯ Yes ◯ No

How often would you expect to meet this person under current physical distancing measures?

◯ 4 or more days a week
◯ 2-3 days a week
◯ Once a week
◯ Less often than once a week
◯ Met for the first time this day

Additional conversational or physical contacts (e.g. as part of a customer service role)
If you had any additional individual contacts where you had a conversation with a person or had physical contact, please enter the number of contacts below. For example, if you work in a shop and had conversations with 24 customers in a day, enter 24. If you do not know the exact number, please enter your best estimate.

__________________________________

Group contacts yesterday In addition to the people you listed in question 4, did you meet with any large groups yesterday (for example, sports teams, tutorials, lectures, religious services, large gatherings with friends and family)?

◯ Yes ◯ No, I didn't meet with any groups yesterday

Group 1 Description:

__________________________________

How many people were in the group in each age category?

Age 0-4:

__________________________________

Age 5-17:

__________________________________

Age 18-24:

__________________________________

Age 25-44:

__________________________________

Age 45-64:

__________________________________

Age 65-80:

__________________________________

Age 81+

__________________________________

Do the majority of this group work or study at the University of Bristol?

◯ Yes ◯ No ◯ Don't know

What is the main faculty this group is associated with?

◯ Arts
◯ Engineering
◯ Health Sciences
◯ Life Sciences
◯ Science
◯ Social Sciences and Law
◯ Other
◯ Don't know

What is the main school this group is associated with?

◯ Bristol Dental School
◯ Bristol Medical School
◯ Bristol Veterinary School
◯ Centre for Academic Language and Development
◯ Centre for Health Sciences Education
◯ Centre for Innovation
◯ School for Policy Studies
◯ School of Arts
◯ School of Biochemistry
◯ School of Biological Sciences
◯ School of Cellular and Molecular Medicine
◯ School of Chemistry
◯ School of Civil, Aerospace and Mechanical Engineering
◯ School of Computer Science, Electrical and Electronic Engineering, and Engineering Mathematics
◯ School of Earth Sciences
◯ School of Economics, Finance and Management
◯ School of Education
◯ School of Geographical Sciences
◯ School of Humanities
◯ School of Mathematics
◯ School of Modern Languages
◯ School of Physics
◯ School of Physiology, Pharmacology and Neuroscience
◯ School of Psychological Science
◯ School of Sociology, Politics and International Studies
◯ University of Bristol Law School
◯ Other
◯ Don't know

Other - please specify

__________________________________

Which school are they in?

◯ School of Arts
◯ School of Humanities
◯ School of Modern Languages
◯ Centre for Academic Language and Development
◯ Centre for Innovation
◯ Don't know

Which school are they in?

◯ School of Computer Science, Electrical and Electronic Engineering, and Engineering Mathematics
◯ School of Civil, Aerospace and Mechanical Engineering
◯ Don't know

Which school are they in?

◯ Bristol Dental School
◯ Bristol Medical School
◯ Bristol Veterinary School
◯ Centre for Health Sciences Education
◯ Don't know

Which school are they in?

◯ School of Biological Sciences
◯ School of Biochemistry
◯ School of Cellular and Molecular Medicine
◯ School of Physiology, Pharmacology and Neuroscience
◯ School of Psychological Science
◯ Don't know

Which school are they in?

◯ School of Chemistry
◯ School of Earth Sciences
◯ School of Geographical Sciences
◯ School of Mathematics
◯ School of Physics
◯ Don't know

Which school are they in?

◯ School of Education
◯ School for Policy Studies
◯ School of Economics, Finance and Management
◯ School of Sociology, Politics and International Studies
◯ University of Bristol Law School
◯ Don't know

Where were you when you spoke to these people?

◯ Home
◯ Another home
◯ University
◯ Work or volunteering not at university
◯ School or nursery
◯ Shopping
◯ Medical or health centre
◯ Transport
◯ Place of worship
◯ Social (pub/nightclub/café/restaurant)
◯ Exercise
◯ Park
◯ Other location

Other location - please specify

__________________________________

Was this inside or outside?

◯ Inside ◯ Outside ◯ Both inside and outside

How long did you talk to this group for?

◯ Less than 10 minutes
◯ Between 10 minutes and an hour
◯ Between 1 and 4 hours
◯ 4+ hours

Did the members of the group also talk to each other?

◯ Yes ◯ No

How often would you expect to meet this group under current physical distancing measures?

◯ 4 or more days a week
◯ 2-3 days a week
◯ Once a week
◯ Less often than once a week
◯ Met for the first time this day

Group 2 Description:

__________________________________

◯ I didn't meet any other groups yesterday.

How many people were in the group in each age category?

Age 0-4:

__________________________________

Age 5-17:

__________________________________

Age 18-24:

__________________________________

Age 25-44:

__________________________________

Age 45-64:

__________________________________

Age 65-80:

__________________________

Age 81+

__________________________

Do the majority of this group work or study at the University of Bristol?

○ Yes ○ No ○ Don't know

What is the main faculty this group is associated with?

○ Arts
○ Engineering
○ Health Sciences
○ Life Sciences
○ Science
○ Social Sciences and Law
○ Other
○ Don't know

What is the main school this group is associated with?

○ Bristol Dental School
○ Bristol Medical School
○ Bristol Veterinary School
○ Centre for Academic Language and Development
○ Centre for Health Sciences Education
○ Centre for Innovation
○ School for Policy Studies
○ School of Arts
○ School of Biochemistry
○ School of Biological Sciences
○ School of Cellular and Molecular Medicine
○ School of Chemistry
○ School of Civil, Aerospace and Mechanical Engineering
○ School of Computer Science, Electrical and Electronic Engineering, and Engineering Mathematics
○ School of Earth Sciences
○ School of Economics, Finance and Management
○ School of Education
○ School of Geographical Sciences
○ School of Humanities
○ School of Mathematics
○ School of Modern Languages
○ School of Physics
○ School of Physiology, Pharmacology and Neuroscience
○ School of Psychological Science
○ School of Sociology, Politics and International Studies
○ University of Bristol Law School
○ Other
○ Don't know

Other - please specify

__________________________

Which school are they in?

◯ School of Arts
◯ School of Humanities
◯ School of Modern Languages
◯ Centre for Academic Language and Development
◯ Centre for Innovation
◯ Don't know

Which school are they in?

◯ School of Computer Science, Electrical and Electronic Engineering, and Engineering Mathematics
◯ School of Civil, Aerospace and Mechanical Engineering
◯ Don't know

Which school are they in?

◯ Bristol Dental School
◯ Bristol Medical School
◯ Bristol Veterinary School
◯ Centre for Health Sciences Education
◯ Don't know

Which school are they in?

◯ School of Biological Sciences
◯ School of Biochemistry
◯ School of Cellular and Molecular Medicine
◯ School of Physiology, Pharmacology and Neuroscience
◯ School of Psychological Science
◯ Don't know

Which school are they in?

◯ School of Chemistry
◯ School of Earth Sciences
◯ School of Geographical Sciences
◯ School of Mathematics
◯ School of Physics
◯ Don't know

Which school are they in?

◯ School of Education
◯ School for Policy Studies
◯ School of Economics, Finance and Management
◯ School of Sociology, Politics and International Studies
◯ University of Bristol Law School
◯ Don't know

Where were you when you spoke to these people?

◯ Home
◯ Another home
◯ University
◯ Work or volunteering not at university
◯ School or nursery
◯ Shopping
◯ Medical or health centre
◯ Transport
◯ Place of worship
◯ Social (pub/nightclub/café/restaurant)
◯ Exercise
◯ Park
◯ Other location

Other location - please specify

__________________________________

Was this inside or outside?

◯ Inside ◯ Outside ◯ Both inside and outside

How long did you talk to this group for?

◯ Less than 10 minutes
◯ Between 10 minutes and an hour
◯ Between 1 and 4 hours
◯ 4+ hours

Did the members of the group also talk to each other?

◯ Yes ◯ No

Group 3 Description:

__________________________________

◯ I didn't meet any other groups yesterday.

How many people were in the group in each age category?

Age 0-4:

__________________________________

Age 5-17:

__________________________________

Age 18-24:

__________________________________

Age 25-44:

__________________________________

Age 45-64:

__________________________________

Age 65-80:

__________________________________

Age 81+

__________________________________

Do the majority of this group work or study at the University of Bristol?

◯ Yes ◯ No ◯ Don't know

What is the main faculty this group is associated with?

◯ Arts
◯ Engineering
◯ Health Sciences
◯ Life Sciences
◯ Science
◯ Social Sciences and Law
◯ Other
◯ Don't know

What is the main school this group is associated with?

◯ Bristol Dental School
◯ Bristol Medical School
◯ Bristol Veterinary School
◯ Centre for Academic Language and Development
◯ Centre for Health Sciences Education
◯ Centre for Innovation
◯ School for Policy Studies
◯ School of Arts
◯ School of Biochemistry
◯ School of Biological Sciences
◯ School of Cellular and Molecular Medicine
◯ School of Chemistry
◯ School of Civil, Aerospace and Mechanical Engineering
◯ School of Computer Science, Electrical and Electronic Engineering, and Engineering Mathematics
◯ School of Earth Sciences
◯ School of Economics, Finance and Management
◯ School of Education
◯ School of Geographical Sciences
◯ School of Humanities
◯ School of Mathematics
◯ School of Modern Languages
◯ School of Physics
◯ School of Physiology, Pharmacology and Neuroscience
◯ School of Psychological Science
◯ School of Sociology, Politics and International Studies
◯ University of Bristol Law School
◯ Other
◯ Don't know

---

Other - please specify

__________________________________

---

Which school are they in?

◯ School of Arts
◯ School of Humanities
◯ School of Modern Languages
◯ Centre for Academic Language and Development
◯ Centre for Innovation
◯ Don't know

---

Which school are they in?

◯ School of Computer Science, Electrical and Electronic Engineering, and Engineering Mathematics
◯ School of Civil, Aerospace and Mechanical Engineering
◯ Don't know

---

Which school are they in?

◯ Bristol Dental School
◯ Bristol Medical School
◯ Bristol Veterinary School
◯ Centre for Health Sciences Education
◯ Don't know

Which school are they in?

◯ School of Biological Sciences
◯ School of Biochemistry
◯ School of Cellular and Molecular Medicine
◯ School of Physiology, Pharmacology and Neuroscience
◯ School of Psychological Science
◯ Don't know

Which school are they in?

◯ School of Chemistry
◯ School of Earth Sciences
◯ School of Geographical Sciences
◯ School of Mathematics
◯ School of Physics
◯ Don't know

Which school are they in?

◯ School of Education
◯ School for Policy Studies
◯ School of Economics, Finance and Management
◯ School of Sociology, Politics and International Studies
◯ University of Bristol Law School
◯ Don't know

Where were you when you spoke to these people?

◯ Home
◯ Another home
◯ University
◯ Work or volunteering not at university
◯ School or nursery
◯ Shopping
◯ Medical or health centre
◯ Transport
◯ Place of worship
◯ Social (pub/nightclub/café/restaurant)
◯ Exercise
◯ Park
◯ Other location

Other location - please specify

__________________________________

Was this inside or outside?

◯ Inside ◯ Outside ◯ Both inside and outside

How long did you talk to this group for?

◯ Less than 10 minutes
◯ Between 10 minutes and an hour
◯ Between 1 and 4 hours
◯ 4+ hours

Did the members of the group also talk to each other?

◯ Yes ◯ No

Group 4 Description:

______________________________

◯ I didn't meet any other groups yesterday.

How many people were in the group in each age category?

Age 0-4:

______________________________

Age 5-17:

______________________________

Age 18-24:

______________________________

Age 25-44:

______________________________

Age 45-64:

______________________________

Age 65-80:

______________________________

Age 81+

______________________________

Do the majority of this group work or study at the University of Bristol?

◯ Yes ◯ No ◯ Don't know

What is the main faculty this group is associated with?

◯ Arts
◯ Engineering
◯ Health Sciences
◯ Life Sciences
◯ Science
◯ Social Sciences and Law
◯ Other
◯ Don't know

What is the main school this group is associated with?

◯ Bristol Dental School
◯ Bristol Medical School
◯ Bristol Veterinary School
◯ Centre for Academic Language and Development
◯ Centre for Health Sciences Education
◯ Centre for Innovation
◯ School for Policy Studies
◯ School of Arts
◯ School of Biochemistry
◯ School of Biological Sciences
◯ School of Cellular and Molecular Medicine
◯ School of Chemistry
◯ School of Civil, Aerospace and Mechanical Engineering
◯ School of Computer Science, Electrical and Electronic Engineering, and Engineering Mathematics
◯ School of Earth Sciences
◯ School of Economics, Finance and Management
◯ School of Education
◯ School of Geographical Sciences
◯ School of Humanities
◯ School of Mathematics
◯ School of Modern Languages
◯ School of Physics
◯ School of Physiology, Pharmacology and Neuroscience
◯ School of Psychological Science
◯ School of Sociology, Politics and International Studies
◯ University of Bristol Law School
◯ Other
◯ Don't know

Other - please specify

__________________________________

Which school are they in?

◯ School of Arts
◯ School of Humanities
◯ School of Modern Languages
◯ Centre for Academic Language and Development
◯ Centre for Innovation
◯ Don't know

Which school are they in?

◯ School of Computer Science, Electrical and Electronic Engineering, and Engineering Mathematics
◯ School of Civil, Aerospace and Mechanical Engineering
◯ Don't know

Which school are they in?

◯ Bristol Dental School
◯ Bristol Medical School
◯ Bristol Veterinary School
◯ Centre for Health Sciences Education
◯ Don't know

Which school are they in?

◯ School of Biological Sciences
◯ School of Biochemistry
◯ School of Cellular and Molecular Medicine
◯ School of Physiology, Pharmacology and Neuroscience
◯ School of Psychological Science
◯ Don't know

Which school are they in?

◯ School of Chemistry
◯ School of Earth Sciences
◯ School of Geographical Sciences
◯ School of Mathematics
◯ School of Physics
◯ Don't know

Which school are they in?

◯ School of Education
◯ School for Policy Studies
◯ School of Economics, Finance and Management
◯ School of Sociology, Politics and International Studies
◯ University of Bristol Law School
◯ Don't know

Where were you when you spoke to these people?

◯ Home
◯ Another home
◯ University
◯ Work or volunteering not at university
◯ School or nursery
◯ Shopping
◯ Medical or health centre
◯ Transport
◯ Place of worship
◯ Social (pub/nightclub/café/restaurant)
◯ Exercise
◯ Park
◯ Other location

Other location - please specify

__________________________________

Was this inside or outside?

◯ Inside ◯ Outside ◯ Both inside and outside

How long did you talk to this group for?

◯ Less than 10 minutes
◯ Between 10 minutes and an hour
◯ Between 1 and 4 hours
◯ 4+ hours

Did the members of the group also talk to each other?

◯ Yes ◯ No

Group 5 Description:

__________________________________

How many people were in the group in each age category?

◯ I didn't meet any other groups yesterday.

Age 0-4:

__________________________________

Age 5-17:

__________________________________

Age 18-24:

__________________________________

Age 25-44:

__________________________________

Age 45-64:

__________________________________

Age 65-80:

__________________________________

Age 81+

__________________________________

Do the majority of this group work or study at the University of Bristol?

◯ Yes ◯ No ◯ Don't know

What is the main faculty this group is associated with?

◯ Arts
◯ Engineering
◯ Health Sciences
◯ Life Sciences
◯ Science
◯ Social Sciences and Law
◯ Other
◯ Don't know

What is the main school this group is associated with?

◯ Bristol Dental School
◯ Bristol Medical School
◯ Bristol Veterinary School
◯ Centre for Academic Language and Development
◯ Centre for Health Sciences Education
◯ Centre for Innovation
◯ School for Policy Studies
◯ School of Arts
◯ School of Biochemistry
◯ School of Biological Sciences
◯ School of Cellular and Molecular Medicine
◯ School of Chemistry
◯ School of Civil, Aerospace and Mechanical Engineering
◯ School of Computer Science, Electrical and Electronic Engineering, and Engineering Mathematics
◯ School of Earth Sciences
◯ School of Economics, Finance and Management
◯ School of Education
◯ School of Geographical Sciences
◯ School of Humanities
◯ School of Mathematics
◯ School of Modern Languages
◯ School of Physics
◯ School of Physiology, Pharmacology and Neuroscience
◯ School of Psychological Science
◯ School of Sociology, Politics and International Studies
◯ University of Bristol Law School
◯ Other
◯ Don't know

Other - please specify

__________________________________

Which school are they in?

◯ School of Arts
◯ School of Humanities
◯ School of Modern Languages
◯ Centre for Academic Language and Development
◯ Centre for Innovation
◯ Don't know

Which school are they in?

◯ School of Computer Science, Electrical and Electronic Engineering, and Engineering Mathematics
◯ School of Civil, Aerospace and Mechanical Engineering
◯ Don't know

Which school are they in?

◯ Bristol Dental School
◯ Bristol Medical School
◯ Bristol Veterinary School
◯ Centre for Health Sciences Education
◯ Don't know

Which school are they in?

◯ School of Biological Sciences
◯ School of Biochemistry
◯ School of Cellular and Molecular Medicine
◯ School of Physiology, Pharmacology and Neuroscience
◯ School of Psychological Science
◯ Don't know

Which school are they in?

◯ School of Chemistry
◯ School of Earth Sciences
◯ School of Geographical Sciences
◯ School of Mathematics
◯ School of Physics
◯ Don't know

Which school are they in?

◯ School of Education
◯ School for Policy Studies
◯ School of Economics, Finance and Management
◯ School of Sociology, Politics and International Studies
◯ University of Bristol Law School
◯ Don't know

Where were you when you spoke to these people?

◯ Home
◯ Another home
◯ University
◯ Work or volunteering not at university
◯ School or nursery
◯ Shopping
◯ Medical or health centre
◯ Transport
◯ Place of worship
◯ Social (pub/nightclub/café/restaurant)
◯ Exercise
◯ Park
◯ Other location

Other location - please specify

__________________________________

Was this inside or outside?

◯ Inside ◯ Outside ◯ Both inside and outside

How long did you talk to this group for?

◯ Less than 10 minutes
◯ Between 10 minutes and an hour
◯ Between 1 and 4 hours
◯ 4+ hours

Did the members of the group also talk to each other?

◯ Yes ◯ No

Additional conversational or physical contacts Description:If you had any additional group contacts where you had a conversation with a group of people or had physical contact, please enter the number of group contacts below. If you do not know the exact number, please enter your best estimate.

__________________________________

◯ I didn't meet any other groups yesterday.
